# Healthy vaccinee effect in SARS-CoV-2 vaccinees without documented infection

**DOI:** 10.64898/2026.09.04.26362245

**Authors:** Uwe Riedmann, Hermann Brenner, John P.A. Ioannidis, Stefan Pilz

## Abstract

**Importance:** Estimates of SARS-CoV-2 vaccine effectiveness (VE) from observational studies may be biased if healthier (healthy vaccinee effect; HVE) or less healthy individuals are preferentially vaccinated. Quantifying HVE is essential for interpreting VE estimates for informed vaccination policy.

**Objective:** To assess the HVE by comparing all-cause mortality, cancer mortality, and deaths from external causes between vaccinated and unvaccinated individuals.

**Design, Setting, and Participants:** This nationwide retrospective cohort study used Austrian national registry data from 2021 to August 31, 2022, and included the entire adult population of Austria with no previously documented SARS-CoV-2 infection on January 1, 2021. Cox regression was used to estimate hazard ratios (HRs) for mortality comparing vaccinated with unvaccinated individuals while adjusting for age, sex and Austrian long-term care allowance (ALTCA) recipiency, stratified by vaccinations (1 to 4). Analyses were performed for the whole observation period, 3-month blocks (quarters) and the 2 months with highest and lowest COVID-19 disease burden per year. Additionally, newly vaccinated individuals were matched on day of vaccination based on the aforementioned covariates plus highest education and HRs were assed for the two weeks following vaccination.

**Exposure:** Receipt of at least 1 dose of SARS-CoV-2 vaccine.

**Main Outcomes and Measures:** All-cause mortality, cancer mortality, and deaths from external causes (e.g., falls and transport accidents).

**Results:** Among 7,416,077 eligible adults, all-cause mortality was lower among vaccinated than unvaccinated individuals regardless of vaccination number. Differences in risk were greatest shortly after vaccination and attenuated over time. E.g., adjusted all-cause HRs in the quarter after vaccination introduction were 0.46, 0.52, 0.38 and 0.43 for vaccinated vs unvaccinated individuals for 1, 2, 3 and 4 vaccinations, respectively. Respective HRs for the first 2 weeks after vaccination in the matched analysis were 0.24-0.32 across all vaccinations. Findings for cancer mortality and deaths from external causes paralleled those for all-cause mortality.

**Conclusions and Relevance:** A pronounced HVE was observed for SARS-CoV-2 vaccination during the pandemic, evidenced by markedly lower mortality from causes not plausibly related to vaccination (cancer, external causes) among vaccinated individuals. This bias needs to be accounted for when interpreting observational estimates of SARS-CoV-2 VE.

## INTRODUCTION

In early stages of the SARS-CoV-2 pandemic, randomized controlled trials (RCTs) indicated high efficacy of SARS-CoV-2 vaccines against COVID-19 infections.^1^ Since then, a large number of mostly observational studies have assessed the relationships of SARS-CoV-2 vaccinations with a variety of medical outcomes, and often reported beneficial effects even for outcomes that had not been thought to be related to COVID-19 infections.^2–6^ For example, a recent cohort study among US Veterans reported substantially larger absolute reduction in major adverse cardiovascular events (MACE) than in COVID-19-associated MACE (more than 11x higher).^6^ The authors attributed this unexpected finding to the extension of the vaccine’s protective association to the hidden burden of undetected SARS-CoV-2 and its sequelae. However, an alternative explanation that continues to receive little attention in the literature is the healthy vaccinee effect (HVE).^2–4,7–9^

The “healthy vaccinee effect” occurs when individuals who get vaccinated have generally better health outcomes than those who do not, leading to an overestimation of vaccine effectiveness (VE).^10–15^ Conversely, individuals with compromised health may be more likely to be vaccinated, coined “confounding by indication”.^10^ These biases oppose each other and may be present simultaneously with varying magnitudes. Capturing health differences between vaccinated and unvaccinated via demographic data is challenging, as they are likely due to often unmeasured factors such as lifestyle, timing of vaccination, or functional status limitations.^10,15,16^

Assessing mortality unrelated to the target disease in vaccinated versus unvaccinated individuals during times of low infection rates has been proposed to evaluate the HVE.^17^ This approach estimates the net effect of the HVE, confounding by indication and other related biases. To understand these biases, one should remove the effect of demographics, since these vaccines may have been prioritized and used differentially by different age groups in different time periods. Moreover, these biases may be explained by differences in comorbidities and socioeconomic factors.^9,10,12^

In a previous nationwide study from Austria, we demonstrated potential HVE to be substantial among previously SARS-CoV-2 infected individuals from Austria, we observed strong indication for a HVE evidenced by significantly lower all-cause, non-COVID-19 and cancer mortality in vaccinated versus unvaccinated individuals. The magnitude of this effect was biggest at the time of vaccination, but waned thereafter.^3^ Whether these findings also apply to individuals with no previously documented infection is currently unknown.

In this nationwide cohort study in all Austrian adults, we compare all-cause mortality in vaccinated versus unvaccinated individuals with no documented previous infection, between January 2021 and September 2022. We also analyse the months with low and high COVID-19 mortality rates, as they are differentially affected by any misclassification of death causes (COVID-19 versus non-COVID-19). Additionally, we match vaccinated with unvaccinated individuals to assess differences in mortality for the first two weeks after vaccination, i.e., before reasonable onset of vaccination effects. Finally, cancer deaths and deaths by external causes are evaluated as negative control outcomes that are assumed to be unrelated to COVID-19 and vaccination.

## METHODS

### Study design, procedures, and participants

We conducted a nationwide retrospective observational study in all Austrian adults (aged ≥ 18 years) from the start of mass vaccinations in January 2021 till the September 2022 (Figure 1). We used national health data from the Austrian Micro Data Center (AMDC) of Statistics Austria.^18^ The dataset contains information on age, sex, date of SARS-CoV-2 infections (up to September 2022) vaccinations against SARS-CoV-2 (up to the fourth vaccination/dose; including date of vaccination and vaccine product), vital status regarding deaths, cause of death, Austrian long-term care allowance recipiency (ALTCA)^19^ and highest education level for every individual registered in Austria, as well as information on individual migration.^3,20^ Education level was categorized in five levels: compulsory schooling (9 years) or lower; pre-secondary education; secondary education; pre-tertiary education; and tertiary education. Detection of SARS-CoV-2 infections was originally provided by the Austrian epidemiological reporting system (German: *Epidemiologisches Meldesystem*; EMS).^21^

**Figure 1:**
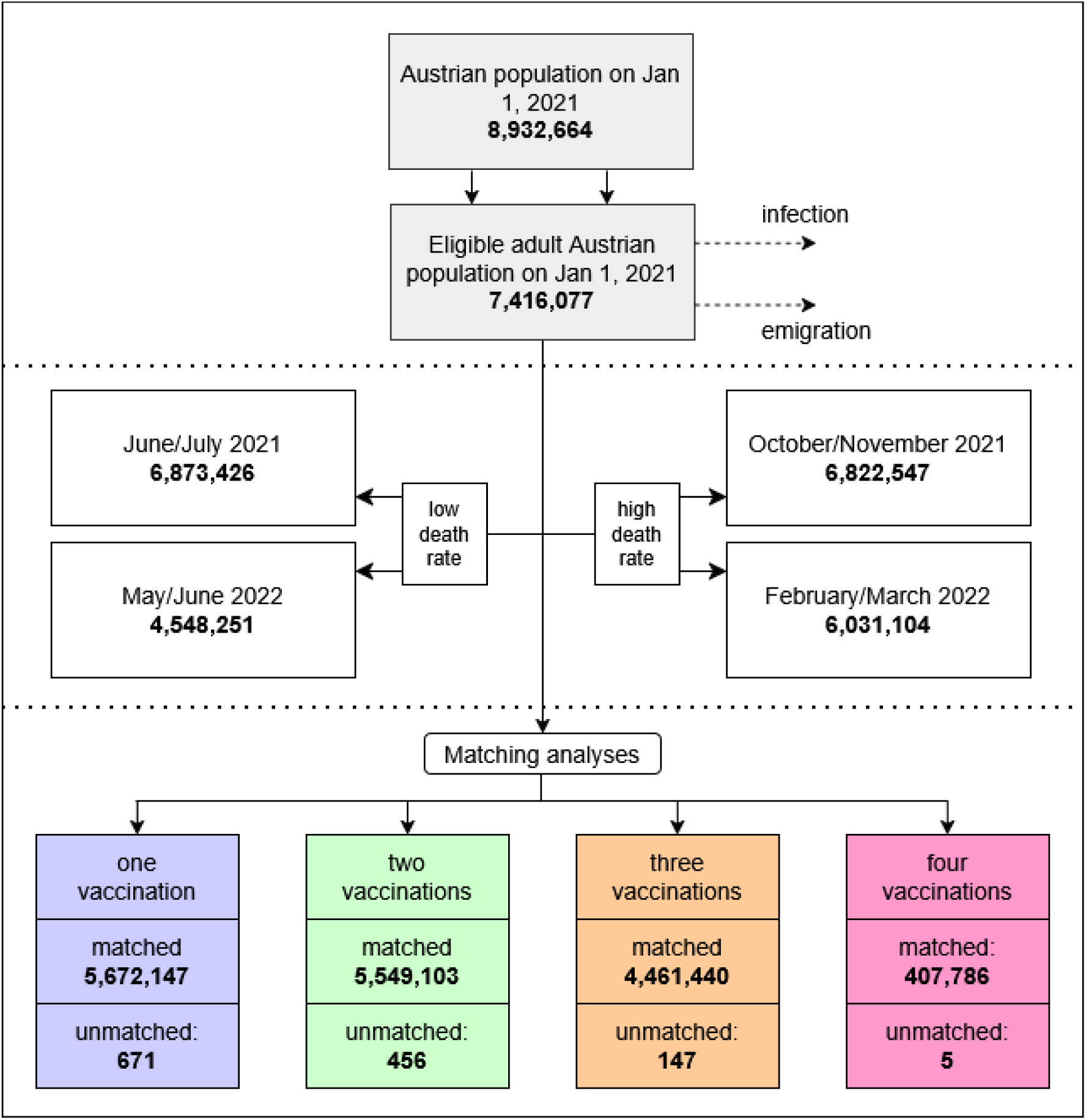
Participant selection flow-chart. Note that all unmatched cases were due to missing data on education.

We followed the Strengthening the Reporting of Observational Studies in Epidemiology (STROBE) checklist (Table S1). A nationwide dataset was used and thus no sample size calculation was performed. Ethical approval was obtained from the ethics committee at the Medical University of Graz, Austria (no. 33-144 ex 20/21).

### Statistical analysis

Categorical data are presented as percentages, and continuous data are shown as medians (with 25^th^ to 75^th^ percentile). We used Cox regression analyses to calculate hazard ratios (HRs) with 95% confidence intervals (CI) for mortality in groups according to the number of vaccinations against SARS-CoV-2. All adults (≥18 years) without a previous documented infection on the respective start date were eligible. General censoring occurred at the end of the respective observation period, on the date of the first documented SARS-CoV-2 infection, the date of emigration or the date of death, whichever happened first. Individuals changed vaccination groups on the day of vaccination (e.g., unvaccinated to vaccinated once). Data on ALTCA was available yearly, thus individuals who received ALTCA within a year changed groups on January 1^st^ of that year. Statistical analyses were performed in R (version 4.4.3).^22^

### Mortality analyses in different time periods

For the entire observation period, we calculated HRs for 1, 2, 3 and 4 times vaccinated individuals versus unvaccinated individuals. We also split the observation period into 3 month blocks, as the previous study showed violation of the proportional hazard assumption when analysing longer observation periods.^3^ As adjusting variables, we considered age (years), sex (binary variable), level of education (five levels, previously described) and ALTCA recipiency (yes/no)^19^. We obtained crude (unadjusted) HRs; adjusted HRs (aHRs) adjusting for age and sex; aHRs adjusting for age, sex and ALTCA (primary outcome); and aHRs adjusting for age, sex, ALTCA and education. The age, sex and ALTCA adjusted analysis is the primary outcome as it accounts for factors of preferential vaccination, while the additional adjustment for education may capture part of the mechanism underlying the HVE itself, rather than acting as an independent confounder. Adjusting for it in the primary analysis would therefore partly remove the selection effect we’re trying to quantify, rather than clarify it. Education is thus treated as a mediator and reserved for sensitivity analyses. We additionally repeated the 3-month mortality analysis, comparing each vaccination number level with the preceding one as the reference group.

We specifically analysed the two consecutive months of the years 2021 and 2022 with the lowest COVID-19 mortality rates. This approach allows us to evaluate “off-season” group differences in times in which the virus is hardly circulating such that our findings should be negligibly driven by COVID-19 deaths. Such “off season” analyses are performed according to several previous studies on the HVE regarding influenza and COVID-19 vaccines.^3,10,17,23–25^ Additional analyses were performed in the respective two months with the highest COVID-19 mortality rates. If SARS-CoV-2 vaccines protect against non-COVID-19 mortality (e.g., due to post SARS-CoV-2 infection sequelae or death misclassifications), all-cause-mortality reductions in vaccinated versus unvaccinated individuals should be more pronounced in time periods with high versus low COVID-19 mortality.

### Sensitivity Analyses

For sensitivity analyses we stratified by sex, age groups (18-39, 40-59, 60-74, 75-84 and 85+ years old), education (five levels, previously described), ALTCA recipiency (yes/no) and mRNA vaccine (yes/no) of the last administered vaccination.

### Matching Analyses

We further investigated the period close to vaccination by matching (exact; 1 to 1; without replacement within the same day) individuals who received a vaccination for each calendar day to unvaccinated controls, based on sex, age group (18 to 24 years, then 5 years intervals up to 95+), education level and ALTCA recipiency. Eligible for the unvaccinated control group were only individuals with no documented previous vaccination or documented vaccination in the respective 14 days after the matching day (observation period). We additionally adjusted the analyses for age to address potential differences within matched age groups.

### Negative control outcome analyses

Analyses were performed for cancer mortality (ICD10: C00-C99) as negative control outcome, representing a proxy for worse physical health which should not be directly affected by vaccination. Thus, differences in cancer mortality between vaccinated and unvaccinated would indicate that vaccination prevalence differs according to people’s health status. Analyses were also performed for deaths by external causes (ICD10: V00-Y99).^26^ This includes for example accidents (transport accidents as well as falls and other accidents), assaults and self-harm.^26^ Some of the external causes are strongly related to physical health status (e.g. falls), mental health status (e.g. suicides), or socioeconomic circumstances (e.g. assaults), while others not necessarily (e.g. transport accidents).

## RESULTS

### Study population

The population in Austria was 8,932,664 (50.8% females) with a median age (25 to 75^th^ percentile) of 43 years (25-60) on January 1^st^, 2021. This included 7,416,077 (51 % females) adults (median age (25 to 75^th^ percentile): 49 years (33-63)) eligible for the main analysis. Participant flow-chart is shown in Figure 1. Population characteristics at different relevant timepoints are shown in Tables S2-S7.

### Analyses of overall observation period

Characteristics of the analysed population at the start and end of the observation period are presented in Table S2 and S3. A total of 118,008 were censored due to death in the eligible population between 1 January 2021 and 31 August 2022 (Table S8). Both overall and 3-month observation period estimates are presented in Figure 2 (Table 1, S9-S13).

**Figure 2:**
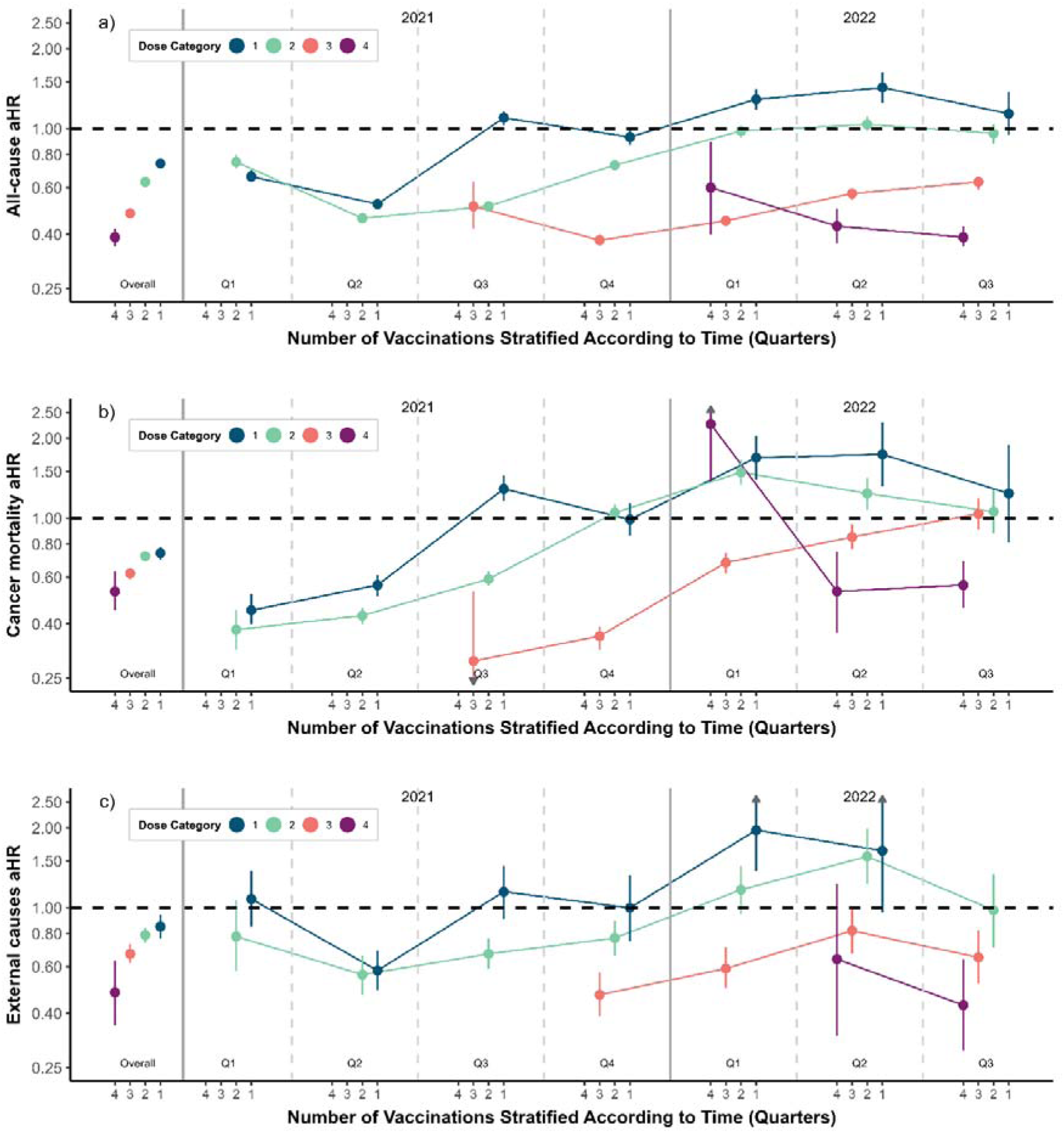
Age, sex and ALTCA recipiency adjusted all-cause-mortality (a), cancer (b) and external causes (c) HRs and 95% CIs for 1, 2, 3 and 4 vaccinations over the whole observation period and split into three-month periods. Unvaccinated individuals are the reference group. See Table 1, and Tables S9-S12 for details and crude estimates. Confidence values with lower bounds below 0.25 and upper bounds above 2.5 are indicated by a grey arrow.

**Table 1:**
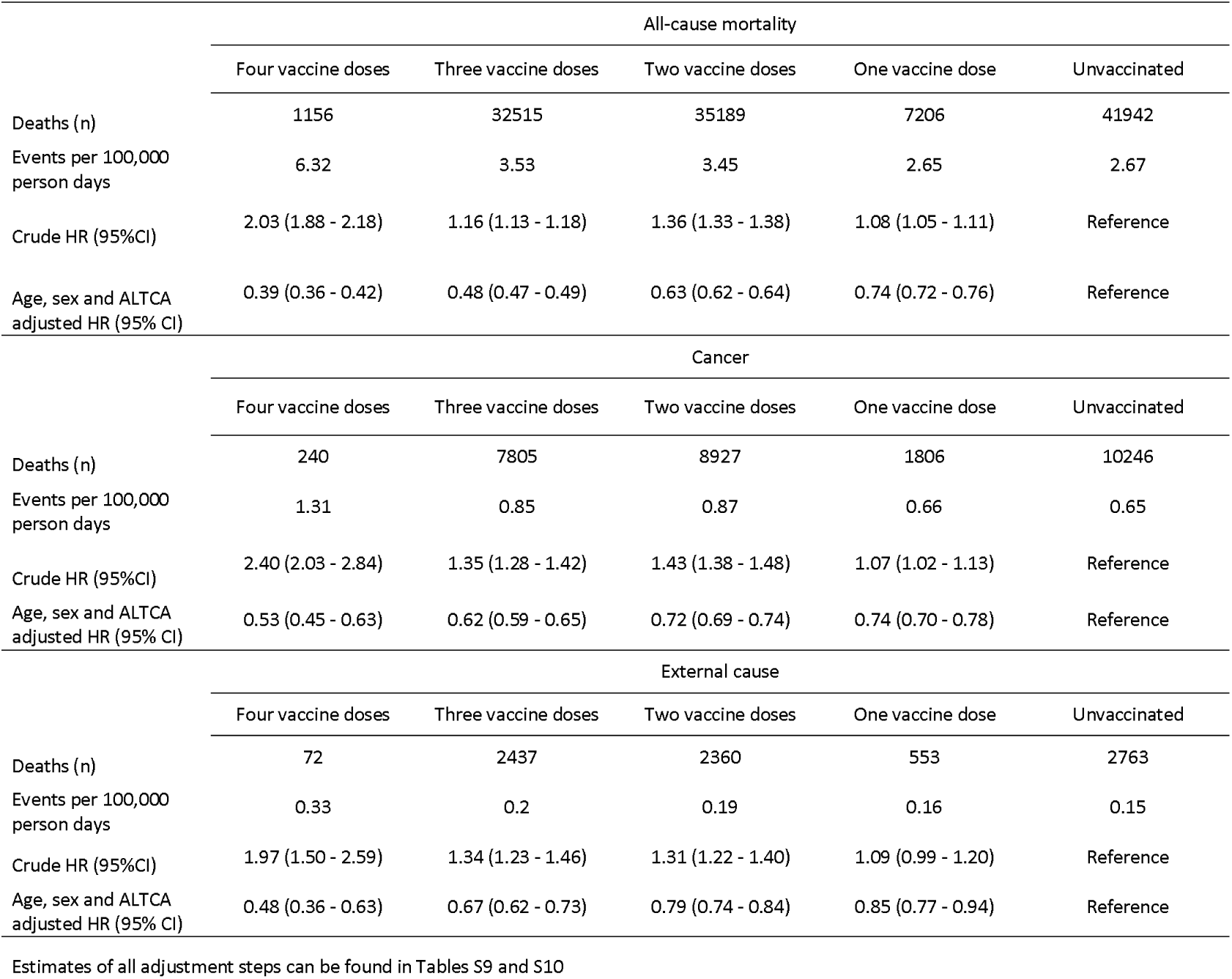
Hazard ratios (HR) with 95% confidence intervals (95% CI) for all-cause mortality, cancer deaths and deaths by external causes according to number of SARS-CoV-2 vaccine doses between 1, January 2021 and 31, August 2022.

The aHRs indicate lower all-cause mortality in vaccinated versus unvaccinated individuals irrespective of the number of vaccinations (Table 1 and Figure 2). Results were similar regardless of adjustments used (Tables S9-S13). Mortality was lower in the vaccinated compared to unvaccinated groups, but this difference diminished and partially reversed as time since first inception of the respective vaccination increased (Figure 2, Table S11 and S12). E.g., all-cause mortality in age, sex and ALTCA adjusted aHR for 1- and 2-vaccinations were 0.52 (0.50 - 0.54) and 0.46 (0.44 - 0.48) in Q2 2021, but became 1.14 (0.95 - 1.37) and 0.96 (0.88 - 1.04) in Q3 2022, while HRs of 3- and 4-vaccinations were at 0.63 (0.59 - 0.66) and 0.39 (0.36 - 0.43) in Q3 2022, respectively. Comparison of doses with the respective preceding dose show that earliest vaccinated populations have either not significantly different or higher risk than the previous doses but following quarters show lower all-cause mortality of subsequent doses (Figure S1 and Table S13).

### Analyses during periods with high and low COVID-19 mortality rates

Baseline characteristics and mortality rates of different periods with high and low COVID-19 mortality rates are shown in Tables S4-S7. Results of Cox regression analyses are shown in Table 2 (see Tables S14 and S15 for details). Vaccinated individuals had significantly decreased all-cause mortality risk in 2021, for both low and high COVID-19 mortality time periods. A similar pattern was present for 3 and 4 vaccinations in 2022, while 1 and 2 vaccinations were not significant or even indicating higher risk for vaccinated (Table 2).

**Table 2:**
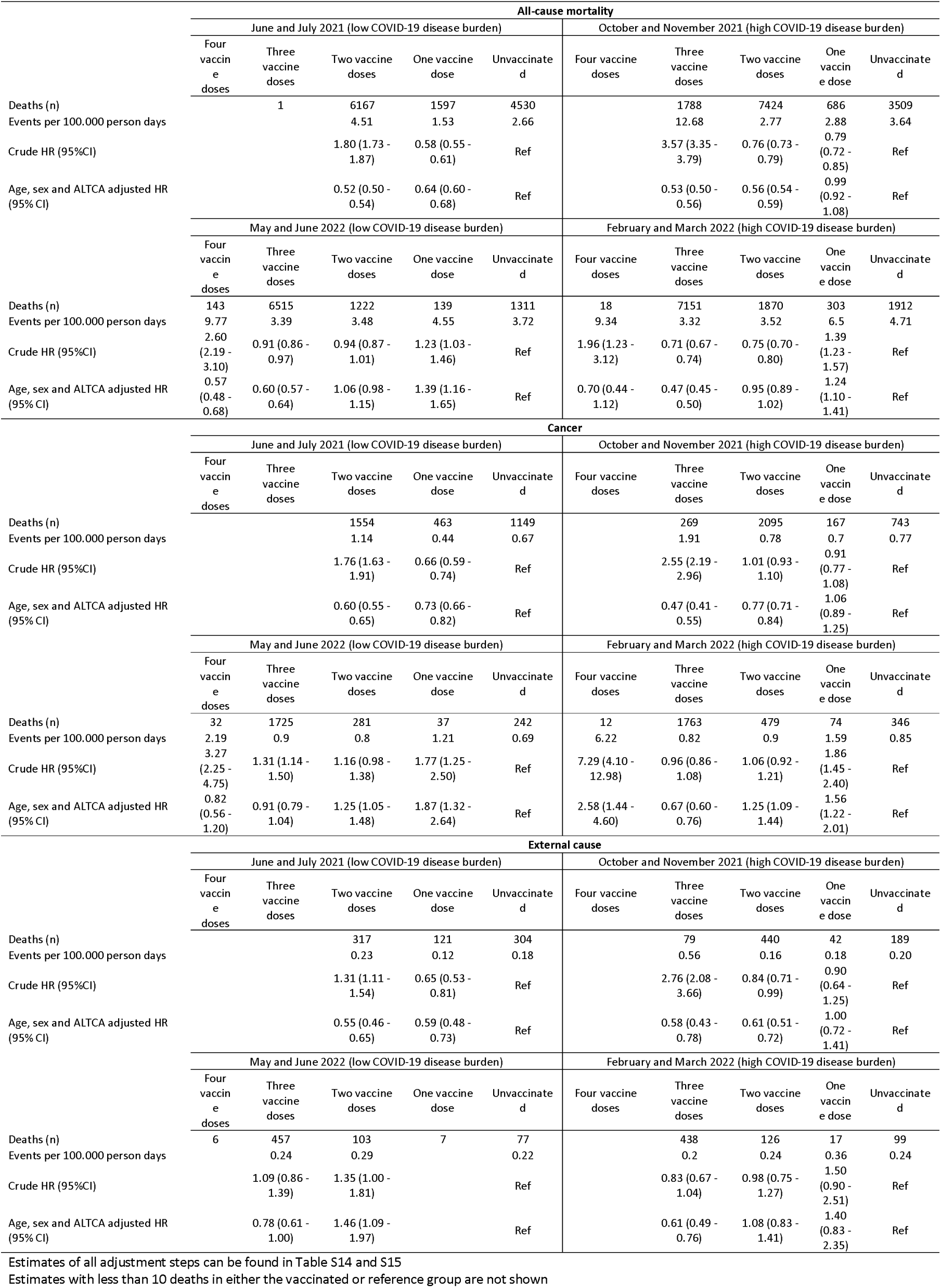
Hazard ratios (HR) with 95% confidence intervals (95% CI) for all-cause mortality, cancer deaths and deaths from external causes according to number of SARS-CoV-2 vaccine doses during different time periods of high and low COVID-19 disease burden.

### Sensitivity analyses

Stratified results of quarterly analyses, and periods of low and high COVID-19 mortality analyses for all-cause mortality can be found in Tables S16-S31 and Tables S32-S47, respectively. Sensitivity analyses outcomes closely mirror the main results independent of age group, sex, education level, ATLCA status and mRNA or non-mRNA vaccination (most recent vaccination). Across all analyses, the inclusion of education did not substantially change the estimates.

### Matched analyses

Matching analysis shows significantly reduced mortality risk in vaccinated compared to unvaccinated individuals within two weeks after vaccination. The HRs are very low (0.24-0.32) irrespective of vaccination numbers (Table 3). Both cancer mortality and external causes mortality mirrored these results (Table 3).

**Table 3:**
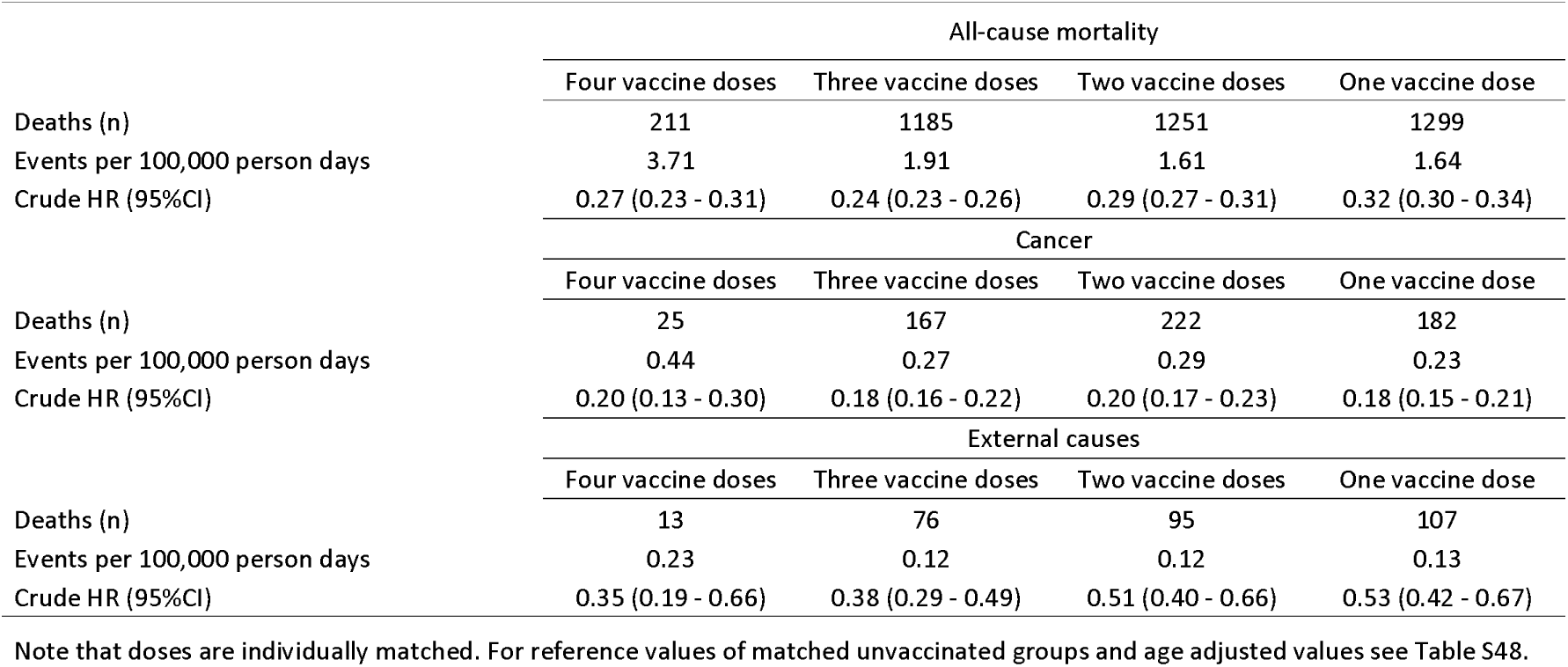
Hazard ratios (HR) with 95% confidence intervals (95% CI) for all-cause mortality, cancer deaths and deaths from external causes according to number of SARS-CoV-2 vaccine doses in the two weeks after vaccination with controls matched based on age group, sex, education and ALTCA status.

|  | All-cause mortality |  |  |  |
| --- | --- | --- | --- | --- |
|  | Four vaccine doses | Three vaccine doses | Two vaccine doses | One vaccine dose |
| Deaths (n) | 211 | 1185 | 1251 | 1299 |
| Events per 100,000 person days | 3.71 | 1.91 | 1.61 | 1.64 |
| Crude HR (95%CI) | 0.27 (0.23 - 0.31) | 0.24 (0.23 - 0.26) | 0.29 (0.27 - 0.31) | 0.32 (0.30 - 0.34) |
|  | Cancer |  |  |  |
|  | Four vaccine doses | Three vaccine doses | Two vaccine doses | One vaccine dose |
| Deaths (n) | 25 | 167 | 222 | 182 |
| Events per 100,000 person days | 0.44 | 0.27 | 0.29 | 0.23 |
| Crude HR (95%CI) | 0.20 (0.13 - 0.30) | 0.18 (0.16 - 0.22) | 0.20 (0.17 - 0.23) | 0.18 (0.15 - 0.21) |
|  | External causes |  |  |  |
|  | Four vaccine doses | Three vaccine doses | Two vaccine doses | One vaccine dose |
| Deaths (n) | 13 | 76 | 95 | 107 |
| Events per 100,000 person days | 0.23 | 0.12 | 0.12 | 0.13 |
| Crude HR (95%CI) | 0.35 (0.19 - 0.66) | 0.38 (0.29 - 0.49) | 0.51 (0.40 - 0.66) | 0.53 (0.42 - 0.67) |

Note that doses are individually matched. For reference values of matched unvaccinated groups and age adjusted values see Table S48.

### Analyses on negative control outcomes

Both cancer mortality and deaths by external cause mirror the same pattern of lower risk in vaccinated as all-cause mortality analyses (Table 1, 2 and 3).

## DISCUSSION

We observed significantly lower all-cause mortality in SARS-CoV-2 vaccinated groups versus the unvaccinated group in the Austrian population of individuals without a documented infection. This indicates the presence of a HVE as individuals with lower mortality risk were more likely to get vaccinated. Furthermore, negative control outcomes, i.e. death by cancer and external causes, show similar patterns as all-cause mortality. The bias was strongest immediately after each vaccination but appeared to wane thereafter.

Our results add to the growing number of studies showing different all-cause-mortality or non-COVID-19 in vaccinated versus unvaccinated populations.^3,9,17,27–33^ Reductions in all-cause-mortality in vaccinated versus unvaccinated individuals were particularly pronounced during and shortly after times of high vaccine rollout.^34^ The pattern of reduced all-cause mortality shortly after vaccinations indicates that good health status is associated with vaccination. This reflects that overall HVE outweighs confounding by indication once demographics and ALTCA are accounted for. However, the additional analysis of 3-month periods using preceding vaccination dose as reference shows that in all doses except the first, the earliest quarter of a doses’ introduction was not significantly different or even higher in risk of all-cause mortality than the preceding dose. This suggests that in the earliest periods when first introducing dose 2, 3 and 4, confounding by indication may play a relevant role, cancelling out HVEs. Among those who already had received doses, those who received the next dose very fast were in equally poor or worse health. However, HVE is a much stronger overall driver of biased VE estimates in COVID-19. Over time, dose specific HVE waned or even reversed as indicated by progressively increasing aHRs in all-cause mortality in vaccinated versus unvaccinated individuals. This may have contributed to the negative effectiveness (seemingly harmfulness) late after vaccination that can be seen in this and previous studies on both HVE and VE.^3,35^ Thus, HVE may contribute to overestimating VE in the short term while simultaneously leading to findings that wrongly suggest harmful long-term effects of vaccinations.

The analyses of periods of low and high COVID-19 disease burden further showed that the issue is unlikely driven by unidentified infections or misclassified deaths as high periods and low periods both experienced clear indications of HVE with magnitude seemingly being driven mainly by duration since rollout.

The matched mortality analyses of the first two weeks after vaccination also indicate a significantly reduced mortality in vaccinated versus unvaccinated individuals. These analyses may offer the most unbiased estimate of HVE, and they tend to show even larger HVE (aHR 0.24-0.32 for all-cause mortality) than the other analyses. In these two weeks, any VE against any cause of death is mechanistically implausible to have been substantial, so that these findings strongly support the presence of an HVE. The indicated difference in risk is substantial and present in all-cause mortality, deaths by cancer and external causes. The bias is strongest for cancer (which is a strong predictor for physical health) and less so for external causes, congruent with the fact that not all external death causes are associated with lower health status.

In a previous study we investigated HVE in individuals with a documented previous infection using the same methodological approach presented in this paper.^3^ Unlike the previous study, which had follow-up to the end of 2023, the current dataset only included infection and vaccination data up to September 2022, when data provision of person data to Statistics Austria stopped. Overall effects between the two investigations are therefore not directly comparable. However, the most important role-out phase of vaccinations 1 to 4 are covered and within the available period the findings presented closely match trends and even magnitude of previous findings. This is true for 3-month period analyses, analyses of high and low COVID-19 death count periods and also matched analyses. Other studies on HVE show how magnitude of the bias is heavily dependent on time-period and analysis setup (Table S49).^3,4,33^

Our study is limited by no access to data on comorbidities and measures of functional or socioeconomic status that may reflect worse health status and higher mortality. Therefore, we could not adjust for them to explore how much they might explain observed biases. However, previous investigations on HVE indicate only a minor impact of adjustments for other comorbidities or even an increase in HVE estimates by such additional adjustments.^10,15^ One recent study that used exact comorbidity matching found that HVE might have a much lower effect on VE than non-COVID mortality differences would suggest.^36^ However, these analyses are based on very few cases with a composite outcome (severe cases or deaths) in a country (Qatar) that may not be representative of most other countries. Thus, generalizability of these findings is not yet established. Our adjustments for demographics and ALTCA capture factors that were used for prioritization of vaccination and are thus important to consider in trying to get a “clean” estimate of HVE. Education, conversely, is a proxy for socioeconomic status and may thus be seen as a mediator rather than confounder for the HVE. Nevertheless, models with different sets of adjusting factors gave very similar results overall.

While we investigate the subpopulation with no documented infection, some of the people probably had already been infected and a substantial part of this population probably had an unrecorded infection within the observation period.^34^ Misclassification may have also led to both under- or overcounting.^37,38^ In other high-income countries, medical record audits suggest that overcounting is more likely ^38,39^ and consideration of multiple causes of deaths in the USA also suggest that COVID-19 deaths were overcounted.^40^ We partially address this ambiguity by investigating periods of low and high COVID-19 disease burden. These analyses indicate that the HVE cannot be explained solely by effects of unidentified SARS-CoV-2 infections, otherwise the effect would be much stronger in times of high versus low COVID-19 disease burden.

In conclusion, we found evidence of a substantial HVE among adults with no previously documented SARS-CoV-2 infection in Austria. This finding adds to a growing body of literature on HVE in COVID-19 vaccination studies, which remains largely overlooked in recent publications on vaccine effects on health outcomes.^6^ Future epidemiological studies should account for HVE when interpreting VE against death and other health outcomes.

## Data Availability

The data that support the findings of this study are available upon request with approval needed from Statistics Austria. The data are not publicly available due to restrictions pertaining to contained information that could compromise the privacy of patients.

## Contributors

UR and SP conceptualized the study with inputs by JPAI. UR and SP wrote the original draft. UR performed the formal analyses and visualization. All authors contributed to supervision, writing, reviewing, and editing the manuscript, and approved the final version before submission.

## Acknowledgements

The authors thank all persons and organizations involved in data collection.

