## Supplements for "Healthy vaccinee effect in SARS-CoV-2 vaccinees without documented infection"

### **Table S1:** STROBE Statement—Checklist of items for *cohort studies*

|  | **Item No** | **Recommendation** | **Main text page** |
| --- | --- | --- | --- |
| **Title and abstract** | 1 | (*a*) Indicate the study’s design with a commonly used term in the title or the abstract | 2 |
|  |  | (*b*) Provide in the abstract an informative and balanced summary of what was done and what was found | 2 & 3 |
| **Introduction** | | |  |
| Background/rationale | 2 | Explain the scientific background and rationale for the investigation being reported | 4 & 5 |
| Objectives | 3 | State specific objectives, including any prespecified hypotheses | 5 |
| **Methods** | | |  |
| Study design | 4 | Present key elements of study design early in the paper | 5-7 |
| Setting | 5 | Describe the setting, locations, and relevant dates, including periods of recruitment, exposure, follow-up, and data collection | 5 |
| Participants | 6 | (*a*) Give the eligibility criteria, and the sources and methods of selection of participants. Describe methods of follow-up | 6-8 |
|  |  | (*b*) For matched studies, give matching criteria and number of exposed and unexposed | 8 & 9 |
| Variables | 7 | Clearly define all outcomes, exposures, predictors, potential confounders, and effect modifiers. Give diagnostic criteria, if applicable | 5-8 |
| Data sources/ measurement | 8* | For each variable of interest, give sources of data and details of methods of assessment (measurement). Describe comparability of assessment methods if there is more than one group | 5 & 6 |
| Bias | 9 | Describe any efforts to address potential sources of bias | 7 & 8 |
| Study size | 10 | Explain how the study size was arrived at | 6 |
| Quantitative variables | 11 | Explain how quantitative variables were handled in the analyses. If applicable, describe which groupings were chosen and why | 5 & 6 |
| Statistical methods | 12 | (*a*) Describe all statistical methods, including those used to control for confounding | 6-8 |
|  |  | (*b*) Describe any methods used to examine subgroups and interactions | 7&8 |
|  |  | (*c*) Explain how missing data were addressed | 6-9 |
|  |  | (*d*) If applicable, explain how loss to follow-up was addressed | n/a |
|  |  | (*e*) Describe any sensitivity analyses | 7-8 |
| **Results** | | |  |
| Participants | 13* | (a) Report numbers of individuals at each stage of study—eg numbers potentially eligible, examined for eligibility, confirmed eligible, included in the study, completing follow-up, and analysed | 9-14 & Supplements |
|  |  | (b) Give reasons for non-participation at each stage | n/a |
|  |  | (c) Consider use of a flow diagram | 9 |
| Descriptive data | 14* | (a) Give characteristics of study participants (eg demographic, clinical, social) and information on exposures and potential confounders | Supplements |
|  |  | (b) Indicate number of participants with missing data for each variable of interest | Supplements |
|  |  | (c) Summarise follow-up time (eg, average and total amount) | Supplements |
| Outcome data | 15* | Report numbers of outcome events or summary measures over time | 9-14 & Supplements |
| Main results | 16 | (*a*) Give unadjusted estimates and, if applicable, confounder-adjusted estimates and their precision (eg, 95% confidence interval). Make clear which confounders were adjusted for and why they were included | 9-14 |
|  |  | (*b*) Report category boundaries when continuous variables were categorized | n/a |
|  |  | (*c*) If relevant, consider translating estimates of relative risk into absolute risk for a meaningful time period | n/a |
| Other analyses | 17 | Report other analyses done—eg analyses of subgroups and interactions, and sensitivity analyses | 14 |
| **Discussion** | | |  |
| Key results | 18 | Summarise key results with reference to study objectives | 15 |
| Limitations | 19 | Discuss limitations of the study, taking into account sources of potential bias or imprecision. Discuss both direction and magnitude of any potential bias | 17 & 18 |
| Interpretation | 20 | Give a cautious overall interpretation of results considering objectives, limitations, multiplicity of analyses, results from similar studies, and other relevant evidence | 15-18 |
| Generalisability | 21 | Discuss the generalisability (external validity) of the study results | 15-17 |
| **Other information** | | |  |
| Funding | 22 | Give the source of funding and the role of the funders for the present study and, if applicable, for the original study on which the present article is based | 18 |

*Give information separately for exposed and unexposed groups.

**Note:** An Explanation and Elaboration article discusses each checklist item and gives methodological background and published examples of transparent reporting. The STROBE checklist is best used in conjunction with this article (freely available on the Web sites of PLoS Medicine at http://www.plosmedicine.org/, Annals of Internal Medicine at http://www.annals.org/, and Epidemiology at http://www.epidem.com/). Information on the STROBE Initiative is available at <http://www.strobe-statement.org>.

| **Descriptive Tables (S2 – S8)**  **Table S2:** Characteristics of population for start of observation period January 1. 2021 | | | | | | | | |
| --- | --- | --- | --- | --- | --- | --- | --- | --- |
|  | Total | Female | Male | four doses | three doses | two doses | one dose | unvaccinated |
| N | 7,416,077 | 3,781,850 | 3,634,227 | 0 | 0 | 0 | 4,873 | 7,411,204 |
| Females | 3,781,850 (51.00%) | 3,781,850 (100.00%) | 0 (0.00%) | 0 (0%) | 0 (0%) | 0 (0%) | 3,147 (64.58%) | 3,778,703 (50.99%) |
| Age | 49 (33 - 63) | 50 (34 - 65) | 48 (33 - 61) | NA (NA - NA) | NA (NA - NA) | NA (NA - NA) | 61 (48 - 82) | 49 (33 - 63) |
| ALTCA recipients | 486,001 (6.55%) | 302,473 (8.00%) | 183,528 (5.05%) | 0 (0%) | 0 (0%) | 0 (0%) | 2,231 (45.78%) | 483,770 (6.53%) |
| Vaccinations against SARS-CoV-2: |  |  |  |  |  |  |  |  |
| Four doses | 0 (0.00%) | 0 (0.00%) | 0 (0.00%) | 0 (0%) | 0 (0%) | 0 (0%) | 0 (0.00%) | 0 (0.00%) |
| Three doses | 0 (0.00%) | 0 (0.00%) | 0 (0.00%) | 0 (0%) | 0 (0%) | 0 (0%) | 0 (0.00%) | 0 (0.00%) |
| Two doses | 0 (0.00%) | 0 (0.00%) | 0 (0.00%) | 0 (0%) | 0 (0%) | 0 (0%) | 0 (0.00%) | 0 (0.00%) |
| One dose | 4,873 (0.07%) | 3,147 (0.08%) | 1,726 (0.05%) | 0 (0%) | 0 (0%) | 0 (0%) | 4,873 (100.00%) | 0 (0.00%) |
| Unvaccinated | 0 (0.00%) | 0 (0.00%) | 0 (0.00%) | 0 (0%) | 0 (0%) | 0 (0%) | 0 (0.00%) | 0 (0.00%) |
| Time since last vaccination | 3 (2 - 4) | 3 (2 - 4) | 3 (2 - 4) | NA (NA - NA) | NA (NA - NA) | NA (NA - NA) | 3 (2 - 4) | NA (NA - NA) |
| Education: |  |  |  |  |  |  |  |  |
| Compulsory schooling or lower | 1,556,063 (20.98%) | 937,534 (24.79%) | 618,529 (17.02%) | 0 (0%) | 0 (0%) | 0 (0%) | 1,226 (25.16%) | 1,554,837 (20.98%) |
| Pre-secondary education | 3,372,610 (45.48%) | 1,531,739 (40.50%) | 1,840,871 (50.65%) | 0 (0%) | 0 (0%) | 0 (0%) | 2,024 (41.53%) | 3,370,586 (45.48%) |
| Secondary education | 1,110,331 (14.97%) | 570,762 (15.09%) | 539,569 (14.85%) | 0 (0%) | 0 (0%) | 0 (0%) | 488 (10.01%) | 1,109,843 (14.98%) |
| Pre-tertiary education | 190,504 (2.57%) | 136,434 (3.61%) | 54,070 (1.49%) | 0 (0%) | 0 (0%) | 0 (0%) | 138 (2.83%) | 190,366 (2.57%) |
| Tertiary education | 1,085,745 (14.64%) | 560,844 (14.83%) | 524,901 (14.44%) | 0 (0%) | 0 (0%) | 0 (0%) | 994 (20.40%) | 1,084,751 (14.64%) |
| Education unkown | 100,801 (1.36%) | 44,527 (1.18%) | 56,274 (1.55%) | 0 (0%) | 0 (0%) | 0 (0%) | 3 (0.06%) | 100,798 (1.36%) |
|  | ALTCA recipients | 85+ year old | 75-84 year old | 60-74 year old | 40-59 year old | 18-39 year old |  |  |
| N | 486,001 | 214,211 | 605,695 | 1,440,761 | 2,542,893 | 2,612,517 |  |  |
| Females | 302,473 (62.24%) | 142,267 (66.41%) | 346,366 (57.18%) | 756,612 (52.51%) | 1,266,988 (49.82%) | 1,269,617 (48.60%) |  |  |
| Age | 80 (69 - 86) | 88 (86 - 91) | 79 (77 - 81) | 66 (63 - 70) | 50 (45 - 55) | 29 (24 - 34) |  |  |
| ALTCA recipients | 486,001 (100.00%) | 151,592 (70.77%) | 163,620 (27.01%) | 99,449 (6.90%) | 49,306 (1.94%) | 22,034 (0.84%) |  |  |
| Vaccinations against SARS-CoV-2: |  |  |  |  |  |  |  |  |
| Four doses | 0 (0.00%) | 0 (0.00%) | 0 (0.00%) | 0 (0.00%) | 0 (0.00%) | 0 (0.00%) |  |  |
| Three doses | 0 (0.00%) | 0 (0.00%) | 0 (0.00%) | 0 (0.00%) | 0 (0.00%) | 0 (0.00%) |  |  |
| Two doses | 0 (0.00%) | 0 (0.00%) | 0 (0.00%) | 0 (0.00%) | 0 (0.00%) | 0 (0.00%) |  |  |
| One dose | 2,231 (0.46%) | 984 (0.46%) | 797 (0.13%) | 766 (0.05%) | 1,626 (0.06%) | 700 (0.03%) |  |  |
| Unvaccinated | 0 (0.00%) | 0 (0.00%) | 0 (0.00%) | 0 (0.00%) | 0 (0.00%) | 0 (0.00%) |  |  |
| Time since last vaccination | 3 (3 - 4) | 3 (3 - 4) | 3 (2 - 4) | 3 (2 - 4) | 3 (2 - 4) | 3 (2 - 4) |  |  |
| Education: |  |  |  |  |  |  |  |  |
| Compulsory schooling or lower | 233,228 (47.99%) | 109,586 (51.16%) | 215,559 (35.59%) | 341,697 (23.72%) | 441,338 (17.36%) | 447,883 (17.14%) |  |  |
| Pre-secondary education | 203,021 (41.77%) | 79,155 (36.95%) | 299,262 (49.41%) | 786,174 (54.57%) | 1,263,262 (49.68%) | 944,757 (36.16%) |  |  |
| Secondary education | 28,095 (5.78%) | 14,063 (6.57%) | 43,935 (7.25%) | 128,081 (8.89%) | 351,738 (13.83%) | 572,514 (21.91%) |  |  |
| Pre-tertiary education | 4,899 (1.01%) | 1,938 (0.90%) | 8,139 (1.34%) | 53,059 (3.68%) | 81,910 (3.22%) | 45,458 (1.74%) |  |  |
| Tertiary education | 16,495 (3.39%) | 9,053 (4.23%) | 37,490 (6.19%) | 125,130 (8.68%) | 380,131 (14.95%) | 533,941 (20.44%) |  |  |
| Education unkown | 263 (0.05%) | 416 (0.19%) | 1,310 (0.22%) | 6,620 (0.46%) | 24,512 (0.96%) | 67,943 (2.60%) |  |  |
|  | BioNTec/Pfizer | Moderna | AstraZeneca | Janssen (J&J) |  |  |  |  |
| N | 4,873 | 0 | 0 | 0 |  |  |  |  |
| Females | 3,147 (64.58%) | 0 (0%) | 0 (0%) | 0 (0%) |  |  |  |  |
| Age | 61 (48 - 82) | NA (NA - NA) | NA (NA - NA) | NA (NA - NA) |  |  |  |  |
| ALTCA recipients | 2,231 (45.78%) | 0 (0%) | 0 (0%) | 0 (0%) |  |  |  |  |
| Vaccinations against SARS-CoV-2: |  |  |  |  |  |  |  |  |
| Four doses | 0 (0.00%) | 0 (0%) | 0 (0%) | 0 (0%) |  |  |  |  |
| Three doses | 0 (0.00%) | 0 (0%) | 0 (0%) | 0 (0%) |  |  |  |  |
| Two doses | 0 (0.00%) | 0 (0%) | 0 (0%) | 0 (0%) |  |  |  |  |
| One dose | 4,873 (100.00%) | 0 (0%) | 0 (0%) | 0 (0%) |  |  |  |  |
| Unvaccinated | 0 (0.00%) | 0 (0%) | 0 (0%) | 0 (0%) |  |  |  |  |
| Time since last vaccination | 3 (2 - 4) | NA (NA - NA) | NA (NA - NA) | NA (NA - NA) |  |  |  |  |
| Education: |  |  |  |  |  |  |  |  |
| Compulsory schooling or lower | 1,226 (25.16%) | 0 (0%) | 0 (0%) | 0 (0%) |  |  |  |  |
| Pre-secondary education | 2,024 (41.53%) | 0 (0%) | 0 (0%) | 0 (0%) |  |  |  |  |
| Secondary education | 488 (10.01%) | 0 (0%) | 0 (0%) | 0 (0%) |  |  |  |  |
| Pre-tertiary education | 138 (2.83%) | 0 (0%) | 0 (0%) | 0 (0%) |  |  |  |  |
| Tertiary education | 994 (20.40%) | 0 (0%) | 0 (0%) | 0 (0%) |  |  |  |  |
| Education unkown | 3 (0.06%) | 0 (0%) | 0 (0%) | 0 (0%) |  |  |  |  |

| **Table S3:** Characteristics of population for end of observation period August 31. 2022 | | | | | | | | |
| --- | --- | --- | --- | --- | --- | --- | --- | --- |
|  | Total | Female | Male | four doses | three doses | two doses | one dose | unvaccinated |
| N | 4,133,426 | 2,067,111 | 2,066,315 | 373,458 | 2,533,699 | 471,744 | 49,647 | 704,878 |
| Females | 2,067,111 (50.01%) | 2,067,111 (100.00%) | 0 (0.00%) | 203,384 (54.46%) | 1,290,845 (50.95%) | 205,409 (43.54%) | 22,158 (44.63%) | 345,315 (48.99%) |
| Age | 54 (37 - 67) | 57 (39 - 70) | 52 (35 - 65) | 74 (65 - 81) | 56 (41 - 67) | 41 (29 - 56) | 39 (28 - 57) | 43 (29 - 59) |
| ALTCA recipients | 332,109 (8.03%) | 210,695 (10.19%) | 121,414 (5.88%) | 70,231 (18.81%) | 201,267 (7.94%) | 25,551 (5.42%) | 2,740 (5.52%) | 32,320 (4.59%) |
| Vaccinations against SARS-CoV-2: |  |  |  |  |  |  |  |  |
| Four doses | 373,458 (9.04%) | 203,384 (9.84%) | 170,074 (8.23%) | 373,458 (100.00%) | 0 (0.00%) | 0 (0.00%) | 0 (0.00%) | 0 (0.00%) |
| Three doses | 2,533,699 (61.30%) | 1,290,845 (62.45%) | 1,242,854 (60.15%) | 0 (0.00%) | 2,533,699 (100.00%) | 0 (0.00%) | 0 (0.00%) | 0 (0.00%) |
| Two doses | 471,744 (11.41%) | 205,409 (9.94%) | 266,335 (12.89%) | 0 (0.00%) | 0 (0.00%) | 471,744 (100.00%) | 0 (0.00%) | 0 (0.00%) |
| One dose | 49,647 (1.20%) | 22,158 (1.07%) | 27,489 (1.33%) | 0 (0.00%) | 0 (0.00%) | 0 (0.00%) | 49,647 (100.00%) | 0 (0.00%) |
| Unvaccinated | 0 (0.00%) | 0 (0.00%) | 0 (0.00%) | 0 (0.00%) | 0 (0.00%) | 0 (0.00%) | 0 (0.00%) | 0 (0.00%) |
| Time since last vaccination | 261 (222 - 279) | 263 (222 - 280) | 260 (223 - 278) | 44 (26 - 60) | 265 (236 - 280) | 267 (241 - 295) | 270 (225 - 341) | NA (NA - NA) |
| Education: |  |  |  |  |  |  |  |  |
| Compulsory schooling or lower | 960,759 (23.24%) | 578,278 (27.98%) | 382,481 (18.51%) | 77,522 (20.76%) | 529,214 (20.89%) | 158,126 (33.52%) | 18,394 (37.05%) | 177,503 (25.18%) |
| Pre-secondary education | 1,896,298 (45.88%) | 844,380 (40.85%) | 1,051,918 (50.91%) | 179,238 (47.99%) | 1,236,389 (48.80%) | 206,058 (43.68%) | 16,355 (32.94%) | 258,258 (36.64%) |
| Secondary education | 556,507 (13.46%) | 273,769 (13.24%) | 282,738 (13.68%) | 46,206 (12.37%) | 354,986 (14.01%) | 60,272 (12.78%) | 6,518 (13.13%) | 88,525 (12.56%) |
| Pre-tertiary education | 99,051 (2.40%) | 68,535 (3.32%) | 30,516 (1.48%) | 13,683 (3.66%) | 62,103 (2.45%) | 5,747 (1.22%) | 879 (1.77%) | 16,639 (2.36%) |
| Tertiary education | 529,912 (12.82%) | 261,680 (12.66%) | 268,232 (12.98%) | 56,762 (15.20%) | 349,465 (13.79%) | 39,292 (8.33%) | 6,163 (12.41%) | 78,230 (11.10%) |
| Education unkown | 90,877 (2.20%) | 40,459 (1.96%) | 50,418 (2.44%) | 47 (0.01%) | 1,542 (0.06%) | 2,248 (0.48%) | 1,337 (2.69%) | 85,703 (12.16%) |
|  | ALTCA recipients | 85+ year old | 75-84 year old | 60-74 year old | 40-59 year old | 18-39 year old |  |  |
| N | 332,109 | 150,227 | 458,307 | 1,029,060 | 1,330,618 | 1,165,214 |  |  |
| Females | 210,695 (63.44%) | 99,689 (66.36%) | 268,408 (58.57%) | 545,119 (52.97%) | 631,690 (47.47%) | 522,205 (44.82%) |  |  |
| Age | 80 (69 - 86) | 88 (86 - 91) | 79 (77 - 81) | 66 (63 - 70) | 51 (46 - 55) | 29 (24 - 34) |  |  |
| ALTCA recipients | 332,109 (100.00%) | 99,439 (66.19%) | 118,130 (25.78%) | 70,215 (6.82%) | 32,133 (2.41%) | 12,192 (1.05%) |  |  |
| Vaccinations against SARS-CoV-2: |  |  |  |  |  |  |  |  |
| Four doses | 70,231 (21.15%) | 44,645 (29.72%) | 133,104 (29.04%) | 142,878 (13.88%) | 41,877 (3.15%) | 10,954 (0.94%) |  |  |
| Three doses | 201,267 (60.60%) | 82,415 (54.86%) | 267,113 (58.28%) | 688,705 (66.93%) | 907,641 (68.21%) | 587,825 (50.45%) |  |  |
| Two doses | 25,551 (7.69%) | 8,972 (5.97%) | 20,774 (4.53%) | 65,335 (6.35%) | 152,225 (11.44%) | 224,438 (19.26%) |  |  |
| One dose | 2,740 (0.83%) | 954 (0.64%) | 2,224 (0.49%) | 7,527 (0.73%) | 13,790 (1.04%) | 25,152 (2.16%) |  |  |
| Unvaccinated | 0 (0.00%) | 0 (0.00%) | 0 (0.00%) | 0 (0.00%) | 0 (0.00%) | 0 (0.00%) |  |  |
| Time since last vaccination | 265 (112 - 288) | 267 (63 - 297) | 265 (63 - 287) | 265 (224 - 280) | 263 (232 - 278) | 255 (222 - 274) |  |  |
| Education: |  |  |  |  |  |  |  |  |
| Compulsory schooling or lower | 154,143 (46.41%) | 74,486 (49.58%) | 156,297 (34.10%) | 244,877 (23.80%) | 251,524 (18.90%) | 233,575 (20.05%) |  |  |
| Pre-secondary education | 142,822 (43.00%) | 57,264 (38.12%) | 230,829 (50.37%) | 561,090 (54.52%) | 651,134 (48.93%) | 395,981 (33.98%) |  |  |
| Secondary education | 19,741 (5.94%) | 9,993 (6.65%) | 34,526 (7.53%) | 92,373 (8.98%) | 179,206 (13.47%) | 240,409 (20.63%) |  |  |
| Pre-tertiary education | 3,564 (1.07%) | 1,431 (0.95%) | 6,656 (1.45%) | 36,423 (3.54%) | 36,241 (2.72%) | 18,300 (1.57%) |  |  |
| Tertiary education | 11,598 (3.49%) | 6,692 (4.45%) | 28,787 (6.28%) | 88,123 (8.56%) | 190,216 (14.30%) | 216,094 (18.55%) |  |  |
| Education unkown | 241 (0.07%) | 361 (0.24%) | 1,212 (0.26%) | 6,174 (0.60%) | 22,295 (1.68%) | 60,835 (5.22%) |  |  |
|  | BioNTec/Pfizer | Moderna | AstraZeneca | Janssen (J&J) |  |  |  |  |
| N | 3,093,423 | 308,817 | 5,742 | 15,213 |  |  |  |  |
| Females | 1,564,930 (50.59%) | 145,603 (47.15%) | 2,856 (49.74%) | 5,564 (36.57%) |  |  |  |  |
| Age | 56 (39 - 69) | 56 (45 - 66) | 51 (32 - 66) | 42 (29 - 56) |  |  |  |  |
| ALTCA recipients | 277,085 (8.96%) | 20,960 (6.79%) | 589 (10.26%) | 665 (4.37%) |  |  |  |  |
| Vaccinations against SARS-CoV-2: |  |  |  |  |  |  |  |  |
| Four doses | 351,345 (11.36%) | 20,761 (6.72%) | 16 (0.28%) | 20 (0.13%) |  |  |  |  |
| Three doses | 2,275,975 (73.57%) | 254,874 (82.53%) | 392 (6.83%) | 748 (4.92%) |  |  |  |  |
| Two doses | 429,286 (13.88%) | 30,706 (9.94%) | 4,709 (82.01%) | 5,013 (32.95%) |  |  |  |  |
| One dose | 36,817 (1.19%) | 2,476 (0.80%) | 625 (10.88%) | 9,432 (62.00%) |  |  |  |  |
| Unvaccinated | 0 (0.00%) | 0 (0.00%) | 0 (0.00%) | 0 (0.00%) |  |  |  |  |
| Time since last vaccination | 260 (221 - 279) | 264 (236 - 278) | 423 (385 - 452) | 330 (264 - 390) |  |  |  |  |
| Education: |  |  |  |  |  |  |  |  |
| Compulsory schooling or lower | 717,390 (23.19%) | 58,550 (18.96%) | 1,516 (26.40%) | 4,933 (32.43%) |  |  |  |  |
| Pre-secondary education | 1,477,618 (47.77%) | 150,111 (48.61%) | 2,067 (36.00%) | 5,792 (38.07%) |  |  |  |  |
| Secondary education | 420,764 (13.60%) | 43,438 (14.07%) | 825 (14.37%) | 2,113 (13.89%) |  |  |  |  |
| Pre-tertiary education | 73,839 (2.39%) | 8,032 (2.60%) | 150 (2.61%) | 233 (1.53%) |  |  |  |  |
| Tertiary education | 399,460 (12.91%) | 48,283 (15.63%) | 1,103 (19.21%) | 1,811 (11.90%) |  |  |  |  |
| Education unkown | 4,350 (0.14%) | 403 (0.13%) | 81 (1.41%) | 331 (2.18%) |  |  |  |  |

| **Table S4:** Characteristics of population for 2021 low COVID-19 disease burden on June 1. 2021 | | | | | | | | |
| --- | --- | --- | --- | --- | --- | --- | --- | --- |
|  | Total | Female | Male | four doses | three doses | two doses | one dose | unvaccinated |
| N | 6,873,426 | 3,518,737 | 3,354,689 | 0 | 10 | 1,089,615 | 1,869,734 | 3,914,067 |
| Females | 3,518,737 (51.19%) | 3,518,737 (100.00%) | 0 (0.00%) | 0 (0%) | 1 (10.00%) | 627,251 (57.57%) | 965,118 (51.62%) | 1,926,367 (49.22%) |
| Age | 50 (34 - 64) | 51 (35 - 65) | 49 (34 - 62) | NA (NA - NA) | 62.5 (57.5 - 68.5) | 71 (53 - 81) | 58 (46 - 67) | 41 (30 - 54) |
| ALTCA recipients | 449,155 (6.53%) | 280,672 (7.98%) | 168,483 (5.02%) | 0 (0%) | 2 (20.00%) | 224,020 (20.56%) | 91,259 (4.88%) | 133,874 (3.42%) |
| Vaccinations against SARS-CoV-2: |  |  |  |  |  |  |  |  |
| Four doses | 0 (0.00%) | 0 (0.00%) | 0 (0.00%) | 0 (0%) | 0 (0.00%) | 0 (0.00%) | 0 (0.00%) | 0 (0.00%) |
| Three doses | 10 (0.00%) | 1 (0.00%) | 9 (0.00%) | 0 (0%) | 10 (100.00%) | 0 (0.00%) | 0 (0.00%) | 0 (0.00%) |
| Two doses | 1,089,615 (15.85%) | 627,251 (17.83%) | 462,364 (13.78%) | 0 (0%) | 0 (0.00%) | 1,089,615 (100.00%) | 0 (0.00%) | 0 (0.00%) |
| One dose | 1,869,734 (27.20%) | 965,118 (27.43%) | 904,616 (26.97%) | 0 (0%) | 0 (0.00%) | 0 (0.00%) | 1,869,734 (100.00%) | 0 (0.00%) |
| Unvaccinated | 0 (0.00%) | 0 (0.00%) | 0 (0.00%) | 0 (0%) | 0 (0.00%) | 0 (0.00%) | 0 (0.00%) | 0 (0.00%) |
| Time since last vaccination | 32 (20 - 55) | 33 (20 - 60) | 32 (20 - 52) | NA (NA - NA) | 21 (18.25 - 21.75) | 53 (32 - 77) | 26 (15 - 39) | NA (NA - NA) |
| Education: |  |  |  |  |  |  |  |  |
| Compulsory schooling or lower | 1,436,204 (20.90%) | 874,505 (24.85%) | 561,699 (16.74%) | 0 (0%) | 1 (10.00%) | 263,313 (24.17%) | 296,557 (15.86%) | 876,333 (22.39%) |
| Pre-secondary education | 3,181,932 (46.29%) | 1,446,987 (41.12%) | 1,734,945 (51.72%) | 0 (0%) | 4 (40.00%) | 504,778 (46.33%) | 896,917 (47.97%) | 1,780,233 (45.48%) |
| Secondary education | 1,046,100 (15.22%) | 537,785 (15.28%) | 508,315 (15.15%) | 0 (0%) | 0 (0.00%) | 124,572 (11.43%) | 269,164 (14.40%) | 652,364 (16.67%) |
| Pre-tertiary education | 175,995 (2.56%) | 127,763 (3.63%) | 48,232 (1.44%) | 0 (0%) | 0 (0.00%) | 33,999 (3.12%) | 75,635 (4.05%) | 66,361 (1.70%) |
| Tertiary education | 1,018,102 (14.81%) | 525,669 (14.94%) | 492,433 (14.68%) | 0 (0%) | 5 (50.00%) | 162,909 (14.95%) | 331,318 (17.72%) | 523,870 (13.38%) |
| Education unkown | 15,088 (0.22%) | 6,026 (0.17%) | 9,062 (0.27%) | 0 (0%) | 0 (0.00%) | 44 (0.00%) | 143 (0.01%) | 14,901 (0.38%) |
|  | ALTCA recipients | 85+ year old | 75-84 year old | 60-74 year old | 40-59 year old | 18-39 year old |  |  |
| N | 449,155 | 209,820 | 590,921 | 1,414,578 | 2,370,100 | 2,288,007 |  |  |
| Females | 280,672 (62.49%) | 138,734 (66.12%) | 337,915 (57.18%) | 743,080 (52.53%) | 1,182,466 (49.89%) | 1,116,542 (48.80%) |  |  |
| Age | 80 (69 - 86) | 88 (86 - 91) | 79 (77 - 81) | 66 (63 - 70) | 50 (45 - 55) | 29 (24 - 34) |  |  |
| ALTCA recipients | 449,155 (100.00%) | 142,699 (68.01%) | 148,252 (25.09%) | 91,528 (6.47%) | 45,569 (1.92%) | 21,107 (0.92%) |  |  |
| Vaccinations against SARS-CoV-2: |  |  |  |  |  |  |  |  |
| Four doses | 0 (0.00%) | 0 (0.00%) | 0 (0.00%) | 0 (0.00%) | 0 (0.00%) | 0 (0.00%) |  |  |
| Three doses | 2 (0.00%) | 1 (0.00%) | 1 (0.00%) | 4 (0.00%) | 3 (0.00%) | 1 (0.00%) |  |  |
| Two doses | 224,020 (49.88%) | 141,652 (67.51%) | 321,633 (54.43%) | 258,713 (18.29%) | 232,590 (9.81%) | 135,027 (5.90%) |  |  |
| One dose | 91,259 (20.32%) | 18,705 (8.91%) | 155,036 (26.24%) | 686,633 (48.54%) | 684,715 (28.89%) | 324,645 (14.19%) |  |  |
| Unvaccinated | 0 (0.00%) | 0 (0.00%) | 0 (0.00%) | 0 (0.00%) | 0 (0.00%) | 0 (0.00%) |  |  |
| Time since last vaccination | 57 (33 - 75) | 69 (59 - 82) | 49 (27 - 63) | 32 (19 - 46) | 26 (18 - 48) | 27 (18 - 53) |  |  |
| Education: |  |  |  |  |  |  |  |  |
| Compulsory schooling or lower | 215,705 (48.02%) | 106,781 (50.89%) | 206,798 (35.00%) | 330,541 (23.37%) | 401,358 (16.93%) | 390,726 (17.08%) |  |  |
| Pre-secondary education | 187,606 (41.77%) | 78,397 (37.36%) | 295,011 (49.92%) | 777,486 (54.96%) | 1,185,117 (50.00%) | 845,921 (36.97%) |  |  |
| Secondary education | 25,996 (5.79%) | 13,640 (6.50%) | 43,588 (7.38%) | 127,565 (9.02%) | 335,515 (14.16%) | 525,792 (22.98%) |  |  |
| Pre-tertiary education | 4,572 (1.02%) | 1,894 (0.90%) | 8,081 (1.37%) | 52,539 (3.71%) | 76,539 (3.23%) | 36,942 (1.61%) |  |  |
| Tertiary education | 15,217 (3.39%) | 8,974 (4.28%) | 37,224 (6.30%) | 125,593 (8.88%) | 367,927 (15.52%) | 478,384 (20.91%) |  |  |
| Education unkown | 59 (0.01%) | 134 (0.06%) | 219 (0.04%) | 854 (0.06%) | 3,644 (0.15%) | 10,237 (0.45%) |  |  |
|  | BioNTec/Pfizer | Moderna | AstraZeneca | Janssen (J&J) |  |  |  |  |
| N | 1,939,956 | 273,908 | 729,085 | 16,410 |  |  |  |  |
| Females | 1,048,774 (54.06%) | 138,990 (50.74%) | 398,331 (54.63%) | 6,275 (38.24%) |  |  |  |  |
| Age | 63 (51 - 76) | 61 (49 - 72) | 56 (41 - 68) | 47 (34 - 57) |  |  |  |  |
| ALTCA recipients | 246,506 (12.71%) | 33,972 (12.40%) | 33,744 (4.63%) | 1,059 (6.45%) |  |  |  |  |
| Vaccinations against SARS-CoV-2: |  |  |  |  |  |  |  |  |
| Four doses | 0 (0.00%) | 0 (0.00%) | 0 (0.00%) | 0 (0.00%) |  |  |  |  |
| Three doses | 5 (0.00%) | 2 (0.00%) | 1 (0.00%) | 2 (0.01%) |  |  |  |  |
| Two doses | 873,929 (45.05%) | 143,484 (52.38%) | 72,192 (9.90%) | 10 (0.06%) |  |  |  |  |
| One dose | 1,066,022 (54.95%) | 130,422 (47.62%) | 656,892 (90.10%) | 16,398 (99.93%) |  |  |  |  |
| Unvaccinated | 0 (0.00%) | 0 (0.00%) | 0 (0.00%) | 0 (0.00%) |  |  |  |  |
| Time since last vaccination | 32 (20 - 55) | 27 (18 - 41) | 45 (20 - 60) | 20 (16 - 26) |  |  |  |  |
| Education: |  |  |  |  |  |  |  |  |
| Compulsory schooling or lower | 406,298 (20.94%) | 53,763 (19.63%) | 97,520 (13.38%) | 2,290 (13.95%) |  |  |  |  |
| Pre-secondary education | 943,783 (48.65%) | 133,457 (48.72%) | 316,156 (43.36%) | 8,303 (50.60%) |  |  |  |  |
| Secondary education | 247,630 (12.76%) | 37,642 (13.74%) | 105,477 (14.47%) | 2,987 (18.20%) |  |  |  |  |
| Pre-tertiary education | 56,589 (2.92%) | 8,092 (2.95%) | 44,693 (6.13%) | 260 (1.58%) |  |  |  |  |
| Tertiary education | 285,544 (14.72%) | 40,939 (14.95%) | 165,182 (22.66%) | 2,567 (15.64%) |  |  |  |  |
| Education unkown | 112 (0.01%) | 15 (0.01%) | 57 (0.01%) | 3 (0.02%) |  |  |  |  |

| **Table S5:** Characteristics of population for 2022 low COVID-19 disease burden on May 1. 2022 | | | | | | | | |
| --- | --- | --- | --- | --- | --- | --- | --- | --- |
|  | Total | Female | Male | four doses | three doses | two doses | one dose | unvaccinated |
| N | 4,548,251 | 2,303,395 | 2,244,856 | 8,492 | 3,252,167 | 633,840 | 52,455 | 601,297 |
| Females | 2,303,395 (50.64%) | 2,303,395 (100.00%) | 0 (0.00%) | 4,165 (49.05%) | 1,680,041 (51.66%) | 288,187 (45.47%) | 23,811 (45.39%) | 307,191 (51.09%) |
| Age | 54 (38 - 68) | 56 (39 - 70) | 53 (36 - 65) | 71 (59 - 80) | 57 (42 - 70) | 42 (29 - 57) | 40 (28 - 58) | 48 (33 - 63) |
| ALTCA recipients | 371,843 (8.18%) | 233,930 (10.16%) | 137,913 (6.14%) | 1,747 (20.57%) | 291,245 (8.96%) | 39,098 (6.17%) | 3,375 (6.43%) | 36,378 (6.05%) |
| Vaccinations against SARS-CoV-2: |  |  |  |  |  |  |  |  |
| Four doses | 8,492 (0.19%) | 4,165 (0.18%) | 4,327 (0.19%) | 8,492 (100.00%) | 0 (0.00%) | 0 (0.00%) | 0 (0.00%) | 0 (0.00%) |
| Three doses | 3,252,167 (71.50%) | 1,680,041 (72.94%) | 1,572,126 (70.03%) | 0 (0.00%) | 3,252,167 (100.00%) | 0 (0.00%) | 0 (0.00%) | 0 (0.00%) |
| Two doses | 633,840 (13.94%) | 288,187 (12.51%) | 345,653 (15.40%) | 0 (0.00%) | 0 (0.00%) | 633,840 (100.00%) | 0 (0.00%) | 0 (0.00%) |
| One dose | 52,455 (1.15%) | 23,811 (1.03%) | 28,644 (1.28%) | 0 (0.00%) | 0 (0.00%) | 0 (0.00%) | 52,455 (100.00%) | 0 (0.00%) |
| Unvaccinated | 0 (0.00%) | 0 (0.00%) | 0 (0.00%) | 0 (0.00%) | 0 (0.00%) | 0 (0.00%) | 0 (0.00%) | 0 (0.00%) |
| Time since last vaccination | 148 (125 - 163) | 149 (130 - 164) | 146 (123 - 161) | 44 (26 - 93) | 148 (129 - 162) | 145 (118 - 170) | 148 (104 - 215) | NA (NA - NA) |
| Education: |  |  |  |  |  |  |  |  |
| Compulsory schooling or lower | 1,043,164 (22.94%) | 631,400 (27.41%) | 411,764 (18.34%) | 1,144 (13.47%) | 649,646 (19.98%) | 204,158 (32.21%) | 19,831 (37.81%) | 168,385 (28.00%) |
| Pre-secondary education | 2,092,059 (46.00%) | 942,776 (40.93%) | 1,149,283 (51.20%) | 3,118 (36.72%) | 1,540,987 (47.38%) | 281,027 (44.34%) | 17,613 (33.58%) | 249,314 (41.46%) |
| Secondary education | 648,931 (14.27%) | 323,001 (14.02%) | 325,930 (14.52%) | 1,164 (13.71%) | 474,217 (14.58%) | 84,606 (13.35%) | 7,083 (13.50%) | 81,861 (13.61%) |
| Pre-tertiary education | 112,258 (2.47%) | 79,495 (3.45%) | 32,763 (1.46%) | 314 (3.70%) | 89,196 (2.74%) | 8,005 (1.26%) | 853 (1.63%) | 13,890 (2.31%) |
| Tertiary education | 633,583 (13.93%) | 317,226 (13.77%) | 316,357 (14.09%) | 2,750 (32.38%) | 497,697 (15.30%) | 55,259 (8.72%) | 6,573 (12.53%) | 71,304 (11.86%) |
| Education unkown | 18,250 (0.40%) | 9,495 (0.41%) | 8,755 (0.39%) | 2 (0.02%) | 424 (0.01%) | 784 (0.12%) | 501 (0.96%) | 16,539 (2.75%) |
|  | ALTCA recipients | 85+ year old | 75-84 year old | 60-74 year old | 40-59 year old | 18-39 year old |  |  |
| N | 371,843 | 176,357 | 509,825 | 1,134,454 | 1,475,376 | 1,252,239 |  |  |
| Females | 233,930 (62.91%) | 116,304 (65.95%) | 295,541 (57.97%) | 600,003 (52.89%) | 713,851 (48.38%) | 577,696 (46.13%) |  |  |
| Age | 80 (70 - 86) | 88 (86 - 91) | 79 (77 - 82) | 66 (63 - 70) | 51 (46 - 55) | 30 (24 - 34) |  |  |
| ALTCA recipients | 371,843 (100.00%) | 117,412 (66.58%) | 129,511 (25.40%) | 76,300 (6.73%) | 34,428 (2.33%) | 14,192 (1.13%) |  |  |
| Vaccinations against SARS-CoV-2: |  |  |  |  |  |  |  |  |
| Four doses | 1,747 (0.47%) | 984 (0.56%) | 2,569 (0.50%) | 2,801 (0.25%) | 1,662 (0.11%) | 476 (0.04%) |  |  |
| Three doses | 291,245 (78.32%) | 141,446 (80.20%) | 433,605 (85.05%) | 902,126 (79.52%) | 1,063,075 (72.05%) | 711,915 (56.85%) |  |  |
| Two doses | 39,098 (10.51%) | 14,814 (8.40%) | 30,962 (6.07%) | 91,203 (8.04%) | 202,659 (13.74%) | 294,202 (23.49%) |  |  |
| One dose | 3,375 (0.91%) | 1,295 (0.73%) | 2,641 (0.52%) | 8,174 (0.72%) | 14,425 (0.98%) | 25,920 (2.07%) |  |  |
| Unvaccinated | 0 (0.00%) | 0 (0.00%) | 0 (0.00%) | 0 (0.00%) | 0 (0.00%) | 0 (0.00%) |  |  |
| Time since last vaccination | 160 (140 - 180) | 172 (152 - 199) | 163 (149 - 176) | 151 (136 - 163) | 143 (122 - 158) | 135 (106 - 154) |  |  |
| Education: |  |  |  |  |  |  |  |  |
| Compulsory schooling or lower | 171,935 (46.24%) | 87,648 (49.70%) | 169,823 (33.31%) | 262,829 (23.17%) | 265,800 (18.02%) | 257,064 (20.53%) |  |  |
| Pre-secondary education | 160,107 (43.06%) | 67,348 (38.19%) | 259,755 (50.95%) | 619,599 (54.62%) | 718,588 (48.71%) | 426,769 (34.08%) |  |  |
| Secondary education | 22,374 (6.02%) | 11,633 (6.60%) | 39,252 (7.70%) | 105,658 (9.31%) | 209,694 (14.21%) | 282,694 (22.58%) |  |  |
| Pre-tertiary education | 4,035 (1.09%) | 1,647 (0.93%) | 7,603 (1.49%) | 41,982 (3.70%) | 42,683 (2.89%) | 18,343 (1.46%) |  |  |
| Tertiary education | 13,343 (3.59%) | 7,922 (4.49%) | 33,121 (6.50%) | 102,945 (9.07%) | 233,869 (15.85%) | 255,726 (20.42%) |  |  |
| Education unkown | 49 (0.01%) | 159 (0.09%) | 271 (0.05%) | 1,441 (0.13%) | 4,741 (0.32%) | 11,638 (0.93%) |  |  |
|  | BioNTec/Pfizer | Moderna | AstraZeneca | Janssen (J&J) |  |  |  |  |
| N | 3,546,888 | 373,131 | 6,813 | 17,039 |  |  |  |  |
| Females | 1,806,557 (50.93%) | 178,047 (47.72%) | 3,440 (50.49%) | 6,544 (38.41%) |  |  |  |  |
| Age | 55 (38 - 69) | 56 (45 - 66) | 53 (34 - 69) | 44 (30 - 57) |  |  |  |  |
| ALTCA recipients | 307,323 (8.66%) | 26,330 (7.06%) | 773 (11.35%) | 887 (5.21%) |  |  |  |  |
| Vaccinations against SARS-CoV-2: |  |  |  |  |  |  |  |  |
| Four doses | 7,536 (0.21%) | 849 (0.23%) | 5 (0.07%) | 10 (0.06%) |  |  |  |  |
| Three doses | 2,922,855 (82.41%) | 327,644 (87.81%) | 569 (8.35%) | 529 (3.10%) |  |  |  |  |
| Two doses | 577,492 (16.28%) | 42,006 (11.26%) | 5,586 (81.99%) | 6,911 (40.56%) |  |  |  |  |
| One dose | 39,005 (1.10%) | 2,632 (0.71%) | 653 (9.58%) | 9,589 (56.28%) |  |  |  |  |
| Unvaccinated | 0 (0.00%) | 0 (0.00%) | 0 (0.00%) | 0 (0.00%) |  |  |  |  |
| Time since last vaccination | 148 (124 - 163) | 146 (130 - 158) | 298 (248 - 328) | 186 (139 - 260) |  |  |  |  |
| Education: |  |  |  |  |  |  |  |  |
| Compulsory schooling or lower | 799,071 (22.53%) | 68,107 (18.25%) | 1,747 (25.64%) | 5,419 (31.80%) |  |  |  |  |
| Pre-secondary education | 1,654,037 (46.63%) | 178,092 (47.73%) | 2,508 (36.81%) | 6,760 (39.67%) |  |  |  |  |
| Secondary education | 508,763 (14.34%) | 54,306 (14.55%) | 1,003 (14.72%) | 2,421 (14.21%) |  |  |  |  |
| Pre-tertiary education | 87,580 (2.47%) | 10,251 (2.75%) | 188 (2.76%) | 260 (1.53%) |  |  |  |  |
| Tertiary education | 495,927 (13.98%) | 62,249 (16.68%) | 1,350 (19.82%) | 2,122 (12.45%) |  |  |  |  |
| Education unkown | 1,508 (0.04%) | 126 (0.03%) | 17 (0.25%) | 57 (0.33%) |  |  |  |  |

| **Table S6:** Characteristics of population for 2021 high COVID-19 disease burden on October 1. 2021 | | | | | | | | |
| --- | --- | --- | --- | --- | --- | --- | --- | --- |
|  | Total | Female | Male | four doses | three doses | two doses | one dose | unvaccinated |
| N | 6,822,547 | 3,491,282 | 3,331,265 | 0 | 22,634 | 4,616,329 | 388,276 | 1,795,308 |
| Females | 3,491,282 (51.17%) | 3,491,282 (100.00%) | 0 (0.00%) | 0 (0%) | 12,771 (56.42%) | 2,395,449 (51.89%) | 171,230 (44.10%) | 911,832 (50.79%) |
| Age | 50 (35 - 64) | 51 (35 - 66) | 49 (34 - 62) | NA (NA - NA) | 72 (55 - 82) | 54 (38 - 67) | 39 (28 - 54) | 42 (30 - 57) |
| ALTCA recipients | 429,806 (6.30%) | 269,739 (7.73%) | 160,067 (4.80%) | 0 (0%) | 7,445 (32.89%) | 323,588 (7.01%) | 16,617 (4.28%) | 82,156 (4.58%) |
| Vaccinations against SARS-CoV-2: |  |  |  |  |  |  |  |  |
| Four doses | 0 (0.00%) | 0 (0.00%) | 0 (0.00%) | 0 (0%) | 0 (0.00%) | 0 (0.00%) | 0 (0.00%) | 0 (0.00%) |
| Three doses | 22,634 (0.33%) | 12,771 (0.37%) | 9,863 (0.30%) | 0 (0%) | 22,634 (100.00%) | 0 (0.00%) | 0 (0.00%) | 0 (0.00%) |
| Two doses | 4,616,329 (67.66%) | 2,395,449 (68.61%) | 2,220,880 (66.67%) | 0 (0%) | 0 (0.00%) | 4,616,329 (100.00%) | 0 (0.00%) | 0 (0.00%) |
| One dose | 388,276 (5.69%) | 171,230 (4.90%) | 217,046 (6.52%) | 0 (0%) | 0 (0.00%) | 0 (0.00%) | 388,276 (100.00%) | 0 (0.00%) |
| Unvaccinated | 0 (0.00%) | 0 (0.00%) | 0 (0.00%) | 0 (0%) | 0 (0.00%) | 0 (0.00%) | 0 (0.00%) | 0 (0.00%) |
| Time since last vaccination | 100 (77 - 127) | 103 (78 - 133) | 98 (75 - 124) | NA (NA - NA) | 20 (16 - 27) | 101 (79 - 129) | 71 (22 - 108) | NA (NA - NA) |
| Education: |  |  |  |  |  |  |  |  |
| Compulsory schooling or lower | 1,426,440 (20.91%) | 864,849 (24.77%) | 561,591 (16.86%) | 0 (0%) | 5,088 (22.48%) | 849,925 (18.41%) | 99,706 (25.68%) | 471,721 (26.28%) |
| Pre-secondary education | 3,154,590 (46.24%) | 1,435,502 (41.12%) | 1,719,088 (51.60%) | 0 (0%) | 9,132 (40.35%) | 2,123,575 (46.00%) | 169,888 (43.75%) | 851,995 (47.46%) |
| Secondary education | 1,044,997 (15.32%) | 537,767 (15.40%) | 507,230 (15.23%) | 0 (0%) | 2,471 (10.92%) | 722,642 (15.65%) | 66,151 (17.04%) | 253,733 (14.13%) |
| Pre-tertiary education | 174,853 (2.56%) | 126,789 (3.63%) | 48,064 (1.44%) | 0 (0%) | 643 (2.84%) | 135,129 (2.93%) | 5,840 (1.50%) | 33,241 (1.85%) |
| Tertiary education | 1,012,374 (14.84%) | 522,510 (14.97%) | 489,864 (14.71%) | 0 (0%) | 5,300 (23.42%) | 784,759 (17.00%) | 46,529 (11.98%) | 175,786 (9.79%) |
| Education unkown | 9,285 (0.14%) | 3,861 (0.11%) | 5,424 (0.16%) | 0 (0%) | 0 (0.00%) | 299 (0.01%) | 162 (0.04%) | 8,824 (0.49%) |
|  | ALTCA recipients | 85+ year old | 75-84 year old | 60-74 year old | 40-59 year old | 18-39 year old |  |  |
| N | 429,806 | 210,284 | 594,632 | 1,419,436 | 2,348,000 | 2,250,195 |  |  |
| Females | 269,739 (62.76%) | 138,664 (65.94%) | 339,962 (57.17%) | 745,398 (52.51%) | 1,170,936 (49.87%) | 1,096,322 (48.72%) |  |  |
| Age | 80 (69 - 86) | 88 (86 - 91) | 79 (77 - 81) | 66 (63 - 70) | 50 (45 - 55) | 29 (24 - 34) |  |  |
| ALTCA recipients | 429,806 (100.00%) | 138,449 (65.84%) | 140,469 (23.62%) | 86,549 (6.10%) | 43,625 (1.86%) | 20,714 (0.92%) |  |  |
| Vaccinations against SARS-CoV-2: |  |  |  |  |  |  |  |  |
| Four doses | 0 (0.00%) | 0 (0.00%) | 0 (0.00%) | 0 (0.00%) | 0 (0.00%) | 0 (0.00%) |  |  |
| Three doses | 7,445 (1.73%) | 4,496 (2.14%) | 5,587 (0.94%) | 5,431 (0.38%) | 5,233 (0.22%) | 1,887 (0.08%) |  |  |
| Two doses | 323,588 (75.29%) | 162,656 (77.35%) | 501,107 (84.27%) | 1,114,198 (78.50%) | 1,602,247 (68.24%) | 1,236,121 (54.93%) |  |  |
| One dose | 16,617 (3.87%) | 5,671 (2.70%) | 13,955 (2.35%) | 43,097 (3.04%) | 130,043 (5.54%) | 195,510 (8.69%) |  |  |
| Unvaccinated | 0 (0.00%) | 0 (0.00%) | 0 (0.00%) | 0 (0.00%) | 0 (0.00%) | 0 (0.00%) |  |  |
| Time since last vaccination | 149 (100 - 188) | 186 (125 - 196) | 149 (112 - 177) | 112 (92 - 131) | 97 (77 - 117) | 82 (61 - 106) |  |  |
| Education: |  |  |  |  |  |  |  |  |
| Compulsory schooling or lower | 206,947 (48.15%) | 106,588 (50.69%) | 204,323 (34.36%) | 327,698 (23.09%) | 395,093 (16.83%) | 392,738 (17.45%) |  |  |
| Pre-secondary education | 179,180 (41.69%) | 78,979 (37.56%) | 299,594 (50.38%) | 780,965 (55.02%) | 1,168,862 (49.78%) | 826,190 (36.72%) |  |  |
| Secondary education | 24,758 (5.76%) | 13,564 (6.45%) | 44,274 (7.45%) | 129,399 (9.12%) | 335,036 (14.27%) | 522,724 (23.23%) |  |  |
| Pre-tertiary education | 4,384 (1.02%) | 1,913 (0.91%) | 8,390 (1.41%) | 53,189 (3.75%) | 75,741 (3.23%) | 35,620 (1.58%) |  |  |
| Tertiary education | 14,485 (3.37%) | 9,108 (4.33%) | 37,848 (6.36%) | 127,438 (8.98%) | 370,721 (15.79%) | 467,259 (20.77%) |  |  |
| Education unkown | 52 (0.01%) | 132 (0.06%) | 203 (0.03%) | 747 (0.05%) | 2,546 (0.11%) | 5,657 (0.25%) |  |  |
|  | BioNTec/Pfizer | Moderna | AstraZeneca | Janssen (J&J) |  |  |  |  |
| N | 3,548,687 | 486,965 | 736,431 | 255,156 |  |  |  |  |
| Females | 1,848,542 (52.09%) | 232,850 (47.82%) | 398,634 (54.13%) | 99,424 (38.97%) |  |  |  |  |
| Age | 53 (37 - 67) | 52 (39 - 65) | 56 (41 - 68) | 39 (27 - 53) |  |  |  |  |
| ALTCA recipients | 269,015 (7.58%) | 35,983 (7.39%) | 33,629 (4.57%) | 9,023 (3.54%) |  |  |  |  |
| Vaccinations against SARS-CoV-2: |  |  |  |  |  |  |  |  |
| Four doses | 0 (0.00%) | 0 (0.00%) | 0 (0.00%) | 0 (0.00%) |  |  |  |  |
| Three doses | 20,563 (0.58%) | 1,575 (0.32%) | 189 (0.03%) | 307 (0.12%) |  |  |  |  |
| Two doses | 3,411,792 (96.14%) | 475,625 (97.67%) | 728,429 (98.91%) | 483 (0.19%) |  |  |  |  |
| One dose | 116,332 (3.28%) | 9,765 (2.01%) | 7,813 (1.06%) | 254,366 (99.69%) |  |  |  |  |
| Unvaccinated | 0 (0.00%) | 0 (0.00%) | 0 (0.00%) | 0 (0.00%) |  |  |  |  |
| Time since last vaccination | 100 (73 - 131) | 99 (79 - 140) | 106 (89 - 124) | 85 (54 - 113) |  |  |  |  |
| Education: |  |  |  |  |  |  |  |  |
| Compulsory schooling or lower | 701,016 (19.75%) | 91,297 (18.75%) | 101,945 (13.84%) | 60,461 (23.70%) |  |  |  |  |
| Pre-secondary education | 1,642,008 (46.27%) | 221,553 (45.50%) | 322,333 (43.77%) | 116,701 (45.74%) |  |  |  |  |
| Secondary education | 561,268 (15.82%) | 78,673 (16.16%) | 106,512 (14.46%) | 44,811 (17.56%) |  |  |  |  |
| Pre-tertiary education | 82,631 (2.33%) | 11,691 (2.40%) | 44,062 (5.98%) | 3,228 (1.27%) |  |  |  |  |
| Tertiary education | 561,478 (15.82%) | 83,702 (17.19%) | 161,555 (21.94%) | 29,853 (11.70%) |  |  |  |  |
| Education unkown | 286 (0.01%) | 49 (0.01%) | 24 (0.00%) | 102 (0.04%) |  |  |  |  |

| **Table S7:** Characteristics of population for 2022 high COVID-19 disease burden on February 1. 2022 | | | | | | | | |
| --- | --- | --- | --- | --- | --- | --- | --- | --- |
|  | Total | Female | Male | four doses | three doses | two doses | one dose | unvaccinated |
| N | 6,031,104 | 3,081,059 | 2,950,045 | 2,315 | 3,722,785 | 1,318,443 | 117,277 | 870,284 |
| Females | 3,081,059 (51.09%) | 3,081,059 (100.00%) | 0 (0.00%) | 1,031 (44.54%) | 1,951,687 (52.43%) | 626,343 (47.51%) | 56,726 (48.37%) | 445,272 (51.16%) |
| Age | 52 (36 - 65) | 53 (37 - 67) | 50 (35 - 64) | 64 (53 - 74) | 57 (42 - 70) | 39 (28 - 54) | 40 (28 - 58) | 46 (33 - 61) |
| ALTCA recipients | 454,921 (7.54%) | 283,172 (9.19%) | 171,749 (5.82%) | 465 (20.09%) | 333,905 (8.97%) | 59,448 (4.51%) | 9,754 (8.32%) | 51,349 (5.90%) |
| Vaccinations against SARS-CoV-2: |  |  |  |  |  |  |  |  |
| Four doses | 2,315 (0.04%) | 1,031 (0.03%) | 1,284 (0.04%) | 2,315 (100.00%) | 0 (0.00%) | 0 (0.00%) | 0 (0.00%) | 0 (0.00%) |
| Three doses | 3,722,785 (61.73%) | 1,951,687 (63.34%) | 1,771,098 (60.04%) | 0 (0.00%) | 3,722,785 (100.00%) | 0 (0.00%) | 0 (0.00%) | 0 (0.00%) |
| Two doses | 1,318,443 (21.86%) | 626,343 (20.33%) | 692,100 (23.46%) | 0 (0.00%) | 0 (0.00%) | 1,318,443 (100.00%) | 0 (0.00%) | 0 (0.00%) |
| One dose | 117,277 (1.94%) | 56,726 (1.84%) | 60,551 (2.05%) | 0 (0.00%) | 0 (0.00%) | 0 (0.00%) | 117,277 (100.00%) | 0 (0.00%) |
| Unvaccinated | 0 (0.00%) | 0 (0.00%) | 0 (0.00%) | 0 (0.00%) | 0 (0.00%) | 0 (0.00%) | 0 (0.00%) | 0 (0.00%) |
| Time since last vaccination | 62 (46 - 78) | 63 (47 - 79) | 61 (45 - 77) | 20 (19 - 35) | 62 (48 - 75) | 63 (28 - 123) | 29 (12 - 84) | NA (NA - NA) |
| Education: |  |  |  |  |  |  |  |  |
| Compulsory schooling or lower | 1,285,407 (21.31%) | 780,099 (25.32%) | 505,308 (17.13%) | 262 (11.32%) | 662,200 (17.79%) | 352,655 (26.75%) | 42,181 (35.97%) | 228,109 (26.21%) |
| Pre-secondary education | 2,769,210 (45.92%) | 1,261,263 (40.94%) | 1,507,947 (51.12%) | 950 (41.04%) | 1,734,774 (46.60%) | 590,147 (44.76%) | 45,593 (38.88%) | 397,746 (45.70%) |
| Secondary education | 912,749 (15.13%) | 464,740 (15.08%) | 448,009 (15.19%) | 263 (11.36%) | 563,607 (15.14%) | 212,135 (16.09%) | 15,401 (13.13%) | 121,343 (13.94%) |
| Pre-tertiary education | 154,296 (2.56%) | 111,020 (3.60%) | 43,276 (1.47%) | 85 (3.67%) | 115,392 (3.10%) | 19,081 (1.45%) | 1,613 (1.38%) | 18,125 (2.08%) |
| Tertiary education | 898,763 (14.90%) | 459,603 (14.92%) | 439,160 (14.89%) | 755 (32.61%) | 646,599 (17.37%) | 143,726 (10.90%) | 12,060 (10.28%) | 95,623 (10.99%) |
| Education unkown | 10,671 (0.18%) | 4,330 (0.14%) | 6,341 (0.21%) | 0 (0.00%) | 213 (0.01%) | 699 (0.05%) | 428 (0.36%) | 9,331 (1.07%) |
|  | ALTCA recipients | 85+ year old | 75-84 year old | 60-74 year old | 40-59 year old | 18-39 year old |  |  |
| N | 454,921 | 202,335 | 577,102 | 1,341,723 | 2,037,884 | 1,872,060 |  |  |
| Females | 283,172 (62.25%) | 133,321 (65.89%) | 330,813 (57.32%) | 706,442 (52.65%) | 1,010,895 (49.61%) | 899,588 (48.05%) |  |  |
| Age | 80 (69 - 86) | 88 (86 - 91) | 79 (77 - 81) | 66 (63 - 70) | 51 (45 - 55) | 30 (24 - 34) |  |  |
| ALTCA recipients | 454,921 (100.00%) | 141,297 (69.83%) | 156,604 (27.14%) | 93,543 (6.97%) | 44,035 (2.16%) | 19,442 (1.04%) |  |  |
| Vaccinations against SARS-CoV-2: |  |  |  |  |  |  |  |  |
| Four doses | 465 (0.10%) | 103 (0.05%) | 447 (0.08%) | 897 (0.07%) | 673 (0.03%) | 195 (0.01%) |  |  |
| Three doses | 333,905 (73.40%) | 156,706 (77.45%) | 477,009 (82.66%) | 1,004,717 (74.88%) | 1,268,190 (62.23%) | 816,163 (43.60%) |  |  |
| Two doses | 59,448 (13.07%) | 18,878 (9.33%) | 45,287 (7.85%) | 154,701 (11.53%) | 439,302 (21.56%) | 660,275 (35.27%) |  |  |
| One dose | 9,754 (2.14%) | 3,784 (1.87%) | 6,486 (1.12%) | 16,770 (1.25%) | 31,865 (1.56%) | 58,372 (3.12%) |  |  |
| Unvaccinated | 0 (0.00%) | 0 (0.00%) | 0 (0.00%) | 0 (0.00%) | 0 (0.00%) | 0 (0.00%) |  |  |
| Time since last vaccination | 75 (57 - 104) | 88 (69 - 113) | 75 (62 - 90) | 64 (50 - 76) | 60 (45 - 74) | 56 (35 - 75) |  |  |
| Education: |  |  |  |  |  |  |  |  |
| Compulsory schooling or lower | 211,788 (46.55%) | 101,761 (50.29%) | 193,831 (33.59%) | 306,039 (22.81%) | 342,710 (16.82%) | 341,066 (18.22%) |  |  |
| Pre-secondary education | 194,925 (42.85%) | 76,643 (37.88%) | 293,384 (50.84%) | 737,001 (54.93%) | 1,003,146 (49.22%) | 659,036 (35.20%) |  |  |
| Secondary education | 26,990 (5.93%) | 13,093 (6.47%) | 43,882 (7.60%) | 124,158 (9.25%) | 293,782 (14.42%) | 437,834 (23.39%) |  |  |
| Pre-tertiary education | 4,933 (1.08%) | 1,849 (0.91%) | 8,432 (1.46%) | 50,852 (3.79%) | 64,495 (3.16%) | 28,668 (1.53%) |  |  |
| Tertiary education | 16,233 (3.57%) | 8,857 (4.38%) | 37,374 (6.48%) | 122,872 (9.16%) | 330,770 (16.23%) | 398,890 (21.31%) |  |  |
| Education unkown | 52 (0.01%) | 132 (0.07%) | 199 (0.03%) | 801 (0.06%) | 2,980 (0.15%) | 6,559 (0.35%) |  |  |
|  | BioNTec/Pfizer | Moderna | AstraZeneca | Janssen (J&J) |  |  |  |  |
| N | 4,603,489 | 495,157 | 28,930 | 33,244 |  |  |  |  |
| Females | 2,372,709 (51.54%) | 236,548 (47.77%) | 13,715 (47.41%) | 12,815 (38.55%) |  |  |  |  |
| Age | 53 (36 - 66) | 54 (42 - 64) | 46 (31 - 61) | 41 (29 - 55) |  |  |  |  |
| ALTCA recipients | 367,474 (7.98%) | 31,814 (6.43%) | 2,385 (8.24%) | 1,899 (5.71%) |  |  |  |  |
| Vaccinations against SARS-CoV-2: |  |  |  |  |  |  |  |  |
| Four doses | 1,907 (0.04%) | 403 (0.08%) | 1 (0.00%) | 4 (0.01%) |  |  |  |  |
| Three doses | 3,340,394 (72.56%) | 381,174 (76.98%) | 684 (2.36%) | 533 (1.60%) |  |  |  |  |
| Two doses | 1,174,385 (25.51%) | 107,113 (21.63%) | 27,037 (93.46%) | 9,908 (29.80%) |  |  |  |  |
| One dose | 86,803 (1.89%) | 6,467 (1.31%) | 1,208 (4.18%) | 22,799 (68.58%) |  |  |  |  |
| Unvaccinated | 0 (0.00%) | 0 (0.00%) | 0 (0.00%) | 0 (0.00%) |  |  |  |  |
| Time since last vaccination | 62 (46 - 78) | 60 (45 - 74) | 144 (8 - 213) | 88 (33 - 165) |  |  |  |  |
| Education: |  |  |  |  |  |  |  |  |
| Compulsory schooling or lower | 953,485 (20.71%) | 86,409 (17.45%) | 6,219 (21.50%) | 11,185 (33.65%) |  |  |  |  |
| Pre-secondary education | 2,115,386 (45.95%) | 230,982 (46.65%) | 12,095 (41.81%) | 13,001 (39.11%) |  |  |  |  |
| Secondary education | 705,584 (15.33%) | 76,136 (15.38%) | 4,852 (16.77%) | 4,834 (14.54%) |  |  |  |  |
| Pre-tertiary education | 120,984 (2.63%) | 13,857 (2.80%) | 866 (2.99%) | 464 (1.40%) |  |  |  |  |
| Tertiary education | 706,909 (15.36%) | 87,673 (17.71%) | 4,877 (16.86%) | 3,681 (11.07%) |  |  |  |  |
| Education unkown | 1,140 (0.02%) | 100 (0.02%) | 21 (0.07%) | 79 (0.24%) |  |  |  |  |

| **Table S8:** Number of documented deaths in the studied population. | | | |
| --- | --- | --- | --- |
|  | **cause of death** | | |
|  | **All-cause** | **cancer** | **external causes** |
| **2021** | 76,045 | 18,886 | 4,715 |
| **2022** | 41,963 | 10,138 | 3,470 |
| **total** | 118,008 | 29,024 | 8,185 |

### **Analyses Tables of different observation periods (S9 – S15)**

| **Table S9:** Hazard ratios (HR) with 95% confidence intervals (95% CI) for all-cause mortality according to number of SARS-CoV-2 vaccine doses between 1, January 2021 and 31, August 2022. | | | | | |
| --- | --- | --- | --- | --- | --- |
|  | All-cause mortality | | | | |
|  | Four vaccine doses | Three vaccine doses | Two vaccine doses | One vaccine dose | Unvaccinated |
| Deaths (n) | 1156 | 32515 | 35189 | 7206 | 41942 |
| Events per 100,000 person days | 6.32 | 3.53 | 3.45 | 2.65 | 2.67 |
| Crude HR (95%CI) | 2.03 (1.88 - 2.18) | 1.16 (1.13 - 1.18) | 1.36 (1.33 - 1.38) | 1.08 (1.05 - 1.11) | Reference |
| Age adjusted HR (95% CI) | 0.39 (0.37 - 0.42) | 0.48 (0.47 - 0.49) | 0.65 (0.64 - 0.66) | 0.74 (0.72 - 0.76) | Reference |
| Age and gender adjusted HR (95% CI) | 0.37 (0.35 - 0.40) | 0.46 (0.45 - 0.48) | 0.63 (0.62 - 0.64) | 0.73 (0.71 - 0.75) | Reference |
| Age, sex and ALTCA adjusted HR (95% CI) | 0.39 (0.36 - 0.42) | 0.48 (0.47 - 0.49) | 0.63 (0.62 - 0.64) | 0.74 (0.72 - 0.76) | Reference |
| Age, sex, ALTCA and education adjusted HR (95% CI) | 0.39 (0.36 - 0.42) | 0.47 (0.46 - 0.48) | 0.63 (0.62 - 0.64) | 0.74 (0.72 - 0.76) | Reference |

| **Table S10:** Hazard ratios (HR) with 95% confidence intervals (95% CI) for cancer deaths and deaths by external causes according to number of SARS-CoV-2 vaccine doses between 1, January 2021 and 31, August 2022. | | | | | |
| --- | --- | --- | --- | --- | --- |
|  | Cancer | | | | |
|  | Four vaccine doses | Three vaccine doses | Two vaccine doses | One vaccine dose | Unvaccinated |
| Deaths (n) | 240 | 7805 | 8927 | 1806 | 10246 |
| Events per 100,000 person days | 1.31 | 0.85 | 0.87 | 0.66 | 0.65 |
| Crude HR (95%CI) | 2.40 (2.03 - 2.84) | 1.35 (1.28 - 1.42) | 1.43 (1.38 - 1.48) | 1.07 (1.02 - 1.13) | Reference |
| Age adjusted HR (95% CI) | 0.54 (0.46 - 0.64) | 0.62 (0.59 - 0.65) | 0.73 (0.71 - 0.76) | 0.73 (0.69 - 0.77) | Reference |
| Age and gender adjusted HR (95% CI) | 0.52 (0.44 - 0.61) | 0.60 (0.57 - 0.63) | 0.72 (0.69 - 0.74) | 0.72 (0.68 - 0.76) | Reference |
| Age, sex and ALTCA adjusted HR (95% CI) | 0.53 (0.45 - 0.63) | 0.62 (0.59 - 0.65) | 0.72 (0.69 - 0.74) | 0.74 (0.70 - 0.78) | Reference |
| Age, sex, ALTCA and education adjusted HR (95% CI) | 0.52 (0.44 - 0.62) | 0.60 (0.58 - 0.64) | 0.70 (0.68 - 0.73) | 0.73 (0.69 - 0.77) | Reference |
|  | External cause | | | | |
|  | Four vaccine doses | Three vaccine doses | Two vaccine doses | One vaccine dose | Unvaccinated |
| Deaths (n) | 72 | 2437 | 2360 | 553 | 2763 |
| Events per 100,000 person days | 0.33 | 0.2 | 0.19 | 0.16 | 0.15 |
| Crude HR (95%CI) | 1.97 (1.50 - 2.59) | 1.34 (1.23 - 1.46) | 1.31 (1.22 - 1.40) | 1.09 (0.99 - 1.20) | Reference |
| Age adjusted HR (95% CI) | 0.52 (0.40 - 0.68) | 0.68 (0.62 - 0.74) | 0.81 (0.76 - 0.87) | 0.87 (0.79 - 0.96) | Reference |
| Age and gender adjusted HR (95% CI) | 0.48 (0.37 - 0.63) | 0.65 (0.60 - 0.71) | 0.80 (0.74 - 0.85) | 0.85 (0.77 - 0.93) | Reference |
| Age, sex and ALTCA adjusted HR (95% CI) | 0.48 (0.36 - 0.63) | 0.67 (0.62 - 0.73) | 0.79 (0.74 - 0.84) | 0.85 (0.77 - 0.94) | Reference |
| Age, sex, ALTCA and education adjusted HR (95% CI) | 0.50 (0.38 - 0.66) | 0.68 (0.62 - 0.74) | 0.79 (0.74 - 0.84) | 0.85 (0.77 - 0.93) | Reference |

| **Table S11:** Hazard ratios (HR) with 95% confidence intervals (95% CI) for all-cause mortality according to number of SARS-CoV-2 vaccine doses for the period from 2021 to 31. August 2022 split into 3 month intervals. | | | | | | | | | | | | | | | | | | | | |
| --- | --- | --- | --- | --- | --- | --- | --- | --- | --- | --- | --- | --- | --- | --- | --- | --- | --- | --- | --- | --- |
|  | 2021 | | | | | | | | | | | | | | | | | | | |
|  | All-cause mortality | | | | | | | | | | | | | | | | | | | |
|  | first quarter | | | | | second quarter | | | | | third quarter | | | | | fourth quarter | | | | |
|  | Four vaccine doses | Three vaccine doses | Two vaccine doses | One vaccine dose | Unvaccinated | Four vaccine doses | Three vaccine doses | Two vaccine doses | One vaccine dose | Unvaccinated | Four vaccine doses | Three vaccine doses | Two vaccine doses | One vaccine dose | Unvaccinated | Four vaccine doses | Three vaccine doses | Two vaccine doses | One vaccine dose | Unvaccinated |
| Deaths (n) |  |  | 1374 | 1371 | 15583 |  | 1 | 6910 | 2546 | 8823 |  | 100 | 11583 | 1388 | 5842 | 1 | 5368 | 9168 | 974 | 5013 |
| Events per 100,000 person days |  |  | 9.89 | 6.04 | 2.50 |  |  | 5.81 | 1.79 | 2.26 |  | 12.6 | 2.98 | 2.43 | 2.84 |  | 4.31 | 2.84 | 2.91 | 3.38 |
| Crude HR (95%CI) |  |  | 4.31 (4.07 - 4.57) | 2.69 (2.54 - 2.85) | Ref |  |  | 2.74 (2.65 - 2.84) | 0.79 (0.75 - 0.82) | Ref |  | 4.31 (3.52 - 5.28) | 1.05 (1.02 - 1.08) | 0.89 (0.84 - 0.94) | Ref |  | 1.54 (1.47 - 1.61) | 0.85 (0.82 - 0.88) | 0.85 (0.80 - 0.91) | Ref |
| Age adjusted HR (95% CI) |  |  | 0.96 (0.90 - 1.02) | 0.75 (0.71 - 0.80) | Ref |  |  | 0.46 (0.44 - 0.47) | 0.44 (0.42 - 0.46) | Ref |  | 0.63 (0.51 - 0.77) | 0.47 (0.45 - 0.48) | 1.18 (1.11 - 1.25) | Ref |  | 0.37 (0.36 - 0.39) | 0.70 (0.67 - 0.72) | 1.03 (0.96 - 1.11) | Ref |
| Age and gender adjusted HR (95% CI) |  |  | 0.98 (0.92 - 1.04) | 0.75 (0.71 - 0.80) | Ref |  |  | 0.45 (0.43 - 0.46) | 0.43 (0.41 - 0.45) | Ref |  | 0.61 (0.50 - 0.75) | 0.45 (0.44 - 0.47) | 1.15 (1.09 - 1.22) | Ref |  | 0.36 (0.34 - 0.38) | 0.68 (0.66 - 0.70) | 1.01 (0.95 - 1.09) | Ref |
| Age, sex and ALTCA adjusted HR (95% CI) |  |  | 0.75 (0.71 - 0.80) | 0.66 (0.63 - 0.70) | Ref |  |  | 0.46 (0.44 - 0.48) | 0.52 (0.50 - 0.54) | Ref |  | 0.51 (0.42 - 0.63) | 0.51 (0.50 - 0.53) | 1.10 (1.03 - 1.16) | Ref |  | 0.38 (0.37 - 0.40) | 0.73 (0.71 - 0.76) | 0.93 (0.87 - 1.00) | Ref |
| Age, sex, ALTCA and education adjusted HR (95% CI) |  |  | 0.75 (0.71 - 0.80) | 0.67 (0.63 - 0.70) | Ref |  |  | 0.46 (0.44 - 0.47) | 0.52 (0.49 - 0.54) | Ref |  | 0.51 (0.42 - 0.63) | 0.51 (0.49 - 0.53) | 1.09 (1.02 - 1.15) | Ref |  | 0.38 (0.37 - 0.40) | 0.72 (0.70 - 0.75) | 0.91 (0.85 - 0.98) | Ref |
|  | 2022 | | | | | | | | | | | | | | | | | | | |
| Deaths (n) | 25 | 11061 | 3199 | 567 | 3174 | 199 | 9924 | 1953 | 238 | 2114 | 931 | 6061 | 1002 | 122 | 1393 |  |  |  |  | 0 |
| Events per 100,000 person days | 8.3 | 3.28 | 3.42 | 6.58 | 3.52 | 7.82 | 3.39 | 3.64 | 4.86 | 3.07 | 6.03 | 3.61 | 3.33 | 3.96 | 3.18 |  |  |  |  |  |
| Crude HR (95%CI) | 2.31 (1.56 - 3.43) | 0.93 (0.89 - 0.96) | 0.98 (0.94 - 1.03) | 1.88 (1.72 - 2.06) | Ref | 2.62 (2.26 - 3.04) | 1.10 (1.05 - 1.16) | 1.18 (1.11 - 1.26) | 1.58 (1.39 - 1.81) | Ref | 1.87 (1.72 - 2.03) | 1.14 (1.07 - 1.21) | 1.05 (0.97 - 1.14) | 1.20 (1.00 - 1.45) | Ref |  |  |  |  | Ref |
| Age adjusted HR (95% CI) | 0.71 (0.48 - 1.06) | 0.45 (0.43 - 0.47) | 1.13 (1.08 - 1.19) | 1.52 (1.39 - 1.67) | Ref | 0.45 (0.39 - 0.52) | 0.57 (0.55 - 0.60) | 1.22 (1.15 - 1.30) | 1.63 (1.43 - 1.87) | Ref | 0.38 (0.35 - 0.42) | 0.64 (0.61 - 0.68) | 1.13 (1.04 - 1.23) | 1.29 (1.07 - 1.55) | Ref |  |  |  |  | Ref |
| Age and gender adjusted HR (95% CI) | 0.66 (0.44 - 0.97) | 0.44 (0.42 - 0.45) | 1.12 (1.06 - 1.17) | 1.51 (1.38 - 1.66) | Ref | 0.42 (0.37 - 0.49) | 0.55 (0.53 - 0.58) | 1.21 (1.14 - 1.29) | 1.62 (1.42 - 1.85) | Ref | 0.37 (0.34 - 0.40) | 0.63 (0.59 - 0.67) | 1.12 (1.03 - 1.21) | 1.28 (1.06 - 1.54) | Ref |  |  |  |  | Ref |
| Age, sex and ALTCA adjusted HR (95% CI) | 0.60 (0.40 - 0.89) | 0.45 (0.44 - 0.47) | 0.98 (0.93 - 1.03) | 1.29 (1.18 - 1.41) | Ref | 0.43 (0.37 - 0.50) | 0.57 (0.54 - 0.59) | 1.04 (0.98 - 1.11) | 1.43 (1.25 - 1.63) | Ref | 0.39 (0.36 - 0.43) | 0.63 (0.59 - 0.66) | 0.96 (0.88 - 1.04) | 1.14 (0.95 - 1.37) | Ref |  |  |  |  | Ref |
| Age, sex, ALTCA and education adjusted HR (95% CI) | 0.62 (0.42 - 0.92) | 0.45 (0.43 - 0.46) | 0.96 (0.91 - 1.01) | 1.27 (1.16 - 1.39) | Ref | 0.44 (0.38 - 0.52) | 0.56 (0.53 - 0.59) | 1.02 (0.96 - 1.08) | 1.39 (1.22 - 1.60) | Ref | 0.39 (0.36 - 0.43) | 0.61 (0.58 - 0.65) | 0.94 (0.86 - 1.02) | 1.11 (0.92 - 1.34) | Ref |  |  |  |  | Ref |

| **Table S12:** Hazard ratios (HR) with 95% confidence intervals (95% CI) for cancer and external causes mortality according to number of SARS-CoV-2 vaccine doses for the period from 2021 to 2022 split into 3 month intervals. | | | | | | | | | | | | | | | | | | | | |
| --- | --- | --- | --- | --- | --- | --- | --- | --- | --- | --- | --- | --- | --- | --- | --- | --- | --- | --- | --- | --- |
|  | 2021 | | | | | | | | | | | | | | | | | | | |
|  | first quarter | | | | | second quarter | | | | | third quarter | | | | | fourth quarter | | | | |
|  | Four vaccine doses | Three vaccine doses | Two vaccine doses | One vaccine dose | Unvaccinated | Four vaccine doses | Three vaccine doses | Two vaccine doses | One vaccine dose | Unvaccinated | Four vaccine doses | Three vaccine doses | Two vaccine doses | One vaccine dose | Unvaccinated | Four vaccine doses | Three vaccine doses | Two vaccine doses | One vaccine dose | Unvaccinated |
|  | Cancer | | | | | Cancer | | | | | Cancer | | | | | Cancer | | | | |
| Deaths (n) |  |  | 162 | 226 | 4145 |  |  | 1498 | 718 | 2414 |  | 11 | 2958 | 420 | 1409 |  | 891 | 2750 | 227 | 1057 |
| Events per 100,000 person days |  |  | 1.17 | 1 | 0.66 |  |  | 1.26 | 0.5 | 0.62 |  | 1.39 | 0.76 | 0.74 | 0.68 |  | 0.71 | 0.85 | 0.68 | 0.71 |
| Crude HR (95%CI) |  |  | 1.84 (1.57 - 2.15) | 1.56 (1.36 - 1.78) | Ref |  |  | 2.13 (1.98 - 2.28) | 0.83 (0.76 - 0.90) | Ref |  | 2.18 (1.20 - 3.99) | 1.12 (1.05 - 1.20) | 1.09 (0.97 - 1.22) | Ref |  | 1.11 (1.00 - 1.23) | 1.21 (1.13 - 1.30) | 0.95 (0.82 - 1.10) | Ref |
| Age adjusted HR (95% CI) |  |  | 0.54 (0.46 - 0.63) | 0.55 (0.48 - 0.63) | Ref |  |  | 0.44 (0.41 - 0.48) | 0.45 (0.41 - 0.49) | Ref |  | 0.39 (0.21 - 0.71) | 0.53 (0.50 - 0.57) | 1.38 (1.23 - 1.54) | Ref |  | 0.35 (0.31 - 0.38) | 0.98 (0.91 - 1.05) | 1.12 (0.97 - 1.29) | Ref |
| Age and gender adjusted HR (95% CI) |  |  | 0.55 (0.47 - 0.65) | 0.55 (0.48 - 0.63) | Ref |  |  | 0.43 (0.40 - 0.46) | 0.44 (0.40 - 0.48) | Ref |  | 0.38 (0.21 - 0.70) | 0.52 (0.48 - 0.55) | 1.35 (1.21 - 1.51) | Ref |  | 0.33 (0.30 - 0.37) | 0.95 (0.89 - 1.03) | 1.10 (0.95 - 1.27) | Ref |
| Age, sex and ALTCA adjusted HR (95% CI) |  |  | 0.38 (0.32 - 0.45) | 0.45 (0.40 - 0.52) | Ref |  |  | 0.43 (0.40 - 0.46) | 0.56 (0.51 - 0.61) | Ref |  | 0.29 (0.16 - 0.53) | 0.59 (0.56 - 0.63) | 1.29 (1.16 - 1.45) | Ref |  | 0.36 (0.32 - 0.39) | 1.05 (0.98 - 1.13) | 0.99 (0.86 - 1.14) | Ref |
| Age, sex, ALTCA and education adjusted HR (95% CI) |  |  | 0.38 (0.33 - 0.45) | 0.46 (0.40 - 0.52) | Ref |  |  | 0.42 (0.39 - 0.45) | 0.54 (0.50 - 0.59) | Ref |  | 0.28 (0.15 - 0.51) | 0.58 (0.54 - 0.62) | 1.26 (1.13 - 1.40) | Ref |  | 0.35 (0.31 - 0.38) | 1.02 (0.95 - 1.10) | 0.97 (0.84 - 1.12) | Ref |
|  | External cause | | | | | External cause | | | | | External cause | | | | | External cause | | | | |
| Deaths (n) |  |  | 47 | 83 | 919 |  |  | 307 | 171 | 545 |  | 5 | 763 | 96 | 335 |  | 272 | 528 | 56 | 255 |
| Events per 100,000 person days |  |  | 0.34 | 0.37 | 0.15 |  |  | 0.26 | 0.12 | 0.14 |  |  | 0.2 | 0.17 | 0.16 |  | 0.22 | 0.16 | 0.17 | 0.17 |
| Crude HR (95%CI) |  |  | 2.38 (1.77 - 3.21) | 2.65 (2.10 - 3.34) | Ref |  |  | 1.86 (1.60 - 2.16) | 0.81 (0.68 - 0.97) | Ref |  |  | 1.21 (1.06 - 1.37) | 1.08 (0.86 - 1.36) | Ref |  | 1.58 (1.29 - 1.93) | 0.95 (0.82 - 1.10) | 0.97 (0.72 - 1.29) | Ref |
| Age adjusted HR (95% CI) |  |  | 1.04 (0.76 - 1.41) | 1.27 (1.00 - 1.62) | Ref |  |  | 0.59 (0.50 - 0.69) | 0.51 (0.43 - 0.61) | Ref |  |  | 0.63 (0.55 - 0.72) | 1.22 (0.97 - 1.54) | Ref |  | 0.47 (0.39 - 0.57) | 0.73 (0.63 - 0.85) | 1.11 (0.83 - 1.48) | Ref |
| Age and gender adjusted HR (95% CI) |  |  | 1.10 (0.81 - 1.50) | 1.30 (1.03 - 1.66) | Ref |  |  | 0.57 (0.49 - 0.68) | 0.50 (0.42 - 0.59) | Ref |  |  | 0.61 (0.53 - 0.69) | 1.17 (0.93 - 1.47) | Ref |  | 0.45 (0.37 - 0.54) | 0.71 (0.61 - 0.82) | 1.06 (0.80 - 1.42) | Ref |
| Age, sex and ALTCA adjusted HR (95% CI) |  |  | 0.78 (0.58 - 1.07) | 1.08 (0.85 - 1.37) | Ref |  |  | 0.56 (0.47 - 0.66) | 0.58 (0.49 - 0.69) | Ref |  |  | 0.67 (0.59 - 0.77) | 1.15 (0.91 - 1.44) | Ref |  | 0.47 (0.39 - 0.57) | 0.77 (0.66 - 0.89) | 1.00 (0.75 - 1.33) | Ref |
| Age, sex, ALTCA and education adjusted HR (95% CI) |  |  | 0.79 (0.58 - 1.08) | 1.09 (0.86 - 1.39) | Ref |  |  | 0.57 (0.48 - 0.67) | 0.58 (0.49 - 0.70) | Ref |  |  | 0.68 (0.59 - 0.77) | 1.16 (0.92 - 1.45) | Ref |  | 0.47 (0.39 - 0.57) | 0.76 (0.65 - 0.88) | 0.97 (0.72 - 1.29) | Ref |

**Table S12 continued:**

|  | 2022 | | | | | | | | | | | | | | | | | | | |
| --- | --- | --- | --- | --- | --- | --- | --- | --- | --- | --- | --- | --- | --- | --- | --- | --- | --- | --- | --- | --- |
|  | first quarter | | | | | second quarter | | | | | third quarter | | | | | fourth quarter | | | | |
|  | Four vaccine doses | Three vaccine doses | Two vaccine doses | One vaccine dose | Unvaccinated | Four vaccine doses | Three vaccine doses | Two vaccine doses | One vaccine dose | Unvaccinated | Four vaccine doses | Three vaccine doses | Two vaccine doses | One vaccine dose | Unvaccinated | Four vaccine doses | Three vaccine doses | Two vaccine doses | One vaccine dose | Unvaccinated |
|  | Cancer | | | | | Cancer | | | | | Cancer | | | | | Cancer | | | | |
| Deaths (n) | 16 | 2688 | 906 | 134 | 570 | 38 | 2584 | 455 | 57 | 404 | 186 | 1631 | 198 | 24 | 247 |  |  |  |  | 0 |
| Events per 100,000 person days | 5.31 | 0.8 | 0.97 | 1.55 | 0.63 | 1.49 | 0.88 | 0.85 | 1.16 | 0.59 | 1.2 | 0.97 | 0.66 | 0.78 | 0.56 |  |  |  |  |  |
| Crude HR (95%CI) | 8.35 (5.07 - 13.75) | 1.25 (1.14 - 1.37) | 1.53 (1.38 - 1.70) | 2.48 (2.05 - 2.99) | Ref | 2.73 (1.94 - 3.84) | 1.51 (1.36 - 1.67) | 1.44 (1.26 - 1.65) | 1.99 (1.50 - 2.62) | Ref | 2.07 (1.71 - 2.52) | 1.73 (1.52 - 1.98) | 1.18 (0.98 - 1.42) | 1.32 (0.87 - 2.01) | Ref |  |  |  |  | Ref |
| Age adjusted HR (95% CI) | 2.92 (1.77 - 4.81) | 0.65 (0.60 - 0.72) | 1.76 (1.59 - 1.96) | 2.17 (1.79 - 2.62) | Ref | 0.58 (0.41 - 0.82) | 0.84 (0.75 - 0.93) | 1.50 (1.32 - 1.72) | 2.09 (1.58 - 2.76) | Ref | 0.55 (0.45 - 0.68) | 1.04 (0.91 - 1.19) | 1.27 (1.05 - 1.53) | 1.43 (0.94 - 2.17) | Ref |  |  |  |  | Ref |
| Age and gender adjusted HR (95% CI) | 2.80 (1.70 - 4.62) | 0.63 (0.58 - 0.69) | 1.74 (1.57 - 1.93) | 2.16 (1.79 - 2.61) | Ref | 0.56 (0.39 - 0.79) | 0.81 (0.73 - 0.91) | 1.49 (1.31 - 1.71) | 2.07 (1.57 - 2.74) | Ref | 0.54 (0.44 - 0.66) | 1.02 (0.89 - 1.17) | 1.26 (1.04 - 1.52) | 1.42 (0.93 - 2.16) | Ref |  |  |  |  | Ref |
| Age, sex and ALTCA adjusted HR (95% CI) | 2.26 (1.37 - 3.73) | 0.68 (0.62 - 0.74) | 1.49 (1.34 - 1.66) | 1.69 (1.40 - 2.05) | Ref | 0.53 (0.37 - 0.75) | 0.85 (0.77 - 0.95) | 1.24 (1.08 - 1.42) | 1.74 (1.32 - 2.30) | Ref | 0.56 (0.46 - 0.69) | 1.04 (0.91 - 1.19) | 1.06 (0.88 - 1.28) | 1.24 (0.81 - 1.89) | Ref |  |  |  |  | Ref |
| Age, sex, ALTCA and education adjusted HR (95% CI) | 2.18 (1.32 - 3.61) | 0.65 (0.60 - 0.72) | 1.46 (1.32 - 1.63) | 1.69 (1.40 - 2.05) | Ref | 0.53 (0.37 - 0.75) | 0.82 (0.74 - 0.91) | 1.23 (1.07 - 1.41) | 1.71 (1.29 - 2.26) | Ref | 0.57 (0.46 - 0.70) | 1.02 (0.89 - 1.17) | 1.06 (0.88 - 1.28) | 1.20 (0.78 - 1.84) | Ref |  |  |  |  | Ref |
|  | External cause | | | | | External cause | | | | | External cause | | | | | External cause | | | | |
| Deaths (n) |  | 644 | 203 | 38 | 167 | 11 | 677 | 155 | 15 | 119 | 48 | 363 | 66 | 8 | 93 |  |  |  |  | 0 |
| Events per 100,000 person days |  | 0.19 | 0.22 | 0.44 | 0.19 | 0.43 | 0.23 | 0.29 | 0.31 | 0.17 | 0.31 | 0.22 | 0.22 |  | 0.21 |  |  |  |  |  |
| Crude HR (95%CI) |  | 1.02 (0.86 - 1.21) | 1.19 (0.97 - 1.46) | 2.35 (1.65 - 3.34) | Ref | 2.40 (1.28 - 4.51) | 1.34 (1.10 - 1.63) | 1.68 (1.32 - 2.13) | 1.78 (1.04 - 3.04) | Ref | 1.43 (1.01 - 2.04) | 1.03 (0.82 - 1.29) | 1.04 (0.76 - 1.43) |  | Ref |  |  |  |  | Ref |
| Age adjusted HR (95% CI) |  | 0.58 (0.49 - 0.69) | 1.29 (1.05 - 1.59) | 2.29 (1.61 - 3.26) | Ref | 0.78 (0.41 - 1.50) | 0.82 (0.67 - 0.99) | 1.75 (1.38 - 2.23) | 1.89 (1.11 - 3.23) | Ref | 0.45 (0.30 - 0.65) | 0.66 (0.52 - 0.83) | 1.10 (0.80 - 1.51) |  | Ref |  |  |  |  | Ref |
| Age and gender adjusted HR (95% CI) |  | 0.56 (0.47 - 0.67) | 1.26 (1.03 - 1.55) | 2.26 (1.59 - 3.22) | Ref | 0.70 (0.36 - 1.35) | 0.79 (0.65 - 0.96) | 1.71 (1.34 - 2.17) | 1.84 (1.08 - 3.15) | Ref | 0.42 (0.29 - 0.62) | 0.64 (0.51 - 0.81) | 1.07 (0.78 - 1.47) |  | Ref |  |  |  |  | Ref |
| Age, sex and ALTCA adjusted HR (95% CI) |  | 0.59 (0.50 - 0.71) | 1.17 (0.95 - 1.44) | 1.96 (1.37 - 2.80) | Ref | 0.64 (0.33 - 1.23) | 0.82 (0.67 - 1.00) | 1.56 (1.23 - 1.99) | 1.64 (0.96 - 2.81) | Ref | 0.43 (0.29 - 0.64) | 0.65 (0.52 - 0.82) | 0.98 (0.71 - 1.34) |  | Ref |  |  |  |  | Ref |
| Age, sex, ALTCA and education adjusted HR (95% CI) |  | 0.59 (0.49 - 0.70) | 1.09 (0.89 - 1.34) | 1.86 (1.30 - 2.66) | Ref | 0.71 (0.37 - 1.37) | 0.80 (0.65 - 0.97) | 1.45 (1.14 - 1.84) | 1.46 (0.84 - 2.55) | Ref | 0.44 (0.30 - 0.65) | 0.64 (0.50 - 0.80) | 0.91 (0.66 - 1.25) |  | Ref |  |  |  |  | Ref |

| **Table S13:** Hazard ratios (HR) with 95% confidence intervals (95% CI) for all-cause mortality according to number of SARS-CoV-2 vaccine doses for the period from 2021 to 2022 split into 3 month intervals, with the reference beeing 1 less vaccination than the dose of interest. | | | | | | | | | | | | | | | | | | | | |
| --- | --- | --- | --- | --- | --- | --- | --- | --- | --- | --- | --- | --- | --- | --- | --- | --- | --- | --- | --- | --- |
|  | 2021 | | | | | | | | | | | | | | | | | | | |
|  | All-cause mortality | | | | | | | | | | | | | | | | | | | |
|  | first quarter | | | | | second quarter | | | | | third quarter | | | | | fourth quarter | | | | |
|  | Four or more vaccine doses | Three vaccine doses | Two vaccine doses | One vaccine dose | Unvaccinated | Four or more vaccine doses | Three vaccine doses | Two vaccine doses | One vaccine dose | Unvaccinated | Four or more vaccine doses | Three vaccine doses | Two vaccine doses | One vaccine dose | Unvaccinated | Four or more vaccine doses | Three vaccine doses | Two vaccine doses | One vaccine dose | Unvaccinated |
| Deaths (n) |  |  | 1374 | 1371 | 15583 |  | 1 | 6910 | 2546 | 8823 |  | 100 | 11583 | 1388 | 5842 | 1 | 5368 | 9168 | 974 | 5013 |
| Events per 100,000 person days |  |  | 9.89 | 6.04 | 2.50 |  |  | 5.81 | 1.79 | 2.26 |  | 12.6 | 2.98 | 2.43 | 2.84 |  | 4.31 | 2.84 | 2.91 | 3.38 |
| Crude HR (95%CI) |  |  | 1.77 (1.63 - 1.92) | 2.69 (2.54 - 2.85) | Ref |  |  | 3.48 (3.33 - 3.65) | 0.79 (0.75 - 0.82) | Ref |  | 4.25 (3.48 - 5.18) | 1.21 (1.14 - 1.28) | 0.89 (0.84 - 0.94) | Ref |  | 1.81 (1.74 - 1.89) | 0.98 (0.92 - 1.05) | 0.85 (0.80 - 0.91) | Ref |
| Age adjusted HR (95% CI) |  |  | 1.47 (1.35 - 1.60) | 0.75 (0.71 - 0.80) | Ref |  |  | 0.98 (0.93 - 1.03) | 0.44 (0.42 - 0.46) | Ref |  | 1.15 (0.94 - 1.41) | 0.38 (0.36 - 0.40) | 1.18 (1.11 - 1.25) | Ref |  | 0.44 (0.42 - 0.46) | 0.73 (0.68 - 0.78) | 1.03 (0.96 - 1.11) | Ref |
| Age and gender adjusted HR (95% CI) |  |  | 1.49 (1.38 - 1.63) | 0.75 (0.71 - 0.80) | Ref |  |  | 0.98 (0.93 - 1.03) | 0.43 (0.41 - 0.45) | Ref |  | 1.17 (0.95 - 1.42) | 0.38 (0.36 - 0.40) | 1.15 (1.09 - 1.22) | Ref |  | 0.43 (0.42 - 0.45) | 0.72 (0.67 - 0.77) | 1.01 (0.95 - 1.09) | Ref |
| Age, sex and ALTCA adjusted HR (95% CI) |  |  | 1.21 (1.11 - 1.32) | 0.66 (0.63 - 0.70) | Ref |  |  | 0.88 (0.84 - 0.93) | 0.52 (0.50 - 0.54) | Ref |  | 0.91 (0.75 - 1.12) | 0.46 (0.44 - 0.49) | 1.10 (1.03 - 1.16) | Ref |  | 0.46 (0.44 - 0.47) | 0.84 (0.78 - 0.89) | 0.93 (0.87 - 1.00) | Ref |
| Age, sex, ALTCA and education adjusted HR (95% CI) |  |  | 1.21 (1.12 - 1.32) | 0.67 (0.63 - 0.70) | Ref |  |  | 1.03 (0.98 - 1.08) | 0.52 (0.49 - 0.54) | Ref |  | 0.92 (0.75 - 1.12) | 0.46 (0.44 - 0.49) | 1.09 (1.02 - 1.15) | Ref |  | 0.40 (0.39 - 0.42) | 0.84 (0.78 - 0.89) | 0.91 (0.85 - 0.98) | Ref |
|  | 2022 | | | | | | | | | | | | | | | | | | | |
| Deaths (n) | 25 | 11061 | 3199 | 567 | 3174 | 199 | 9924 | 1953 | 238 | 2114 | 931 | 6061 | 1002 | 122 | 1393 |  |  |  |  |  |
| Events per 100,000 person days | 8.3 | 3.28 | 3.42 | 6.58 | 3.52 | 7.82 | 3.39 | 3.64 | 4.86 | 3.07 | 6.03 | 3.61 | 3.33 | 3.96 | 3.18 |  |  |  |  |  |
| Crude HR (95%CI) | 2.51 (1.69 - 3.71) | 0.95 (0.91 - 0.98) | 0.52 (0.47 - 0.57) | 1.88 (1.72 - 2.06) | Ref | 2.25 (1.96 - 2.60) | 0.93 (0.89 - 0.98) | 0.75 (0.65 - 0.85) | 1.58 (1.39 - 1.81) | Ref | 1.64 (1.53 - 1.76) | 1.08 (1.01 - 1.16) | 0.87 (0.72 - 1.05) | 1.20 (1.00 - 1.45) | Ref |  |  |  |  | Ref |
| Age adjusted HR (95% CI) | 1.50 (1.01 - 2.22) | 0.40 (0.38 - 0.41) | 0.73 (0.67 - 0.80) | 1.52 (1.39 - 1.67) | Ref | 0.74 (0.64 - 0.85) | 0.47 (0.45 - 0.49) | 0.74 (0.65 - 0.85) | 1.63 (1.43 - 1.87) | Ref | 0.59 (0.55 - 0.63) | 0.57 (0.53 - 0.61) | 0.87 (0.72 - 1.06) | 1.29 (1.07 - 1.55) | Ref |  |  |  |  | Ref |
| Age and gender adjusted HR (95% CI) | 1.42 (0.96 - 2.10) | 0.39 (0.37 - 0.41) | 0.73 (0.66 - 0.79) | 1.51 (1.38 - 1.66) | Ref | 0.72 (0.62 - 0.83) | 0.46 (0.44 - 0.48) | 0.74 (0.65 - 0.85) | 1.62 (1.42 - 1.85) | Ref | 0.57 (0.53 - 0.61) | 0.56 (0.53 - 0.60) | 0.87 (0.72 - 1.05) | 1.28 (1.06 - 1.54) | Ref |  |  |  |  | Ref |
| Age, sex and ALTCA adjusted HR (95% CI) | 1.26 (0.85 - 1.87) | 0.47 (0.45 - 0.49) | 0.76 (0.70 - 0.84) | 1.29 (1.18 - 1.41) | Ref | 0.74 (0.64 - 0.85) | 0.55 (0.53 - 0.58) | 0.73 (0.64 - 0.83) | 1.43 (1.25 - 1.63) | Ref | 0.64 (0.59 - 0.68) | 0.66 (0.62 - 0.70) | 0.84 (0.70 - 1.02) | 1.14 (0.95 - 1.37) | Ref |  |  |  |  | Ref |
| Age, sex, ALTCA and education adjusted HR (95% CI) | 1.31 (0.88 - 1.94) | 0.47 (0.45 - 0.49) | 0.76 (0.69 - 0.83) | 1.27 (1.16 - 1.39) | Ref | 0.75 (0.65 - 0.86) | 0.55 (0.53 - 0.58) | 0.73 (0.64 - 0.84) | 1.39 (1.22 - 1.60) | Ref | 0.64 (0.59 - 0.68) | 0.66 (0.61 - 0.70) | 0.84 (0.70 - 1.02) | 1.11 (0.92 - 1.34) | Ref |  |  |  |  | Ref |

| **Table S14:** Hazard ratios (HR) with 95% confidence intervals (95% CI) for all-cause mortality according to number of SARS-CoV-2 vaccine doses during different time periods of high and low COVID-19 disease burden. | | | | | | | | | | |
| --- | --- | --- | --- | --- | --- | --- | --- | --- | --- | --- |
|  | June and July 2021 (low COVID-19 disease burden) | | | | | October and November 2021 (high COVID-19 disease burden) | | | | |
|  | Four vaccine doses | Three vaccine doses | Two vaccine doses | One vaccine dose | Unvaccinated | Four vaccine doses | Three vaccine doses | Two vaccine doses | One vaccine dose | Unvaccinated |
|  | All-cause mortality | | | | | All-cause mortality | | | | |
| Deaths (n) |  | 1 | 6167 | 1597 | 4530 |  | 1788 | 7424 | 686 | 3509 |
| Events per 100.000 person days |  |  | 4.51 | 1.53 | 2.66 |  | 12.68 | 2.77 | 2.88 | 3.64 |
| Crude HR (95%CI) |  |  | 1.80 (1.73 - 1.87) | 0.58 (0.55 - 0.61) | Ref |  | 3.57 (3.35 - 3.79) | 0.76 (0.73 - 0.79) | 0.79 (0.72 - 0.85) | Ref |
| Age adjusted HR (95% CI) |  |  | 0.48 (0.46 - 0.50) | 0.55 (0.52 - 0.59) | Ref |  | 0.58 (0.54 - 0.62) | 0.51 (0.49 - 0.53) | 1.11 (1.02 - 1.21) | Ref |
| Age and gender adjusted HR (95% CI) |  |  | 0.47 (0.45 - 0.48) | 0.54 (0.51 - 0.57) | Ref |  | 0.56 (0.53 - 0.60) | 0.50 (0.48 - 0.52) | 1.09 (1.00 - 1.18) | Ref |
| Age, sex and ALTCA adjusted HR (95% CI) |  |  | 0.52 (0.50 - 0.54) | 0.64 (0.60 - 0.68) | Ref |  | 0.53 (0.50 - 0.56) | 0.56 (0.54 - 0.59) | 0.99 (0.92 - 1.08) | Ref |
| Age, sex, ALTCA and education adjusted HR (95% CI) |  |  | 0.52 (0.50 - 0.54) | 0.64 (0.61 - 0.68) | Ref |  | 0.54 (0.51 - 0.57) | 0.57 (0.55 - 0.59) | 0.99 (0.92 - 1.08) | Ref |
|  | May and June 2022 (low COVID-19 disease burden) | | | | | February and March 2022 (high COVID-19 disease burden) | | | | |
|  | Four vaccine doses | Three vaccine doses | Two vaccine doses | One vaccine dose | Unvaccinated | Four vaccine doses | Three vaccine doses | Two vaccine doses | One vaccine dose | Unvaccinated |
|  | All-cause mortality | | | | | All-cause mortality | | | | |
| Deaths (n) | 143 | 6515 | 1222 | 139 | 1311 | 18 | 7151 | 1870 | 303 | 1912 |
| Events per 100.000 person days | 9.77 | 3.39 | 3.48 | 4.55 | 3.72 | 9.34 | 3.32 | 3.52 | 6.5 | 4.71 |
| Crude HR (95%CI) | 2.60 (2.19 - 3.10) | 0.91 (0.86 - 0.97) | 0.94 (0.87 - 1.01) | 1.23 (1.03 - 1.46) | Ref | 1.96 (1.23 - 3.12) | 0.71 (0.67 - 0.74) | 0.75 (0.70 - 0.80) | 1.39 (1.23 - 1.57) | Ref |
| Age adjusted HR (95% CI) | 0.62 (0.52 - 0.74) | 0.62 (0.58 - 0.66) | 1.27 (1.17 - 1.37) | 1.60 (1.34 - 1.91) | Ref | 0.87 (0.54 - 1.38) | 0.48 (0.46 - 0.51) | 1.12 (1.05 - 1.20) | 1.50 (1.32 - 1.69) | Ref |
| Age and gender adjusted HR (95% CI) | 0.59 (0.49 - 0.70) | 0.60 (0.57 - 0.64) | 1.26 (1.16 - 1.36) | 1.59 (1.34 - 1.89) | Ref | 0.80 (0.50 - 1.28) | 0.47 (0.44 - 0.49) | 1.11 (1.04 - 1.18) | 1.49 (1.32 - 1.68) | Ref |
| Age, sex and ALTCA adjusted HR (95% CI) | 0.57 (0.48 - 0.68) | 0.60 (0.57 - 0.64) | 1.06 (0.98 - 1.15) | 1.39 (1.16 - 1.65) | Ref | 0.70 (0.44 - 1.12) | 0.47 (0.45 - 0.50) | 0.95 (0.89 - 1.02) | 1.24 (1.10 - 1.41) | Ref |
| Age, sex, ALTCA and education adjusted HR (95% CI) | 0.59 (0.50 - 0.71) | 0.61 (0.57 - 0.64) | 1.06 (0.98 - 1.15) | 1.37 (1.15 - 1.63) | Ref | 0.73 (0.46 - 1.17) | 0.48 (0.45 - 0.50) | 0.95 (0.89 - 1.01) | 1.24 (1.10 - 1.40) | Ref |

| **Table S15:** Hazard ratios (HR) with 95% confidence intervals (95% CI) for cancer and external cause mortality according to number of SARS-CoV-2 vaccine doses during different time periods of high and low COVID-19 disease burden. | | | | | | | | | | |
| --- | --- | --- | --- | --- | --- | --- | --- | --- | --- | --- |
|  | June and July 2021 (low COVID-19 disease burden) | | | | | October and November 2021 (high COVID-19 disease burden) | | | | |
|  | Four vaccine doses | Three vaccine doses | Two vaccine doses | One vaccine dose | Unvaccinated | Four vaccine doses | Three vaccine doses | Two vaccine doses | One vaccine dose | Unvaccinated |
|  | Cancer | | | | | Cancer | | | | |
| Deaths (n) |  |  | 1554 | 463 | 1149 |  | 269 | 2095 | 167 | 743 |
| Events per 100.000 person days |  |  | 1.14 | 0.44 | 0.67 |  | 1.91 | 0.78 | 0.7 | 0.77 |
| Crude HR (95%CI) |  |  | 1.76 (1.63 - 1.91) | 0.66 (0.59 - 0.74) | Ref |  | 2.55 (2.19 - 2.96) | 1.01 (0.93 - 1.10) | 0.91 (0.77 - 1.08) | Ref |
| Age adjusted HR (95% CI) |  |  | 0.55 (0.51 - 0.60) | 0.60 (0.53 - 0.66) | Ref |  | 0.56 (0.48 - 0.65) | 0.68 (0.62 - 0.74) | 1.21 (1.02 - 1.43) | Ref |
| Age and gender adjusted HR (95% CI) |  |  | 0.54 (0.50 - 0.58) | 0.58 (0.52 - 0.65) | Ref |  | 0.54 (0.46 - 0.63) | 0.66 (0.61 - 0.72) | 1.18 (1.00 - 1.40) | Ref |
| Age, sex and ALTCA adjusted HR (95% CI) |  |  | 0.60 (0.55 - 0.65) | 0.73 (0.66 - 0.82) | Ref |  | 0.47 (0.41 - 0.55) | 0.77 (0.71 - 0.84) | 1.06 (0.89 - 1.25) | Ref |
| Age, sex, ALTCA and education adjusted HR (95% CI) |  |  | 0.59 (0.55 - 0.64) | 0.73 (0.65 - 0.81) | Ref |  | 0.47 (0.41 - 0.55) | 0.77 (0.71 - 0.84) | 1.06 (0.90 - 1.26) | Ref |
|  | External cause | | | | | External cause | | | | |
| Deaths (n) |  |  | 317 | 121 | 304 |  | 79 | 440 | 42 | 189 |
| Events per 100.000 person days |  |  | 0.23 | 0.12 | 0.18 |  | 0.56 | 0.16 | 0.18 | 0.20 |
| Crude HR (95%CI) |  |  | 1.31 (1.11 - 1.54) | 0.65 (0.53 - 0.81) | Ref |  | 2.76 (2.08 - 3.66) | 0.84 (0.71 - 0.99) | 0.90 (0.64 - 1.25) | Ref |
| Age adjusted HR (95% CI) |  |  | 0.53 (0.44 - 0.62) | 0.53 (0.43 - 0.66) | Ref |  | 0.66 (0.49 - 0.89) | 0.56 (0.48 - 0.67) | 1.12 (0.80 - 1.57) | Ref |
| Age and gender adjusted HR (95% CI) |  |  | 0.51 (0.43 - 0.61) | 0.51 (0.41 - 0.63) | Ref |  | 0.63 (0.47 - 0.85) | 0.54 (0.46 - 0.65) | 1.07 (0.76 - 1.49) | Ref |
| Age, sex and ALTCA adjusted HR (95% CI) |  |  | 0.55 (0.46 - 0.65) | 0.59 (0.48 - 0.73) | Ref |  | 0.58 (0.43 - 0.78) | 0.61 (0.51 - 0.72) | 1.00 (0.72 - 1.41) | Ref |
| Age, sex, ALTCA and education adjusted HR (95% CI) |  |  | 0.56 (0.47 - 0.67) | 0.61 (0.49 - 0.75) | Ref |  | 0.59 (0.44 - 0.79) | 0.61 (0.52 - 0.73) | 1.00 (0.72 - 1.41) | Ref |
|  | May and June 2022 (low COVID-19 disease burden) | | | | | February and March 2022 (high COVID-19 disease burden) | | | | |
|  | Four vaccine doses | Three vaccine doses | Two vaccine doses | One vaccine dose | Unvaccinated | Four vaccine doses | Three vaccine doses | Two vaccine doses | One vaccine dose | Unvaccinated |
|  | Cancer | | | | | Cancer | | | | |
| Deaths (n) | 32 | 1725 | 281 | 37 | 242 | 12 | 1763 | 479 | 74 | 346 |
| Events per 100.000 person days | 2.19 | 0.9 | 0.8 | 1.21 | 0.69 | 6.22 | 0.82 | 0.9 | 1.59 | 0.85 |
| Crude HR (95%CI) | 3.27 (2.25 - 4.75) | 1.31 (1.14 - 1.50) | 1.16 (0.98 - 1.38) | 1.77 (1.25 - 2.50) | Ref | 7.29 (4.10 - 12.98) | 0.96 (0.86 - 1.08) | 1.06 (0.92 - 1.21) | 1.86 (1.45 - 2.40) | Ref |
| Age adjusted HR (95% CI) | 0.93 (0.64 - 1.36) | 0.92 (0.80 - 1.05) | 1.54 (1.30 - 1.83) | 2.27 (1.61 - 3.21) | Ref | 3.50 (1.96 - 6.23) | 0.67 (0.59 - 0.75) | 1.53 (1.33 - 1.75) | 2.07 (1.61 - 2.67) | Ref |
| Age and gender adjusted HR (95% CI) | 0.89 (0.61 - 1.30) | 0.89 (0.78 - 1.02) | 1.53 (1.29 - 1.82) | 2.25 (1.60 - 3.19) | Ref | 3.41 (1.91 - 6.08) | 0.65 (0.58 - 0.73) | 1.51 (1.32 - 1.74) | 2.07 (1.61 - 2.66) | Ref |
| Age, sex and ALTCA adjusted HR (95% CI) | 0.82 (0.56 - 1.20) | 0.91 (0.79 - 1.04) | 1.25 (1.05 - 1.48) | 1.87 (1.32 - 2.64) | Ref | 2.58 (1.44 - 4.60) | 0.67 (0.60 - 0.76) | 1.25 (1.09 - 1.44) | 1.56 (1.22 - 2.01) | Ref |
| Age, sex, ALTCA and education adjusted HR (95% CI) | 0.81 (0.55 - 1.20) | 0.92 (0.80 - 1.05) | 1.27 (1.07 - 1.52) | 1.89 (1.34 - 2.68) | Ref | 2.54 (1.42 - 4.56) | 0.67 (0.60 - 0.76) | 1.26 (1.10 - 1.45) | 1.57 (1.22 - 2.02) | Ref |
|  | External cause | | | | | External cause | | | | |
| Deaths (n) | 6 | 457 | 103 | 7 | 77 |  | 438 | 126 | 17 | 99 |
| Events per 100.000 person days |  | 0.24 | 0.29 |  | 0.22 |  | 0.2 | 0.24 | 0.36 | 0.24 |
| Crude HR (95%CI) |  | 1.09 (0.86 - 1.39) | 1.35 (1.00 - 1.81) |  | Ref |  | 0.83 (0.67 - 1.04) | 0.98 (0.75 - 1.27) | 1.50 (0.90 - 2.51) | Ref |
| Age adjusted HR (95% CI) |  | 0.80 (0.63 - 1.02) | 1.66 (1.23 - 2.23) |  | Ref |  | 0.60 (0.48 - 0.75) | 1.21 (0.93 - 1.58) | 1.68 (1.00 - 2.81) | Ref |
| Age and gender adjusted HR (95% CI) |  | 0.77 (0.60 - 0.98) | 1.61 (1.19 - 2.16) |  | Ref |  | 0.59 (0.47 - 0.73) | 1.19 (0.91 - 1.55) | 1.66 (0.99 - 2.79) | Ref |
| Age, sex and ALTCA adjusted HR (95% CI) |  | 0.78 (0.61 - 1.00) | 1.46 (1.09 - 1.97) |  | Ref |  | 0.61 (0.49 - 0.76) | 1.08 (0.83 - 1.41) | 1.40 (0.83 - 2.35) | Ref |
| Age, sex, ALTCA and education adjusted HR (95% CI) |  | 0.80 (0.62 - 1.02) | 1.43 (1.06 - 1.94) |  | Ref |  | 0.62 (0.50 - 0.78) | 1.06 (0.81 - 1.38) | 1.38 (0.82 - 2.32) | Ref |
| Estimates with less than 10 deaths in either the vaccinated or reference group are not shown | | | | | | | | | | |

### **Sensitivity Analyses Tables (S16 – S47)**

| **Table S16:** Hazard ratios (HR) with 95% confidence intervals (95% CI) for all-cause mortality according to number of SARS-CoV-2 vaccine doses for the period from 2021 to 2022 split into 3 month intervals for males only. | | | | | | | | | | | | | | | | | | | | |
| --- | --- | --- | --- | --- | --- | --- | --- | --- | --- | --- | --- | --- | --- | --- | --- | --- | --- | --- | --- | --- |
|  | 2021 | | | | | | | | | | | | | | | | | | | |
|  | All-cause mortality | | | | | | | | | | | | | | | | | | | |
|  | first quarter | | | | | second quarter | | | | | third quarter | | | | | fourth quarter | | | | |
|  | Four vaccine doses | Three vaccine doses | Two vaccine doses | One vaccine dose | Unvaccinated | Four vaccine doses | Three vaccine doses | Two vaccine doses | One vaccine dose | Unvaccinated | Four vaccine doses | Three vaccine doses | Two vaccine doses | One vaccine dose | Unvaccinated | Four vaccine doses | Three vaccine doses | Two vaccine doses | One vaccine dose | Unvaccinated |
| Deaths (n) |  |  | 503 | 586 | 8020 |  |  | 3316 | 1417 | 4597 |  | 31 | 6017 | 734 | 2803 | 1 | 2502 | 4945 | 486 | 2260 |
| Events per 100,000 person days |  |  | 10.25 | 6.5 | 2.59 |  |  | 6.43 | 2.04 | 2.32 |  | 8.84 | 3.24 | 2.37 | 2.74 |  | 4.37 | 3.11 | 2.59 | 3.11 |
| Crude HR (95%CI) |  |  | 4.26 (3.88 - 4.67) | 2.75 (2.52 - 3.00) | Ref |  |  | 2.98 (2.84 - 3.13) | 0.89 (0.83 - 0.94) | Ref |  | 3.27 (2.28 - 4.68) | 1.19 (1.14 - 1.24) | 0.88 (0.81 - 0.96) | Ref |  | 1.75 (1.64 - 1.88) | 1.00 (0.95 - 1.05) | 0.82 (0.75 - 0.91) | Ref |
| Age adjusted HR (95% CI) |  |  | 0.94 (0.85 - 1.04) | 0.71 (0.65 - 0.77) | Ref |  |  | 0.43 (0.41 - 0.45) | 0.41 (0.38 - 0.43) | Ref |  | 0.44 (0.30 - 0.63) | 0.45 (0.43 - 0.47) | 1.10 (1.02 - 1.20) | Ref |  | 0.35 (0.33 - 0.37) | 0.69 (0.65 - 0.72) | 1.01 (0.91 - 1.11) | Ref |
| Age and ALTCA adjusted HR (95% CI) |  |  | 0.69 (0.63 - 0.76) | 0.62 (0.57 - 0.68) | Ref |  |  | 0.45 (0.43 - 0.47) | 0.50 (0.47 - 0.53) | Ref |  | 0.37 (0.26 - 0.53) | 0.52 (0.50 - 0.55) | 1.05 (0.97 - 1.14) | Ref |  | 0.38 (0.36 - 0.41) | 0.75 (0.71 - 0.79) | 0.92 (0.83 - 1.01) | Ref |
| Age, ALTCA and education adjusted HR (95% CI) |  |  | 0.70 (0.64 - 0.77) | 0.62 (0.57 - 0.68) | Ref |  |  | 0.45 (0.43 - 0.47) | 0.50 (0.47 - 0.53) | Ref |  | 0.37 (0.26 - 0.54) | 0.52 (0.50 - 0.54) | 1.05 (0.96 - 1.14) | Ref |  | 0.38 (0.36 - 0.41) | 0.74 (0.70 - 0.78) | 0.89 (0.81 - 0.99) | Ref |
|  | 2022 | | | | | | | | | | | | | | | | | | | |
| Deaths (n) | 13 | 5733 | 1641 | 259 | 1410 | 99 | 5196 | 945 | 116 | 928 | 466 | 3189 | 520 | 59 | 608 |  |  |  |  | 0 |
| Events per 100,000 person days | 8.06 | 3.55 | 3.33 | 5.74 | 3.17 | 8.11 | 3.67 | 3.21 | 4.31 | 2.66 | 6.57 | 3.9 | 3.08 | 3.46 | 2.73 |  |  |  |  |  |
| Crude HR (95%CI) | 2.52 (1.46 - 4.35) | 1.12 (1.05 - 1.18) | 1.06 (0.99 - 1.14) | 1.82 (1.60 - 2.08) | Ref | 3.12 (2.53 - 3.86) | 1.38 (1.28 - 1.48) | 1.20 (1.10 - 1.32) | 1.62 (1.33 - 1.96) | Ref | 2.37 (2.10 - 2.68) | 1.43 (1.31 - 1.56) | 1.14 (1.01 - 1.28) | 1.22 (0.93 - 1.59) | Ref |  |  |  |  | Ref |
| Age adjusted HR (95% CI) | 0.56 (0.32 - 0.97) | 0.45 (0.43 - 0.48) | 1.20 (1.12 - 1.29) | 1.60 (1.40 - 1.83) | Ref | 0.43 (0.34 - 0.53) | 0.61 (0.57 - 0.65) | 1.28 (1.17 - 1.40) | 1.76 (1.45 - 2.14) | Ref | 0.40 (0.35 - 0.45) | 0.71 (0.65 - 0.77) | 1.26 (1.12 - 1.41) | 1.36 (1.04 - 1.78) | Ref |  |  |  |  | Ref |
| Age and ALTCA adjusted HR (95% CI) | 0.50 (0.29 - 0.87) | 0.47 (0.45 - 0.50) | 1.03 (0.96 - 1.11) | 1.30 (1.14 - 1.49) | Ref | 0.41 (0.33 - 0.52) | 0.62 (0.58 - 0.67) | 1.07 (0.97 - 1.17) | 1.48 (1.22 - 1.79) | Ref | 0.41 (0.36 - 0.47) | 0.70 (0.64 - 0.76) | 1.05 (0.94 - 1.18) | 1.14 (0.87 - 1.49) | Ref |  |  |  |  | Ref |
| Age, ALTCA and education adjusted HR (95% CI) | 0.53 (0.31 - 0.92) | 0.47 (0.44 - 0.49) | 1.00 (0.93 - 1.07) | 1.28 (1.12 - 1.46) | Ref | 0.43 (0.34 - 0.53) | 0.60 (0.56 - 0.65) | 1.03 (0.94 - 1.13) | 1.43 (1.18 - 1.74) | Ref | 0.43 (0.37 - 0.49) | 0.68 (0.62 - 0.74) | 1.02 (0.90 - 1.15) | 1.09 (0.83 - 1.43) | Ref |  |  |  |  | Ref |
| Estimates with less than 10 deaths in either the vaccinated or reference group are not shown | | | | | | | | | | | | | | | | | | | | |

| **Table S17:** Hazard ratios (HR) with 95% confidence intervals (95% CI) for all-cause mortality according to number of SARS-CoV-2 vaccine doses for the period from 2021 to 2022 split into 3 month intervals for females only. | | | | | | | | | | | | | | | | | | | | |
| --- | --- | --- | --- | --- | --- | --- | --- | --- | --- | --- | --- | --- | --- | --- | --- | --- | --- | --- | --- | --- |
|  | 2021 | | | | | | | | | | | | | | | | | | | |
|  | All-cause mortality | | | | | | | | | | | | | | | | | | | |
|  | first quarter | | | | | second quarter | | | | | third quarter | | | | | fourth quarter | | | | |
|  | Four vaccine doses | Three vaccine doses | Two vaccine doses | One vaccine dose | Unvaccinated | Four vaccine doses | Three vaccine doses | Two vaccine doses | One vaccine dose | Unvaccinated | Four vaccine doses | Three vaccine doses | Two vaccine doses | One vaccine dose | Unvaccinated | Four vaccine doses | Three vaccine doses | Two vaccine doses | One vaccine dose | Unvaccinated |
| Deaths (n) |  |  | 871 | 785 | 7563 |  | 1 | 3594 | 1129 | 4226 |  | 69 | 5566 | 654 | 3039 |  | 2866 | 4223 | 488 | 2753 |
| Events per 100,000 person days |  |  | 9.69 | 5.74 | 2.41 |  |  | 5.33 | 1.55 | 2.20 |  | 15.58 | 2.74 | 2.51 | 2.93 |  | 4.25 | 2.58 | 3.31 | 3.65 |
| Crude HR (95%CI) |  |  | 4.46 (4.14 - 4.80) | 2.70 (2.50 - 2.92) | Ref |  |  | 2.57 (2.45 - 2.70) | 0.69 (0.65 - 0.74) | Ref |  | 4.96 (3.87 - 6.35) | 0.93 (0.89 - 0.98) | 0.90 (0.83 - 0.98) | Ref |  | 1.38 (1.29 - 1.46) | 0.72 (0.69 - 0.76) | 0.90 (0.82 - 0.99) | Ref |
| Age adjusted HR (95% CI) |  |  | 0.98 (0.91 - 1.06) | 0.79 (0.73 - 0.85) | Ref |  |  | 0.47 (0.45 - 0.50) | 0.46 (0.43 - 0.50) | Ref |  | 0.74 (0.58 - 0.96) | 0.47 (0.45 - 0.49) | 1.21 (1.11 - 1.32) | Ref |  | 0.38 (0.36 - 0.40) | 0.68 (0.65 - 0.71) | 1.01 (0.92 - 1.12) | Ref |
| Age and ALTCA adjusted HR (95% CI) |  |  | 0.79 (0.73 - 0.85) | 0.70 (0.65 - 0.75) | Ref |  |  | 0.47 (0.45 - 0.49) | 0.55 (0.51 - 0.59) | Ref |  | 0.62 (0.48 - 0.79) | 0.51 (0.48 - 0.53) | 1.15 (1.05 - 1.25) | Ref |  | 0.39 (0.37 - 0.41) | 0.72 (0.68 - 0.75) | 0.94 (0.86 - 1.04) | Ref |
| Age, ALTCA and education adjusted HR (95% CI) |  |  | 0.79 (0.73 - 0.85) | 0.70 (0.65 - 0.75) | Ref |  |  | 0.47 (0.45 - 0.49) | 0.54 (0.51 - 0.58) | Ref |  | 0.61 (0.48 - 0.79) | 0.50 (0.48 - 0.53) | 1.13 (1.04 - 1.24) | Ref |  | 0.39 (0.36 - 0.41) | 0.71 (0.67 - 0.74) | ( - ) | Ref |
|  | 2022 | | | | | | | | | | | | | | | | | | | |
| Deaths (n) | 12 | 5328 | 1558 | 308 | 1764 | 100 | 4728 | 1008 | 122 | 1186 | 465 | 2872 | 482 | 63 | 785 |  |  |  |  | 0 |
| Events per 100,000 person days | 8.59 | 3.03 | 3.53 | 7.49 | 3.87 | 7.56 | 3.12 | 4.16 | 5.54 | 3.48 | 5.57 | 3.34 | 3.65 | 4.57 | 3.64 |  |  |  |  |  |
| Crude HR (95%CI) | 2.16 (1.22 - 3.81) | 0.78 (0.74 - 0.82) | 0.92 (0.86 - 0.99) | 1.95 (1.73 - 2.21) | Ref | 2.23 (1.81 - 2.75) | 0.90 (0.84 - 0.96) | 1.19 (1.10 - 1.30) | 1.59 (1.32 - 1.92) | Ref | 1.50 (1.34 - 1.69) | 0.92 (0.85 - 0.99) | 1.01 (0.90 - 1.13) | 1.22 (0.94 - 1.58) | Ref |  |  |  |  | Ref |
| Age adjusted HR (95% CI) | 0.83 (0.47 - 1.46) | 0.44 (0.41 - 0.46) | 1.05 (0.98 - 1.12) | 1.44 (1.28 - 1.63) | Ref | 0.44 (0.36 - 0.55) | 0.53 (0.50 - 0.56) | 1.15 (1.06 - 1.25) | 1.50 (1.24 - 1.80) | Ref | 0.36 (0.32 - 0.40) | 0.58 (0.54 - 0.63) | 1.00 (0.89 - 1.12) | 1.20 (0.93 - 1.55) | Ref |  |  |  |  | Ref |
| Age and ALTCA adjusted HR (95% CI) | ( - ) | 0.44 (0.42 - 0.46) | 0.94 (0.88 - 1.01) | 1.27 (1.13 - 1.44) | Ref | 0.44 (0.36 - 0.54) | 0.53 (0.50 - 0.56) | 1.02 (0.93 - 1.11) | 1.38 (1.15 - 1.66) | Ref | 0.38 (0.33 - 0.42) | 0.57 (0.53 - 0.62) | 0.88 (0.78 - 0.98) | 1.13 (0.88 - 1.47) | Ref |  |  |  |  | Ref |
| Age, ALTCA and education adjusted HR (95% CI) | ( - ) | 0.43 (0.41 - 0.46) | 0.93 (0.87 - 0.99) | 1.26 (1.12 - 1.43) | Ref | 0.46 (0.37 - 0.57) | 0.53 (0.50 - 0.56) | 1.01 (0.93 - 1.10) | 1.36 (1.13 - 1.64) | Ref | 0.38 (0.33 - 0.42) | 0.57 (0.52 - 0.61) | 0.87 (0.78 - 0.97) | 1.13 (0.88 - 1.46) | Ref |  |  |  |  | Ref |
| Estimates with less than 10 deaths in either the vaccinated or reference group are not shown | | | | | | | | | | | | | | | | | | | | |

| **Table S18:** Hazard ratios (HR) with 95% confidence intervals (95% CI) for all-cause mortality according to number of SARS-CoV-2 vaccine doses for the period from 2021 to 2022 split into 3 month intervals for 18-39 year olds only. | | | | | | | | | | | | | | | | | | | | |
| --- | --- | --- | --- | --- | --- | --- | --- | --- | --- | --- | --- | --- | --- | --- | --- | --- | --- | --- | --- | --- |
|  | 2021 | | | | | | | | | | | | | | | | | | | |
|  | All-cause mortality | | | | | | | | | | | | | | | | | | | |
|  | first quarter | | | | | second quarter | | | | | third quarter | | | | | fourth quarter | | | | |
|  | Four vaccine doses | Three vaccine doses | Two vaccine doses | One vaccine dose | Unvaccinated | Four vaccine doses | Three vaccine doses | Two vaccine doses | One vaccine dose | Unvaccinated | Four vaccine doses | Three vaccine doses | Two vaccine doses | One vaccine dose | Unvaccinated | Four vaccine doses | Three vaccine doses | Two vaccine doses | One vaccine dose | Unvaccinated |
| Deaths (n) |  |  | 4 | 5 | 275 |  |  | 19 | 30 | 251 |  |  | 95 | 43 | 157 |  | 15 | 117 | 31 | 106 |
| Events per 100,000 person days |  |  |  |  | 0.12 |  |  | 0.11 | 0.09 | 0.14 |  |  | 0.1 | 0.15 | 0.15 |  | 0.07 | 0.11 | 0.18 | 0.15 |
| Crude HR (95%CI) |  |  |  |  | Ref |  |  | 0.73 (0.46 - 1.18) | 0.59 (0.40 - 0.87) | Ref |  |  | 0.67 (0.52 - 0.87) | 0.99 (0.70 - 1.39) | Ref |  | 0.49 (0.27 - 0.89) | 0.74 (0.57 - 0.96) | 1.19 (0.79 - 1.77) | Ref |
| Age adjusted HR (95% CI) |  |  |  |  | Ref |  |  | 0.69 (0.43 - 1.11) | 0.56 (0.38 - 0.82) | Ref |  |  | 0.64 (0.49 - 0.83) | 0.99 (0.70 - 1.40) | Ref |  | 0.44 (0.24 - 0.79) | 0.71 (0.55 - 0.93) | 1.22 (0.82 - 1.82) | Ref |
| Age and gender adjusted HR (95% CI) |  |  |  |  | Ref |  |  | 0.75 (0.46 - 1.20) | 0.56 (0.38 - 0.83) | Ref |  |  | 0.65 (0.50 - 0.84) | 0.96 (0.68 - 1.35) | Ref |  | 0.45 (0.25 - 0.82) | 0.71 (0.55 - 0.93) | 1.16 (0.78 - 1.74) | Ref |
| Age, sex and ALTCA adjusted HR (95% CI) |  |  |  |  | Ref |  |  | 0.40 (0.24 - 0.65) | 0.53 (0.36 - 0.77) | Ref |  |  | 0.56 (0.43 - 0.73) | 0.99 (0.70 - 1.39) | Ref |  | 0.32 (0.18 - 0.59) | 0.67 (0.51 - 0.87) | 1.17 (0.78 - 1.75) | Ref |
| Age, sex, ALTCA and education adjusted HR (95% CI) |  |  |  |  | Ref |  |  | 0.46 (0.28 - 0.77) | 0.61 (0.41 - 0.91) | Ref |  |  | 0.60 (0.46 - 0.79) | 1.02 (0.72 - 1.44) | Ref |  | 0.34 (0.19 - 0.63) | 0.69 (0.53 - 0.90) | 1.10 (0.73 - 1.64) | Ref |
|  | 2022 | | | | | | | | | | | | | | | | | | | |
| Deaths (n) | 1 | 78 | 100 | 19 | 71 | 1 | 107 | 59 | 7 | 50 |  | 61 | 35 | 5 | 38 |  |  |  |  | 0 |
| Events per 100,000 person days |  | 0.1 | 0.22 | 0.44 | 0.17 |  | 0.17 | 0.23 |  | 0.16 |  | 0.16 | 0.24 |  | 0.19 |  |  |  |  |  |
| Crude HR (95%CI) |  | 0.57 (0.41 - 0.78) | 1.29 (0.95 - 1.75) | 2.57 (1.54 - 4.26) | Ref |  | 1.03 (0.74 - 1.44) | 1.45 (1.00 - 2.12) |  | Ref |  | 0.85 (0.57 - 1.28) | 1.30 (0.82 - 2.06) |  | Ref |  |  |  |  | Ref |
| Age adjusted HR (95% CI) |  | 0.53 (0.38 - 0.73) | 1.29 (0.95 - 1.75) | 2.65 (1.59 - 4.40) | Ref |  | 0.94 (0.67 - 1.32) | 1.44 (0.99 - 2.10) |  | Ref |  | 0.81 (0.54 - 1.22) | 1.29 (0.81 - 2.05) |  | Ref |  |  |  |  | Ref |
| Age and gender adjusted HR (95% CI) |  | 0.53 (0.39 - 0.74) | 1.27 (0.94 - 1.72) | 2.59 (1.56 - 4.31) | Ref |  | 0.95 (0.68 - 1.33) | 1.39 (0.95 - 2.02) |  | Ref |  | 0.82 (0.54 - 1.24) | 1.25 (0.79 - 1.99) |  | Ref |  |  |  |  | Ref |
| Age, sex and ALTCA adjusted HR (95% CI) |  | 0.47 (0.34 - 0.65) | 1.27 (0.93 - 1.72) | 2.44 (1.47 - 4.06) | Ref |  | 0.81 (0.58 - 1.14) | 1.35 (0.93 - 1.97) |  | Ref |  | 0.71 (0.47 - 1.08) | 1.23 (0.77 - 1.95) |  | Ref |  |  |  |  | Ref |
| Age, sex, ALTCA and education adjusted HR (95% CI) |  | 0.52 (0.37 - 0.73) | 1.15 (0.84 - 1.56) | 2.18 (1.31 - 3.64) | Ref |  | 0.79 (0.56 - 1.12) | 1.15 (0.78 - 1.69) |  | Ref |  | 0.68 (0.44 - 1.05) | 1.09 (0.67 - 1.76) |  | Ref |  |  |  |  | Ref |
| Estimates with less than 10 deaths in either the vaccinated or reference group are not shown | | | | | | | | | | | | | | | | | | | | |

| **Table S19:** Hazard ratios (HR) with 95% confidence intervals (95% CI) for all-cause mortality according to number of SARS-CoV-2 vaccine doses for the period from 2021 to 2022 split into 3 month intervals for 40-59 year olds only. | | | | | | | | | | | | | | | | | | | | |
| --- | --- | --- | --- | --- | --- | --- | --- | --- | --- | --- | --- | --- | --- | --- | --- | --- | --- | --- | --- | --- |
|  | 2021 | | | | | | | | | | | | | | | | | | | |
|  | All-cause mortality | | | | | | | | | | | | | | | | | | | |
|  | first quarter | | | | | second quarter | | | | | third quarter | | | | | fourth quarter | | | | |
|  | Four vaccine doses | Three vaccine doses | Two vaccine doses | One vaccine dose | Unvaccinated | Four vaccine doses | Three vaccine doses | Two vaccine doses | One vaccine dose | Unvaccinated | Four vaccine doses | Three vaccine doses | Two vaccine doses | One vaccine dose | Unvaccinated | Four vaccine doses | Three vaccine doses | Two vaccine doses | One vaccine dose | Unvaccinated |
| Deaths (n) |  |  | 19 | 27 | 1390 |  |  | 244 | 215 | 1060 |  | 3 | 795 | 159 | 689 |  | 189 | 801 | 109 | 493 |
| Events per 100,000 person days |  |  | 0.4 | 0.38 | 0.65 |  |  | 0.8 | 0.41 | 0.76 |  |  | 0.58 | 0.79 | 1.03 |  | 0.5 | 0.68 | 0.99 | 1.02 |
| Crude HR (95%CI) |  |  | 0.61 (0.39 - 0.96) | 0.57 (0.39 - 0.84) | Ref |  |  | 0.98 (0.84 - 1.14) | 0.50 (0.43 - 0.58) | Ref |  |  | 0.57 (0.51 - 0.63) | 0.79 (0.67 - 0.95) | Ref |  | 0.48 (0.39 - 0.58) | 0.68 (0.61 - 0.76) | 0.96 (0.78 - 1.19) | Ref |
| Age adjusted HR (95% CI) |  |  | 0.59 (0.37 - 0.92) | 0.55 (0.37 - 0.80) | Ref |  |  | 0.83 (0.71 - 0.96) | 0.43 (0.37 - 0.50) | Ref |  |  | 0.49 (0.44 - 0.55) | 0.80 (0.67 - 0.95) | Ref |  | 0.40 (0.33 - 0.49) | 0.63 (0.56 - 0.70) | 0.98 (0.80 - 1.21) | Ref |
| Age and gender adjusted HR (95% CI) |  |  | 0.67 (0.43 - 1.06) | 0.61 (0.41 - 0.89) | Ref |  |  | 0.88 (0.76 - 1.03) | 0.43 (0.37 - 0.51) | Ref |  |  | 0.50 (0.45 - 0.55) | 0.77 (0.65 - 0.92) | Ref |  | 0.41 (0.34 - 0.50) | 0.62 (0.55 - 0.69) | 0.93 (0.76 - 1.15) | Ref |
| Age, sex and ALTCA adjusted HR (95% CI) |  |  | 0.50 (0.32 - 0.79) | 0.43 (0.29 - 0.63) | Ref |  |  | 0.61 (0.53 - 0.71) | 0.45 (0.39 - 0.53) | Ref |  |  | 0.50 (0.45 - 0.55) | 0.81 (0.68 - 0.96) | Ref |  | 0.37 (0.31 - 0.45) | 0.65 (0.58 - 0.72) | 0.93 (0.75 - 1.14) | Ref |
| Age, sex, ALTCA and education adjusted HR (95% CI) |  |  | 0.52 (0.33 - 0.82) | 0.44 (0.30 - 0.64) | Ref |  |  | 0.62 (0.53 - 0.72) | 0.45 (0.39 - 0.53) | Ref |  |  | 0.51 (0.46 - 0.56) | 0.80 (0.67 - 0.96) | Ref |  | 0.38 (0.31 - 0.46) | 0.64 (0.57 - 0.71) | 0.88 (0.71 - 1.08) | Ref |
|  | 2022 | | | | | | | | | | | | | | | | | | | |
| Deaths (n) | 3 | 612 | 335 | 60 | 318 | 8 | 726 | 218 | 28 | 212 | 18 | 476 | 136 | 15 | 136 |  |  |  |  | 0 |
| Events per 100,000 person days |  | 0.53 | 1.07 | 2.51 | 1.09 |  | 0.74 | 1.26 | 2.07 | 1.00 | 1.06 | 0.81 | 1.4 | 1.75 | 1.01 |  |  |  |  |  |
| Crude HR (95%CI) |  | 0.48 (0.42 - 0.55) | 0.99 (0.85 - 1.15) | 2.29 (1.74 - 3.02) | Ref |  | 0.74 (0.63 - 0.86) | 1.25 (1.04 - 1.51) | 2.07 (1.39 - 3.06) | Ref | 1.00 (0.61 - 1.65) | 0.80 (0.66 - 0.97) | 1.37 (1.08 - 1.73) | 1.74 (1.02 - 2.96) | Ref |  |  |  |  | Ref |
| Age adjusted HR (95% CI) |  | 0.43 (0.37 - 0.49) | 1.00 (0.86 - 1.17) | 2.34 (1.77 - 3.08) | Ref |  | 0.66 (0.56 - 0.77) | 1.26 (1.04 - 1.52) | 2.15 (1.45 - 3.18) | Ref | 0.76 (0.46 - 1.27) | 0.72 (0.59 - 0.87) | 1.37 (1.08 - 1.74) | 1.81 (1.06 - 3.09) | Ref |  |  |  |  | Ref |
| Age and gender adjusted HR (95% CI) |  | 0.43 (0.37 - 0.49) | 0.98 (0.84 - 1.15) | 2.31 (1.75 - 3.05) | Ref |  | 0.66 (0.57 - 0.77) | 1.23 (1.02 - 1.49) | 2.11 (1.42 - 3.13) | Ref | 0.77 (0.47 - 1.28) | 0.72 (0.60 - 0.87) | 1.32 (1.04 - 1.68) | 1.78 (1.04 - 3.03) | Ref |  |  |  |  | Ref |
| Age, sex and ALTCA adjusted HR (95% CI) |  | 0.43 (0.38 - 0.50) | 0.95 (0.81 - 1.10) | 1.88 (1.42 - 2.49) | Ref |  | 0.67 (0.57 - 0.78) | 1.17 (0.97 - 1.42) | 1.85 (1.25 - 2.75) | Ref | 0.59 (0.36 - 0.98) | 0.73 (0.60 - 0.88) | 1.30 (1.02 - 1.65) | 1.57 (0.92 - 2.68) | Ref |  |  |  |  | Ref |
| Age, sex, ALTCA and education adjusted HR (95% CI) |  | 0.42 (0.37 - 0.49) | 0.91 (0.78 - 1.07) | 1.86 (1.40 - 2.45) | Ref |  | 0.66 (0.56 - 0.77) | 1.12 (0.93 - 1.36) | 1.72 (1.15 - 2.58) | Ref | 0.58 (0.35 - 0.96) | 0.69 (0.57 - 0.84) | 1.21 (0.95 - 1.54) | 1.57 (0.92 - 2.68) | Ref |  |  |  |  | Ref |
| Estimates with less than 10 deaths in either the vaccinated or reference group are not shown | | | | | | | | | | | | | | | | | | | | |

| **Table S20:** Hazard ratios (HR) with 95% confidence intervals (95% CI) for all-cause mortality according to number of SARS-CoV-2 vaccine doses for the period from 2021 to 2022 split into 3 month intervals for 60-74 year olds only. | | | | | | | | | | | | | | | | | | | | |
| --- | --- | --- | --- | --- | --- | --- | --- | --- | --- | --- | --- | --- | --- | --- | --- | --- | --- | --- | --- | --- |
|  | 2021 | | | | | | | | | | | | | | | | | | | |
|  | All-cause mortality | | | | | | | | | | | | | | | | | | | |
|  | first quarter | | | | | second quarter | | | | | third quarter | | | | | fourth quarter | | | | |
|  | Four vaccine doses | Three vaccine doses | Two vaccine doses | One vaccine dose | Unvaccinated | Four vaccine doses | Three vaccine doses | Two vaccine doses | One vaccine dose | Unvaccinated | Four vaccine doses | Three vaccine doses | Two vaccine doses | One vaccine dose | Unvaccinated | Four vaccine doses | Three vaccine doses | Two vaccine doses | One vaccine dose | Unvaccinated |
| Deaths (n) |  |  | 139 | 148 | 3750 |  |  | 938 | 810 | 2414 |  | 12 | 2454 | 380 | 1389 | 1 | 726 | 2363 | 226 | 1194 |
| Events per 100,000 person days |  |  | 7.97 | 4.48 | 3.03 |  |  | 3.06 | 1.87 | 4.40 |  | 6.26 | 2.49 | 6.32 | 5.33 |  | 2.1 | 3.4 | 6.26 | 5.79 |
| Crude HR (95%CI) |  |  | 2.70 (2.27 - 3.21) | 1.53 (1.29 - 1.81) | Ref |  |  | 0.62 (0.57 - 0.67) | 0.38 (0.35 - 0.41) | Ref |  | 1.11 (0.62 - 1.96) | 0.46 (0.43 - 0.50) | 1.23 (1.09 - 1.38) | Ref |  | 0.40 (0.36 - 0.44) | 0.64 (0.60 - 0.69) | 1.07 (0.93 - 1.24) | Ref |
| Age adjusted HR (95% CI) |  |  | 2.69 (2.27 - 3.20) | 1.50 (1.27 - 1.78) | Ref |  |  | 0.52 (0.47 - 0.56) | 0.34 (0.31 - 0.37) | Ref |  | 0.93 (0.52 - 1.65) | 0.43 (0.40 - 0.46) | 1.26 (1.12 - 1.41) | Ref |  | 0.35 (0.31 - 0.39) | 0.62 (0.58 - 0.67) | 1.08 (0.93 - 1.24) | Ref |
| Age and gender adjusted HR (95% CI) |  |  | 2.66 (2.24 - 3.15) | 1.47 (1.24 - 1.75) | Ref |  |  | 0.51 (0.47 - 0.55) | 0.34 (0.31 - 0.37) | Ref |  | 0.87 (0.49 - 1.54) | 0.42 (0.40 - 0.45) | 1.23 (1.10 - 1.38) | Ref |  | 0.34 (0.31 - 0.38) | 0.61 (0.57 - 0.66) | 1.05 (0.91 - 1.21) | Ref |
| Age, sex and ALTCA adjusted HR (95% CI) |  |  | 1.19 (1.00 - 1.41) | 0.97 (0.82 - 1.14) | Ref |  |  | 0.49 (0.45 - 0.53) | 0.41 (0.38 - 0.44) | Ref |  | 0.50 (0.28 - 0.88) | 0.49 (0.45 - 0.52) | 1.11 (0.99 - 1.25) | Ref |  | 0.35 (0.31 - 0.39) | 0.65 (0.61 - 0.70) | 0.90 (0.78 - 1.04) | Ref |
| Age, sex, ALTCA and education adjusted HR (95% CI) |  |  | 1.21 (1.02 - 1.44) | 0.98 (0.83 - 1.16) | Ref |  |  | 0.49 (0.45 - 0.54) | 0.41 (0.37 - 0.44) | Ref |  | 0.51 (0.29 - 0.91) | 0.48 (0.45 - 0.52) | 1.10 (0.98 - 1.24) | Ref |  | 0.35 (0.31 - 0.39) | 0.64 (0.60 - 0.69) | 0.89 (0.77 - 1.02) | Ref |
|  | 2022 | | | | | | | | | | | | | | | | | | | |
| Deaths (n) | 7 | 2253 | 843 | 129 | 741 | 39 | 2272 | 478 | 64 | 512 | 146 | 1480 | 231 | 35 | 361 |  |  |  |  | 0 |
| Events per 100,000 person days |  | 2.51 | 7.33 | 10.46 | 5.10 | 5.45 | 2.79 | 6.35 | 8.81 | 4.24 | 2.58 | 3.19 | 5.54 | 7.54 | 4.67 |  |  |  |  |  |
| Crude HR (95%CI) |  | 0.49 (0.45 - 0.53) | 1.44 (1.30 - 1.59) | 2.06 (1.71 - 2.49) | Ref | 1.31 (0.94 - 1.82) | 0.66 (0.60 - 0.72) | 1.50 (1.32 - 1.70) | 2.08 (1.60 - 2.69) | Ref | 0.56 (0.46 - 0.69) | 0.70 (0.62 - 0.78) | 1.20 (1.02 - 1.41) | 1.52 (1.07 - 2.15) | Ref |  |  |  |  | Ref |
| Age adjusted HR (95% CI) |  | 0.46 (0.42 - 0.50) | 1.45 (1.31 - 1.60) | 1.99 (1.65 - 2.40) | Ref | 1.11 (0.79 - 1.55) | 0.63 (0.57 - 0.69) | 1.51 (1.33 - 1.71) | 2.02 (1.56 - 2.63) | Ref | 0.47 (0.39 - 0.58) | 0.68 (0.60 - 0.76) | 1.22 (1.03 - 1.44) | 1.49 (1.05 - 2.11) | Ref |  |  |  |  | Ref |
| Age and gender adjusted HR (95% CI) |  | 0.45 (0.41 - 0.49) | 1.43 (1.29 - 1.58) | 1.96 (1.62 - 2.36) | Ref | 1.07 (0.76 - 1.49) | 0.62 (0.56 - 0.68) | 1.49 (1.31 - 1.69) | 1.98 (1.53 - 2.57) | Ref | 0.46 (0.38 - 0.56) | 0.67 (0.60 - 0.75) | 1.20 (1.02 - 1.41) | 1.44 (1.02 - 2.04) | Ref |  |  |  |  | Ref |
| Age, sex and ALTCA adjusted HR (95% CI) |  | 0.47 (0.44 - 0.51) | 1.18 (1.07 - 1.31) | 1.52 (1.26 - 1.83) | Ref | 0.89 (0.63 - 1.23) | 0.63 (0.57 - 0.70) | 1.22 (1.07 - 1.38) | 1.67 (1.28 - 2.16) | Ref | 0.46 (0.37 - 0.56) | 0.67 (0.59 - 0.75) | 0.99 (0.84 - 1.17) | 1.12 (0.78 - 1.61) | Ref |  |  |  |  | Ref |
| Age, sex, ALTCA and education adjusted HR (95% CI) |  | 0.46 (0.43 - 0.50) | 1.15 (1.04 - 1.27) | 1.48 (1.22 - 1.79) | Ref | 0.93 (0.66 - 1.30) | 0.63 (0.57 - 0.69) | 1.19 (1.05 - 1.35) | 1.63 (1.26 - 2.12) | Ref | 0.48 (0.39 - 0.59) | 0.66 (0.59 - 0.74) | 0.98 (0.83 - 1.15) | 1.06 (0.74 - 1.53) | Ref |  |  |  |  | Ref |
| Estimates with less than 10 deaths in either the vaccinated or reference group are not shown | | | | | | | | | | | | | | | | | | | | |

| **Table S21:** Hazard ratios (HR) with 95% confidence intervals (95% CI) for all-cause mortality according to number of SARS-CoV-2 vaccine doses for the period from 2021 to 2022 split into 3 month intervals for 75-84+ year olds only. | | | | | | | | | | | | | | | | | | | | |
| --- | --- | --- | --- | --- | --- | --- | --- | --- | --- | --- | --- | --- | --- | --- | --- | --- | --- | --- | --- | --- |
|  | 2021 | | | | | | | | | | | | | | | | | | | |
|  | All-cause mortality | | | | | | | | | | | | | | | | | | | |
|  | first quarter | | | | | second quarter | | | | | third quarter | | | | | fourth quarter | | | | |
|  | Four vaccine doses | Three vaccine doses | Two vaccine doses | One vaccine dose | Unvaccinated | Four vaccine doses | Three vaccine doses | Two vaccine doses | One vaccine dose | Unvaccinated | Four vaccine doses | Three vaccine doses | Two vaccine doses | One vaccine dose | Unvaccinated | Four vaccine doses | Three vaccine doses | Two vaccine doses | One vaccine dose | Unvaccinated |
| Deaths (n) |  |  | 361 | 398 | 4549 |  |  | 2200 | 822 | 2246 |  | 19 | 3738 | 417 | 1535 |  | 1713 | 3061 | 272 | 1310 |
| Events per 100,000 person days |  |  | 15.05 | 8.56 | 9.73 |  |  | 7.59 | 6.62 | 17.97 |  | 9.69 | 8.26 | 26.88 | 20.73 |  | 7.61 | 12.81 | 24.36 | 22.67 |
| Crude HR (95%CI) |  |  | 1.52 (1.36 - 1.70) | 0.90 (0.81 - 1.01) | Ref |  |  | 0.41 (0.39 - 0.44) | 0.37 (0.34 - 0.40) | Ref |  | 0.48 (0.30 - 0.76) | 0.40 (0.37 - 0.42) | 1.31 (1.18 - 1.46) | Ref |  | 0.34 (0.31 - 0.37) | 0.68 (0.63 - 0.73) | 1.07 (0.94 - 1.22) | Ref |
| Age adjusted HR (95% CI) |  |  | 1.19 (1.06 - 1.34) | 0.73 (0.65 - 0.81) | Ref |  |  | 0.37 (0.35 - 0.40) | 0.40 (0.37 - 0.44) | Ref |  | 0.43 (0.27 - 0.68) | 0.39 (0.37 - 0.42) | 1.33 (1.19 - 1.48) | Ref |  | 0.32 (0.29 - 0.34) | 0.70 (0.65 - 0.75) | 1.07 (0.94 - 1.22) | Ref |
| Age and gender adjusted HR (95% CI) |  |  | 1.20 (1.06 - 1.35) | 0.72 (0.65 - 0.81) | Ref |  |  | 0.36 (0.34 - 0.39) | 0.39 (0.36 - 0.43) | Ref |  | 0.41 (0.25 - 0.64) | 0.38 (0.36 - 0.40) | 1.31 (1.17 - 1.46) | Ref |  | 0.31 (0.28 - 0.33) | 0.68 (0.64 - 0.73) | 1.06 (0.93 - 1.20) | Ref |
| Age, sex and ALTCA adjusted HR (95% CI) |  |  | 0.94 (0.83 - 1.05) | 0.71 (0.63 - 0.79) | Ref |  |  | 0.42 (0.39 - 0.44) | 0.45 (0.41 - 0.48) | Ref |  | 0.35 (0.22 - 0.55) | 0.44 (0.41 - 0.47) | 1.19 (1.07 - 1.32) | Ref |  | 0.35 (0.33 - 0.38) | 0.72 (0.67 - 0.77) | 0.93 (0.82 - 1.06) | Ref |
| Age, sex, ALTCA and education adjusted HR (95% CI) |  |  | 0.94 (0.83 - 1.05) | 0.71 (0.63 - 0.79) | Ref |  |  | 0.41 (0.39 - 0.44) | 0.44 (0.40 - 0.48) | Ref |  | 0.34 (0.21 - 0.54) | 0.43 (0.41 - 0.46) | 1.17 (1.05 - 1.31) | Ref |  | 0.35 (0.32 - 0.38) | 0.71 (0.66 - 0.76) | 0.92 (0.80 - 1.05) | Ref |
|  | 2022 | | | | | | | | | | | | | | | | | | | |
| Deaths (n) | 9 | 3620 | 901 | 154 | 807 | 63 | 3282 | 502 | 52 | 540 | 324 | 1937 | 259 | 28 | 365 |  |  |  |  | 0 |
| Events per 100,000 person days |  | 8.63 | 25.41 | 34.4 | 20.14 | 6.24 | 8.74 | 19.96 | 22.9 | 15.69 | 5.7 | 10.14 | 19.18 | 20.03 | 16.69 |  |  |  |  |  |
| Crude HR (95%CI) |  | 0.43 (0.40 - 0.46) | 1.26 (1.14 - 1.38) | 1.70 (1.43 - 2.03) | Ref | 0.41 (0.31 - 0.53) | 0.56 (0.51 - 0.61) | 1.27 (1.12 - 1.43) | 1.46 (1.10 - 1.94) | Ref | 0.34 (0.29 - 0.39) | 0.61 (0.54 - 0.68) | 1.15 (0.98 - 1.35) | 1.21 (0.82 - 1.77) | Ref |  |  |  |  | Ref |
| Age adjusted HR (95% CI) |  | 0.42 (0.38 - 0.45) | 1.24 (1.13 - 1.36) | 1.65 (1.38 - 1.96) | Ref | 0.37 (0.28 - 0.48) | 0.54 (0.50 - 0.59) | 1.24 (1.10 - 1.40) | 1.44 (1.09 - 1.92) | Ref | 0.31 (0.27 - 0.37) | 0.59 (0.53 - 0.66) | 1.12 (0.96 - 1.32) | 1.20 (0.81 - 1.76) | Ref |  |  |  |  | Ref |
| Age and gender adjusted HR (95% CI) |  | 0.40 (0.37 - 0.43) | 1.23 (1.11 - 1.35) | 1.64 (1.38 - 1.95) | Ref | 0.35 (0.27 - 0.46) | 0.53 (0.48 - 0.58) | 1.24 (1.09 - 1.40) | 1.43 (1.08 - 1.91) | Ref | 0.31 (0.26 - 0.36) | 0.58 (0.51 - 0.64) | 1.11 (0.95 - 1.31) | 1.19 (0.81 - 1.75) | Ref |  |  |  |  | Ref |
| Age, sex and ALTCA adjusted HR (95% CI) |  | 0.43 (0.39 - 0.46) | 1.05 (0.95 - 1.16) | 1.37 (1.15 - 1.63) | Ref | 0.37 (0.28 - 0.49) | 0.54 (0.49 - 0.59) | 1.04 (0.92 - 1.17) | 1.24 (0.93 - 1.65) | Ref | 0.35 (0.30 - 0.40) | 0.57 (0.51 - 0.64) | 0.94 (0.80 - 1.10) | 1.04 (0.71 - 1.52) | Ref |  |  |  |  | Ref |
| Age, sex, ALTCA and education adjusted HR (95% CI) |  | 0.42 (0.39 - 0.45) | 1.03 (0.94 - 1.14) | 1.35 (1.14 - 1.61) | Ref | 0.38 (0.29 - 0.50) | 0.53 (0.49 - 0.59) | 1.04 (0.92 - 1.17) | 1.24 (0.93 - 1.64) | Ref | 0.35 (0.30 - 0.41) | 0.56 (0.50 - 0.63) | 0.92 (0.78 - 1.08) | 1.03 (0.70 - 1.51) | Ref |  |  |  |  | Ref |
| Estimates with less than 10 deaths in either the vaccinated or reference group are not shown | | | | | | | | | | | | | | | | | | | | |

| **Table S22:** Hazard ratios (HR) with 95% confidence intervals (95% CI) for all-cause mortality according to number of SARS-CoV-2 vaccine doses for the period from 2021 to 2022 split into 3 month intervals for 85+ year olds only. | | | | | | | | | | | | | | | | | | | | |
| --- | --- | --- | --- | --- | --- | --- | --- | --- | --- | --- | --- | --- | --- | --- | --- | --- | --- | --- | --- | --- |
|  | 2021 | | | | | | | | | | | | | | | | | | | |
|  | All-cause mortality | | | | | | | | | | | | | | | | | | | |
|  | first quarter | | | | | second quarter | | | | | third quarter | | | | | fourth quarter | | | | |
|  | Four vaccine doses | Three vaccine doses | Two vaccine doses | One vaccine dose | Unvaccinated | Four vaccine doses | Three vaccine doses | Two vaccine doses | One vaccine dose | Unvaccinated | Four vaccine doses | Three vaccine doses | Two vaccine doses | One vaccine dose | Unvaccinated | Four vaccine doses | Three vaccine doses | Two vaccine doses | One vaccine dose | Unvaccinated |
| Deaths (n) |  |  | 851 | 793 | 5619 |  | 1 | 3509 | 669 | 2852 |  | 66 | 4501 | 389 | 2072 |  | 2725 | 2826 | 336 | 1910 |
| Events per 100,000 person days |  |  | 33.52 | 28 | 41.87 |  |  | 28.22 | 42.27 | 62.68 |  | 41.2 | 32.03 | 70.46 | 60.23 |  | 29.63 | 48.52 | 69.21 | 71.46 |
| Crude HR (95%CI) |  |  | 0.74 (0.68 - 0.80) | 0.64 (0.59 - 0.69) | Ref |  |  | 0.45 (0.42 - 0.47) | 0.68 (0.62 - 0.73) | Ref |  | 0.70 (0.54 - 0.91) | 0.53 (0.50 - 0.56) | 1.17 (1.05 - 1.31) | Ref |  | 0.40 (0.38 - 0.43) | 0.75 (0.71 - 0.80) | 0.97 (0.86 - 1.09) | Ref |
| Age adjusted HR (95% CI) |  |  | 0.67 (0.62 - 0.73) | 0.64 (0.59 - 0.69) | Ref |  |  | 0.49 (0.46 - 0.51) | 0.73 (0.67 - 0.80) | Ref |  | 0.68 (0.52 - 0.88) | 0.60 (0.57 - 0.63) | 1.21 (1.08 - 1.34) | Ref |  | 0.44 (0.42 - 0.47) | 0.84 (0.79 - 0.89) | 0.98 (0.87 - 1.10) | Ref |
| Age and gender adjusted HR (95% CI) |  |  | 0.68 (0.63 - 0.73) | 0.63 (0.59 - 0.69) | Ref |  |  | 0.48 (0.45 - 0.50) | 0.72 (0.66 - 0.78) | Ref |  | 0.67 (0.52 - 0.87) | 0.59 (0.56 - 0.62) | 1.19 (1.07 - 1.33) | Ref |  | 0.43 (0.41 - 0.46) | 0.82 (0.77 - 0.87) | 0.97 (0.86 - 1.09) | Ref |
| Age, sex and ALTCA adjusted HR (95% CI) |  |  | 0.64 (0.59 - 0.70) | 0.62 (0.58 - 0.68) | Ref |  |  | 0.48 (0.46 - 0.51) | 0.71 (0.66 - 0.78) | Ref |  | 0.64 (0.49 - 0.83) | 0.59 (0.56 - 0.62) | 1.15 (1.03 - 1.28) | Ref |  | 0.43 (0.41 - 0.46) | 0.81 (0.76 - 0.86) | 0.94 (0.83 - 1.05) | Ref |
| Age, sex, ALTCA and education adjusted HR (95% CI) |  |  | 0.64 (0.59 - 0.69) | 0.62 (0.58 - 0.67) | Ref |  |  | 0.48 (0.46 - 0.50) | 0.71 (0.65 - 0.77) | Ref |  | 0.63 (0.49 - 0.82) | 0.59 (0.56 - 0.62) | 1.14 (1.02 - 1.27) | Ref |  | 0.43 (0.41 - 0.46) | 0.80 (0.75 - 0.85) | 0.93 (0.83 - 1.04) | Ref |
|  | 2022 | | | | | | | | | | | | | | | | | | | |
| Deaths (n) | 5 | 4498 | 1020 | 205 | 1237 | 88 | 3537 | 696 | 87 | 800 | 443 | 2107 | 341 | 39 | 493 |  |  |  |  | 0 |
| Events per 100,000 person days |  | 33.85 | 66.11 | 81.26 | 73.21 | 19.78 | 30.73 | 59.83 | 83.46 | 58.79 | 22.64 | 35.92 | 57.15 | 64.12 | 58.90 |  |  |  |  |  |
| Crude HR (95%CI) |  | 0.46 (0.43 - 0.49) | 0.90 (0.83 - 0.98) | 1.11 (0.96 - 1.29) | Ref | 0.35 (0.28 - 0.44) | 0.52 (0.48 - 0.56) | 1.01 (0.92 - 1.12) | 1.41 (1.13 - 1.76) | Ref | 0.39 (0.34 - 0.45) | 0.62 (0.56 - 0.69) | 0.98 (0.86 - 1.13) | 1.05 (0.76 - 1.46) | Ref |  |  |  |  | Ref |
| Age adjusted HR (95% CI) |  | 0.52 (0.49 - 0.55) | 0.92 (0.84 - 0.99) | 1.08 (0.93 - 1.26) | Ref | 0.37 (0.30 - 0.47) | 0.59 (0.54 - 0.63) | 1.01 (0.91 - 1.12) | 1.39 (1.12 - 1.74) | Ref | 0.42 (0.37 - 0.48) | 0.69 (0.62 - 0.76) | 0.98 (0.86 - 1.13) | 1.05 (0.76 - 1.46) | Ref |  |  |  |  | Ref |
| Age and gender adjusted HR (95% CI) |  | 0.51 (0.47 - 0.54) | 0.90 (0.83 - 0.98) | 1.08 (0.93 - 1.25) | Ref | 0.36 (0.28 - 0.45) | 0.57 (0.53 - 0.62) | 1.00 (0.91 - 1.11) | 1.39 (1.11 - 1.73) | Ref | 0.41 (0.36 - 0.47) | 0.67 (0.61 - 0.74) | 0.97 (0.85 - 1.12) | 1.05 (0.76 - 1.46) | Ref |  |  |  |  | Ref |
| Age, sex and ALTCA adjusted HR (95% CI) |  | 0.49 (0.46 - 0.52) | 0.84 (0.77 - 0.91) | 1.00 (0.87 - 1.17) | Ref | 0.36 (0.29 - 0.46) | 0.54 (0.50 - 0.58) | 0.92 (0.83 - 1.02) | 1.30 (1.04 - 1.62) | Ref | 0.41 (0.36 - 0.47) | 0.63 (0.57 - 0.70) | 0.89 (0.77 - 1.02) | 1.01 (0.73 - 1.40) | Ref |  |  |  |  | Ref |
| Age, sex, ALTCA and education adjusted HR (95% CI) |  | 0.48 (0.45 - 0.52) | 0.83 (0.77 - 0.91) | 1.00 (0.86 - 1.16) | Ref | 0.37 (0.29 - 0.47) | 0.54 (0.50 - 0.58) | 0.92 (0.83 - 1.01) | 1.28 (1.02 - 1.60) | Ref | 0.41 (0.36 - 0.47) | 0.63 (0.57 - 0.70) | 0.89 (0.77 - 1.02) | 1.01 (0.73 - 1.40) | Ref |  |  |  |  | Ref |
| Estimates with less than 10 deaths in either the vaccinated or reference group are not shown | | | | | | | | | | | | | | | | | | | | |

| **Table S23:** Hazard ratios (HR) with 95% confidence intervals (95% CI) for all-cause mortality according to number of SARS-CoV-2 vaccine doses for the period from 2021 to 2022 split into 3 month intervals for individuals with compulsory schooling (9 years) or lower only. | | | | | | | | | | | | | | | | | | | | |
| --- | --- | --- | --- | --- | --- | --- | --- | --- | --- | --- | --- | --- | --- | --- | --- | --- | --- | --- | --- | --- |
|  | 2021 | | | | | | | | | | | | | | | | | | | |
|  | All-cause mortality | | | | | | | | | | | | | | | | | | | |
|  | first quarter | | | | | second quarter | | | | | third quarter | | | | | fourth quarter | | | | |
|  | Four vaccine doses | Three vaccine doses | Two vaccine doses | One vaccine dose | Unvaccinated | Four vaccine doses | Three vaccine doses | Two vaccine doses | One vaccine dose | Unvaccinated | Four vaccine doses | Three vaccine doses | Two vaccine doses | One vaccine dose | Unvaccinated | Four vaccine doses | Three vaccine doses | Two vaccine doses | One vaccine dose | Unvaccinated |
| Deaths (n) |  |  | 673 | 637 | 6227 |  | 1 | 2875 | 852 | 3528 |  | 41 | 4317 | 496 | 2597 |  | 2275 | 3234 | 407 | 2321 |
| Events per 100,000 person days |  |  | 21.39 | 13.27 | 4.78 |  |  | 10.72 | 3.74 | 4.10 |  | 22.53 | 6.09 | 4.16 | 4.97 |  | 9.76 | 5.29 | 4.3 | 6.41 |
| Crude HR (95%CI) |  |  | 5.07 (4.66 - 5.51) | 3.16 (2.91 - 3.44) | Ref |  |  | 2.72 (2.58 - 2.86) | 0.92 (0.85 - 0.99) | Ref |  | 4.16 (3.03 - 5.73) | 1.23 (1.17 - 1.29) | 0.85 (0.77 - 0.94) | Ref |  | 1.74 (1.62 - 1.85) | 0.83 (0.78 - 0.87) | 0.66 (0.60 - 0.74) | Ref |
| Age adjusted HR (95% CI) |  |  | 0.97 (0.89 - 1.06) | 0.79 (0.72 - 0.86) | Ref |  |  | 0.50 (0.47 - 0.52) | 0.49 (0.46 - 0.53) | Ref |  | 0.63 (0.45 - 0.86) | 0.49 (0.47 - 0.51) | 1.10 (1.00 - 1.21) | Ref |  | 0.42 (0.39 - 0.45) | 0.65 (0.61 - 0.68) | 0.95 (0.86 - 1.06) | Ref |
| Age and sex adjusted HR (95% CI) |  |  | 0.99 (0.90 - 1.08) | 0.79 (0.72 - 0.86) | Ref |  |  | 0.49 (0.47 - 0.52) | 0.50 (0.46 - 0.54) | Ref |  | 0.63 (0.46 - 0.87) | 0.49 (0.47 - 0.51) | 1.10 (1.00 - 1.21) | Ref |  | 0.42 (0.39 - 0.44) | 0.64 (0.61 - 0.68) | 0.94 (0.85 - 1.05) | Ref |
| Age, sex and ALTCA adjusted HR (95% CI) |  |  | 0.80 (0.73 - 0.87) | 0.71 (0.65 - 0.77) | Ref |  |  | 0.48 (0.46 - 0.51) | 0.56 (0.52 - 0.60) | Ref |  | 0.52 (0.38 - 0.72) | 0.51 (0.49 - 0.54) | 1.07 (0.98 - 1.18) | Ref |  | 0.42 (0.39 - 0.45) | 0.68 (0.64 - 0.72) | 0.91 (0.81 - 1.01) | Ref |
|  | 2022 | | | | | | | | | | | | | | | | | | | |
| Deaths (n) | 5 | 4145 | 1319 | 269 | 1461 | 68 | 3611 | 851 | 111 | 996 | 327 | 2224 | 422 | 52 | 610 |  |  |  |  | 0 |
| Events per 100,000 person days |  | 6.73 | 5.13 | 8.88 | 6.74 | 14.1 | 6.2 | 4.96 | 6.21 | 5.78 | 10.61 | 6.4 | 4.21 | 4.56 | 5.53 |  |  |  |  |  |
| Crude HR (95%CI) |  | 1.00 (0.94 - 1.07) | 0.76 (0.71 - 0.82) | 1.33 (1.16 - 1.51) | Ref | 2.53 (1.97 - 3.25) | 1.07 (1.00 - 1.15) | 0.86 (0.78 - 0.94) | 1.07 (0.88 - 1.31) | Ref | 1.87 (1.63 - 2.15) | 1.16 (1.06 - 1.26) | 0.77 (0.68 - 0.88) | 0.81 (0.61 - 1.08) | Ref |  |  |  |  | Ref |
| Age adjusted HR (95% CI) |  | 0.44 (0.42 - 0.47) | 0.97 (0.90 - 1.04) | 1.34 (1.17 - 1.53) | Ref | 0.53 (0.41 - 0.68) | 0.52 (0.48 - 0.56) | 1.02 (0.93 - 1.12) | 1.40 (1.15 - 1.70) | Ref | 0.44 (0.38 - 0.50) | 0.60 (0.55 - 0.66) | 0.99 (0.87 - 1.12) | 1.07 (0.81 - 1.42) | Ref |  |  |  |  | Ref |
| Age and sex adjusted HR (95% CI) |  | 0.44 (0.42 - 0.47) | 0.96 (0.89 - 1.03) | 1.33 (1.16 - 1.51) | Ref | 0.52 (0.40 - 0.67) | 0.52 (0.48 - 0.56) | 1.02 (0.93 - 1.11) | 1.39 (1.14 - 1.69) | Ref | 0.43 (0.38 - 0.50) | 0.60 (0.55 - 0.66) | 0.98 (0.87 - 1.11) | 1.07 (0.80 - 1.42) | Ref |  |  |  |  | Ref |
| Age, sex and ALTCA adjusted HR (95% CI) |  | 0.44 (0.42 - 0.47) | 0.88 (0.82 - 0.95) | 1.21 (1.06 - 1.38) | Ref | 0.49 (0.38 - 0.63) | 0.51 (0.48 - 0.55) | 0.92 (0.84 - 1.00) | 1.31 (1.08 - 1.59) | Ref | 0.43 (0.38 - 0.50) | 0.59 (0.54 - 0.65) | 0.89 (0.78 - 1.00) | 1.05 (0.79 - 1.39) | Ref |  |  |  |  | Ref |
| Estimates with less than 10 deaths in either the vaccinated or reference group are not shown | | | | | | | | | | | | | | | | | | | | |

| **Table S24:** Hazard ratios (HR) with 95% confidence intervals (95% CI) for all-cause mortality according to number of SARS-CoV-2 vaccine doses for the period from 2021 to 2022 split into 3 month intervals for individuals with non-secondary education only. | | | | | | | | | | | | | | | | | | | | |
| --- | --- | --- | --- | --- | --- | --- | --- | --- | --- | --- | --- | --- | --- | --- | --- | --- | --- | --- | --- | --- |
|  | 2021 | | | | | | | | | | | | | | | | | | | |
|  | All-cause mortality | | | | | | | | | | | | | | | | | | | |
|  | first quarter | | | | | second quarter | | | | | third quarter | | | | | fourth quarter | | | | |
|  | Four vaccine doses | Three vaccine doses | Two vaccine doses | One vaccine dose | Unvaccinated | Four vaccine doses | Three vaccine doses | Two vaccine doses | One vaccine dose | Unvaccinated | Four vaccine doses | Three vaccine doses | Two vaccine doses | One vaccine dose | Unvaccinated | Four vaccine doses | Three vaccine doses | Two vaccine doses | One vaccine dose | Unvaccinated |
| Deaths (n) |  |  | 565 | 565 | 7250 |  |  | 3149 | 1301 | 4136 |  | 48 | 5565 | 693 | 2553 |  | 2369 | 4682 | 458 | 2112 |
| Events per 100,000 person days |  |  | 9.67 | 5.99 | 2.55 |  |  | 5.75 | 1.93 | 2.37 |  | 14.75 | 3.08 | 2.81 | 2.79 |  | 4.14 | 3.15 | 3.05 | 3.25 |
| Crude HR (95%CI) |  |  | 4.10 (3.75 - 4.48) | 2.58 (2.36 - 2.82) | Ref |  |  | 2.59 (2.46 - 2.72) | 0.82 (0.77 - 0.87) | Ref |  | 5.44 (4.05 - 7.31) | 1.11 (1.05 - 1.16) | 1.03 (0.95 - 1.13) | Ref |  | 1.49 (1.39 - 1.59) | 0.98 (0.93 - 1.03) | 0.93 (0.84 - 1.03) | Ref |
| Age adjusted HR (95% CI) |  |  | 1.03 (0.94 - 1.14) | 0.75 (0.69 - 0.82) | Ref |  |  | 0.44 (0.42 - 0.47) | 0.40 (0.38 - 0.43) | Ref |  | 0.74 (0.55 - 1.00) | 0.44 (0.42 - 0.46) | 1.14 (1.04 - 1.24) | Ref |  | 0.33 (0.31 - 0.35) | 0.69 (0.66 - 0.73) | 1.02 (0.92 - 1.13) | Ref |
| Age and sex adjusted HR (95% CI) |  |  | 1.06 (0.97 - 1.16) | 0.75 (0.68 - 0.82) | Ref |  |  | 0.43 (0.41 - 0.45) | 0.40 (0.37 - 0.42) | Ref |  | 0.72 (0.54 - 0.98) | 0.43 (0.41 - 0.45) | 1.11 (1.02 - 1.20) | Ref |  | 0.32 (0.30 - 0.34) | 0.67 (0.64 - 0.71) | 0.99 (0.90 - 1.10) | Ref |
| Age, sex and ALTCA adjusted HR (95% CI) |  |  | 0.78 (0.71 - 0.85) | 0.65 (0.60 - 0.71) | Ref |  |  | 0.46 (0.44 - 0.48) | 0.49 (0.46 - 0.52) | Ref |  | 0.57 (0.43 - 0.77) | 0.50 (0.48 - 0.53) | 1.07 (0.98 - 1.16) | Ref |  | 0.36 (0.33 - 0.38) | 0.75 (0.71 - 0.79) | 0.91 (0.82 - 1.01) | Ref |
|  | 2022 | | | | | | | | | | | | | | | | | | | |
| Deaths (n) | 14 | 5425 | 1500 | 237 | 1332 | 89 | 4838 | 877 | 102 | 843 | 425 | 3008 | 476 | 57 | 586 |  |  |  |  | 0 |
| Events per 100,000 person days | 11.95 | 3.43 | 3.57 | 7.33 | 3.65 | 8.33 | 3.46 | 3.7 | 6.27 | 3.30 | 5.88 | 3.69 | 3.62 | 5.6 | 3.64 |  |  |  |  |  |
| Crude HR (95%CI) | 3.12 (1.84 - 5.29) | 0.93 (0.88 - 0.99) | 0.99 (0.92 - 1.06) | 2.05 (1.78 - 2.35) | Ref | 2.53 (2.02 - 3.17) | 1.05 (0.98 - 1.13) | 1.12 (1.02 - 1.23) | 1.90 (1.55 - 2.34) | Ref | 1.63 (1.43 - 1.85) | 1.02 (0.93 - 1.12) | 1.00 (0.89 - 1.13) | 1.49 (1.14 - 1.96) | Ref |  |  |  |  | Ref |
| Age adjusted HR (95% CI) | 0.90 (0.53 - 1.52) | 0.43 (0.41 - 0.46) | 1.14 (1.06 - 1.22) | 1.59 (1.38 - 1.83) | Ref | 0.45 (0.36 - 0.57) | 0.56 (0.52 - 0.61) | 1.26 (1.14 - 1.38) | 1.85 (1.51 - 2.27) | Ref | 0.35 (0.31 - 0.40) | 0.61 (0.56 - 0.67) | 1.16 (1.03 - 1.31) | 1.52 (1.16 - 2.00) | Ref |  |  |  |  | Ref |
| Age and sex adjusted HR (95% CI) | 0.84 (0.50 - 1.42) | 0.41 (0.39 - 0.44) | 1.10 (1.02 - 1.19) | 1.56 (1.35 - 1.79) | Ref | 0.43 (0.34 - 0.54) | 0.54 (0.50 - 0.58) | 1.22 (1.11 - 1.34) | 1.81 (1.47 - 2.22) | Ref | 0.33 (0.29 - 0.38) | 0.59 (0.54 - 0.65) | 1.13 (1.00 - 1.27) | 1.48 (1.13 - 1.95) | Ref |  |  |  |  | Ref |
| Age, sex and ALTCA adjusted HR (95% CI) | 0.70 (0.42 - 1.19) | 0.45 (0.43 - 0.48) | 0.99 (0.92 - 1.07) | 1.31 (1.14 - 1.51) | Ref | 0.42 (0.34 - 0.53) | 0.58 (0.54 - 0.63) | 1.07 (0.98 - 1.18) | 1.57 (1.28 - 1.93) | Ref | 0.36 (0.32 - 0.42) | 0.62 (0.57 - 0.68) | 0.99 (0.88 - 1.12) | 1.27 (0.97 - 1.67) | Ref |  |  |  |  | Ref |

| **Table S25:** Hazard ratios (HR) with 95% confidence intervals (95% CI) for all-cause mortality according to number of SARS-CoV-2 vaccine doses for the period from 2021 to 2022 split into 3 month intervals for individuals with secondary education only. | | | | | | | | | | | | | | | | | | | | |
| --- | --- | --- | --- | --- | --- | --- | --- | --- | --- | --- | --- | --- | --- | --- | --- | --- | --- | --- | --- | --- |
|  | 2021 | | | | | | | | | | | | | | | | | | | |
|  | All-cause mortality | | | | | | | | | | | | | | | | | | | |
|  | first quarter | | | | | second quarter | | | | | third quarter | | | | | fourth quarter | | | | |
|  | Four vaccine doses | Three vaccine doses | Two vaccine doses | One vaccine dose | Unvaccinated | Four vaccine doses | Three vaccine doses | Two vaccine doses | One vaccine dose | Unvaccinated | Four vaccine doses | Three vaccine doses | Two vaccine doses | One vaccine dose | Unvaccinated | Four vaccine doses | Three vaccine doses | Two vaccine doses | One vaccine dose | Unvaccinated |
| Deaths (n) |  |  | 76 | 100 | 1135 |  |  | 449 | 196 | 669 |  | 5 | 900 | 114 | 383 | 1 | 393 | 642 | 58 | 304 |
| Events per 100,000 person days |  |  | 5.11 | 3.52 | 1.20 |  |  | 3.14 | 0.91 | 1.08 |  |  | 1.53 | 1.08 | 1.34 |  | 2.31 | 1.23 | 1.15 | 1.51 |
| Crude HR (95%CI) |  |  | 4.62 (3.64 - 5.87) | 3.28 (2.65 - 4.06) | Ref |  |  | 2.90 (2.54 - 3.31) | 0.80 (0.68 - 0.94) | Ref |  |  | 1.16 (1.03 - 1.31) | 0.89 (0.72 - 1.10) | Ref |  | 2.04 (1.69 - 2.46) | 0.82 (0.71 - 0.94) | 0.76 (0.57 - 1.01) | Ref |
| Age adjusted HR (95% CI) |  |  | 0.84 (0.65 - 1.08) | 0.80 (0.65 - 1.00) | Ref |  |  | 0.34 (0.30 - 0.39) | 0.39 (0.33 - 0.46) | Ref |  |  | 0.49 (0.43 - 0.55) | 1.45 (1.17 - 1.79) | Ref |  | 0.39 (0.33 - 0.45) | 0.76 (0.66 - 0.88) | 1.11 (0.84 - 1.47) | Ref |
| Age and sex adjusted HR (95% CI) |  |  | 0.86 (0.67 - 1.10) | 0.80 (0.65 - 1.00) | Ref |  |  | 0.33 (0.29 - 0.37) | 0.37 (0.32 - 0.44) | Ref |  |  | 0.46 (0.41 - 0.52) | 1.41 (1.14 - 1.75) | Ref |  | 0.37 (0.31 - 0.44) | 0.74 (0.64 - 0.85) | 1.10 (0.83 - 1.45) | Ref |
| Age, sex and ALTCA adjusted HR (95% CI) |  |  | 0.61 (0.48 - 0.79) | 0.68 (0.55 - 0.84) | Ref |  |  | 0.35 (0.30 - 0.39) | 0.46 (0.39 - 0.54) | Ref |  |  | 0.52 (0.46 - 0.59) | 1.20 (0.97 - 1.49) | Ref |  | 0.39 (0.33 - 0.46) | 0.75 (0.66 - 0.87) | 0.91 (0.69 - 1.21) | Ref |
|  | 2022 | | | | | | | | | | | | | | | | | | | |
| Deaths (n) | 1 | 797 | 205 | 33 | 194 | 19 | 805 | 119 | 13 | 119 | 89 | 437 | 56 | 4 | 97 |  |  |  |  | 0 |
| Events per 100,000 person days |  | 1.58 | 1.45 | 2.9 | 1.63 | 5.51 | 1.91 | 1.67 | 2 | 1.35 | 4.49 | 1.86 | 1.44 |  | 1.75 |  |  |  |  |  |
| Crude HR (95%CI) |  | 0.97 (0.83 - 1.14) | 0.89 (0.73 - 1.08) | 1.80 (1.24 - 2.60) | Ref | 4.42 (2.68 - 7.28) | 1.41 (1.17 - 1.72) | 1.24 (0.96 - 1.60) | 1.47 (0.83 - 2.61) | Ref | 2.48 (1.85 - 3.32) | 1.06 (0.85 - 1.32) | 0.83 (0.60 - 1.16) |  | Ref |  |  |  |  | Ref |
| Age adjusted HR (95% CI) |  | 0.50 (0.42 - 0.58) | 1.40 (1.15 - 1.71) | 1.75 (1.21 - 2.54) | Ref | 0.44 (0.26 - 0.73) | 0.81 (0.66 - 0.98) | 1.64 (1.27 - 2.12) | 1.84 (1.04 - 3.26) | Ref | 0.37 (0.27 - 0.50) | 0.71 (0.57 - 0.89) | 1.14 (0.82 - 1.59) |  | Ref |  |  |  |  | Ref |
| Age and sex adjusted HR (95% CI) |  | 0.47 (0.40 - 0.56) | 1.38 (1.14 - 1.69) | 1.75 (1.20 - 2.53) | Ref | 0.38 (0.23 - 0.64) | 0.76 (0.63 - 0.93) | 1.61 (1.25 - 2.08) | 1.84 (1.04 - 3.26) | Ref | 0.35 (0.26 - 0.48) | 0.68 (0.54 - 0.84) | 1.13 (0.81 - 1.57) |  | Ref |  |  |  |  | Ref |
| Age, sex and ALTCA adjusted HR (95% CI) |  | 0.48 (0.41 - 0.56) | 1.11 (0.91 - 1.35) | 1.37 (0.95 - 2.00) | Ref | 0.37 (0.22 - 0.61) | 0.75 (0.62 - 0.92) | 1.31 (1.01 - 1.69) | 1.48 (0.83 - 2.62) | Ref | 0.37 (0.27 - 0.51) | 0.65 (0.52 - 0.81) | 0.88 (0.63 - 1.23) |  | Ref |  |  |  |  | Ref |
| Estimates with less than 10 deaths in either the vaccinated or reference group are not shown | | | | | | | | | | | | | | | | | | | | |

| **Table S26:** Hazard ratios (HR) with 95% confidence intervals (95% CI) for all-cause mortality according to number of SARS-CoV-2 vaccine doses for the period from 2021 to 2022 split into 3 month intervals for individuals withnon-tertiary education only. | | | | | | | | | | | | | | | | | | | | |
| --- | --- | --- | --- | --- | --- | --- | --- | --- | --- | --- | --- | --- | --- | --- | --- | --- | --- | --- | --- | --- |
|  | 2021 | | | | | | | | | | | | | | | | | | | |
|  | All-cause mortality | | | | | | | | | | | | | | | | | | | |
|  | first quarter | | | | | second quarter | | | | | third quarter | | | | | fourth quarter | | | | |
|  | Four vaccine doses | Three vaccine doses | Two vaccine doses | One vaccine dose | Unvaccinated | Four vaccine doses | Three vaccine doses | Two vaccine doses | One vaccine dose | Unvaccinated | Four vaccine doses | Three vaccine doses | Two vaccine doses | One vaccine dose | Unvaccinated | Four vaccine doses | Three vaccine doses | Two vaccine doses | One vaccine dose | Unvaccinated |
| Deaths (n) |  |  | 9 | 7 | 204 |  |  | 79 | 40 | 85 |  |  | 138 | 16 | 48 |  | 69 | 138 | 5 | 50 |
| Events per 100,000 person days |  |  |  |  | 1.32 |  |  | 1.95 | 0.76 | 1.12 |  |  | 1.16 | 1.77 | 1.17 |  | 1.57 | 1.66 |  | 1.56 |
| Crude HR (95%CI) |  |  |  |  | Ref |  |  | 1.84 (1.31 - 2.58) | 0.65 (0.44 - 0.95) | Ref |  |  | 0.99 (0.72 - 1.38) | 1.49 (0.84 - 2.67) | Ref |  | 1.21 (0.77 - 1.89) | 1.17 (0.84 - 1.63) |  | Ref |
| Age adjusted HR (95% CI) |  |  |  |  | Ref |  |  | 0.46 (0.33 - 0.65) | 0.45 (0.30 - 0.66) | Ref |  |  | 0.51 (0.37 - 0.71) | 1.67 (0.94 - 2.97) | Ref |  | 0.37 (0.25 - 0.55) | 0.96 (0.68 - 1.34) |  | Ref |
| Age and sex adjusted HR (95% CI) |  |  |  |  | Ref |  |  | 0.46 (0.33 - 0.64) | 0.45 (0.30 - 0.66) | Ref |  |  | 0.50 (0.36 - 0.70) | 1.67 (0.94 - 2.97) | Ref |  | 0.37 (0.25 - 0.55) | 0.95 (0.68 - 1.34) |  | Ref |
| Age, sex and ALTCA adjusted HR (95% CI) |  |  |  |  | Ref |  |  | 0.47 (0.34 - 0.65) | 0.58 (0.39 - 0.85) | Ref |  |  | 0.58 (0.41 - 0.80) | 1.36 (0.77 - 2.40) | Ref |  | 0.39 (0.26 - 0.57) | 0.99 (0.70 - 1.38) |  | Ref |
|  | 2022 | | | | | | | | | | | | | | | | | | | |
| Deaths (n) |  | 138 | 44 | 8 | 38 | 3 | 131 | 20 | 1 | 27 | 15 | 64 | 8 | 3 | 11 |  |  |  |  | 0 |
| Events per 100,000 person days |  | 1.36 | 3.21 |  | 1.78 |  | 1.63 | 2.88 |  | 1.64 | 2.59 | 1.51 |  |  | 1.06 |  |  |  |  |  |
| Crude HR (95%CI) |  | 0.76 (0.53 - 1.08) | 1.91 (1.24 - 2.95) |  | Ref |  | 0.99 (0.66 - 1.50) | 1.74 (0.98 - 3.11) |  | Ref | 1.79 (0.79 - 4.08) | 1.42 (0.75 - 2.69) |  |  | Ref |  |  |  |  | Ref |
| Age adjusted HR (95% CI) |  | 0.41 (0.28 - 0.58) | 1.85 (1.20 - 2.86) |  | Ref |  | 0.54 (0.36 - 0.82) | 1.55 (0.87 - 2.77) |  | Ref | 0.46 (0.19 - 1.10) | 0.86 (0.45 - 1.63) |  |  | Ref |  |  |  |  | Ref |
| Age and sex adjusted HR (95% CI) |  | 0.40 (0.28 - 0.58) | 1.86 (1.20 - 2.88) |  | Ref |  | 0.55 (0.36 - 0.83) | 1.55 (0.87 - 2.77) |  | Ref | 0.46 (0.19 - 1.09) | 0.86 (0.45 - 1.64) |  |  | Ref |  |  |  |  | Ref |
| Age, sex and ALTCA adjusted HR (95% CI) |  | 0.40 (0.28 - 0.58) | 1.37 (0.88 - 2.13) |  | Ref |  | 0.54 (0.36 - 0.82) | 1.17 (0.65 - 2.11) |  | Ref | 0.48 (0.20 - 1.17) | 0.80 (0.42 - 1.53) |  |  | Ref |  |  |  |  | Ref |
| Estimates with less than 10 deaths in either the vaccinated or reference group are not shown | | | | | | | | | | | | | | | | | | | | |

| **Table S27:** Hazard ratios (HR) with 95% confidence intervals (95% CI) for all-cause mortality according to number of SARS-CoV-2 vaccine doses for the period from 2021 to 2022 split into 3 month intervals for individuals with tertiary education only. | | | | | | | | | | | | | | | | | | | | |
| --- | --- | --- | --- | --- | --- | --- | --- | --- | --- | --- | --- | --- | --- | --- | --- | --- | --- | --- | --- | --- |
|  | 2021 | | | | | | | | | | | | | | | | | | | |
|  | All-cause mortality | | | | | | | | | | | | | | | | | | | |
|  | first quarter | | | | | second quarter | | | | | third quarter | | | | | fourth quarter | | | | |
|  | Four vaccine doses | Three vaccine doses | Two vaccine doses | One vaccine dose | Unvaccinated | Four vaccine doses | Three vaccine doses | Two vaccine doses | One vaccine dose | Unvaccinated | Four vaccine doses | Three vaccine doses | Two vaccine doses | One vaccine dose | Unvaccinated | Four vaccine doses | Three vaccine doses | Two vaccine doses | One vaccine dose | Unvaccinated |
| Deaths (n) |  |  | 51 | 62 | 747 |  |  | 354 | 154 | 381 |  | 6 | 655 | 66 | 225 |  | 262 | 466 | 42 | 210 |
| Events per 100,000 person days |  |  | 1.74 | 1.34 | 0.84 |  |  | 1.85 | 0.6 | 0.74 |  |  | 0.98 | 0.74 | 1.09 |  | 1.15 | 0.89 | 1.25 | 1.37 |
| Crude HR (95%CI) |  |  | 2.10 (1.57 - 2.80) | 1.67 (1.28 - 2.19) | Ref |  |  | 2.86 (2.44 - 3.35) | 0.82 (0.68 - 1.00) | Ref |  |  | 0.90 (0.77 - 1.05) | 0.74 (0.55 - 0.98) | Ref |  | 0.98 (0.79 - 1.21) | 0.67 (0.57 - 0.79) | 0.91 (0.66 - 1.27) | Ref |
| Age adjusted HR (95% CI) |  |  | 0.68 (0.50 - 0.92) | 0.60 (0.46 - 0.79) | Ref |  |  | 0.43 (0.37 - 0.51) | 0.48 (0.40 - 0.58) | Ref |  |  | 0.53 (0.46 - 0.62) | 1.42 (1.07 - 1.88) | Ref |  | 0.32 (0.27 - 0.39) | 0.83 (0.69 - 0.98) | 1.28 (0.92 - 1.78) | Ref |
| Age and sex adjusted HR (95% CI) |  |  | 0.67 (0.50 - 0.91) | 0.59 (0.45 - 0.78) | Ref |  |  | 0.42 (0.36 - 0.49) | 0.47 (0.39 - 0.57) | Ref |  |  | 0.51 (0.44 - 0.59) | 1.40 (1.06 - 1.85) | Ref |  | 0.31 (0.25 - 0.38) | 0.80 (0.67 - 0.95) | 1.27 (0.91 - 1.77) | Ref |
| Age, sex and ALTCA adjusted HR (95% CI) |  |  | 0.54 (0.40 - 0.72) | 0.54 (0.41 - 0.71) | Ref |  |  | 0.44 (0.38 - 0.52) | 0.58 (0.48 - 0.70) | Ref |  |  | 0.55 (0.47 - 0.64) | 1.13 (0.85 - 1.49) | Ref |  | 0.32 (0.26 - 0.39) | 0.77 (0.65 - 0.91) | 0.98 (0.70 - 1.37) | Ref |
|  | 2022 | | | | | | | | | | | | | | | | | | | |
| Deaths (n) | 5 | 556 | 131 | 20 | 139 | 20 | 538 | 85 | 8 | 99 | 74 | 328 | 38 | 4 | 66 |  |  |  |  | 0 |
| Events per 100,000 person days |  | 0.97 | 1.32 | 2.04 | 1.39 | 3.57 | 1.2 | 1.77 |  | 1.27 | 2.88 | 1.4 | 1.49 |  | 1.35 |  |  |  |  |  |
| Crude HR (95%CI) |  | 0.69 (0.57 - 0.83) | 0.97 (0.76 - 1.23) | 1.46 (0.91 - 2.33) | Ref | 2.94 (1.80 - 4.80) | 0.95 (0.77 - 1.18) | 1.40 (1.05 - 1.88) |  | Ref | 2.22 (1.58 - 3.10) | 1.03 (0.79 - 1.35) | 1.09 (0.73 - 1.62) |  | Ref |  |  |  |  | Ref |
| Age adjusted HR (95% CI) |  | 0.47 (0.39 - 0.57) | 1.46 (1.15 - 1.86) | 1.50 (0.94 - 2.40) | Ref | 0.43 (0.26 - 0.71) | 0.66 (0.53 - 0.82) | 1.69 (1.26 - 2.25) |  | Ref | 0.41 (0.29 - 0.58) | 0.85 (0.65 - 1.11) | 1.27 (0.85 - 1.90) |  | Ref |  |  |  |  | Ref |
| Age and sex adjusted HR (95% CI) |  | 0.45 (0.37 - 0.54) | 1.46 (1.15 - 1.85) | 1.50 (0.94 - 2.40) | Ref | 0.41 (0.24 - 0.68) | 0.63 (0.51 - 0.79) | 1.68 (1.26 - 2.24) |  | Ref | 0.40 (0.28 - 0.57) | 0.82 (0.63 - 1.07) | 1.25 (0.83 - 1.87) |  | Ref |  |  |  |  | Ref |
| Age, sex and ALTCA adjusted HR (95% CI) |  | 0.43 (0.35 - 0.52) | 1.07 (0.84 - 1.36) | 1.10 (0.68 - 1.77) | Ref | 0.38 (0.23 - 0.64) | 0.58 (0.47 - 0.73) | 1.25 (0.93 - 1.67) |  | Ref | 0.41 (0.28 - 0.58) | 0.73 (0.56 - 0.95) | 0.90 (0.59 - 1.36) |  | Ref |  |  |  |  | Ref |
| Estimates with less than 10 deaths in either the vaccinated or reference group are not shown | | | | | | | | | | | | | | | | | | | | |

| **Table S28:** Hazard ratios (HR) with 95% confidence intervals (95% CI) for all-cause mortality according to number of SARS-CoV-2 vaccine doses for the period from 2021 to 2022 split into 3 month intervals for ALTCA recipients only. | | | | | | | | | | | | | | | | | | | | |
| --- | --- | --- | --- | --- | --- | --- | --- | --- | --- | --- | --- | --- | --- | --- | --- | --- | --- | --- | --- | --- |
|  | 2021 | | | | | | | | | | | | | | | | | | | |
|  | All-cause mortality | | | | | | | | | | | | | | | | | | | |
|  | first quarter | | | | | second quarter | | | | | third quarter | | | | | fourth quarter | | | | |
|  | Four vaccine doses | Three vaccine doses | Two vaccine doses | One vaccine dose | Unvaccinated | Four vaccine doses | Three vaccine doses | Two vaccine doses | One vaccine dose | Unvaccinated | Four vaccine doses | Three vaccine doses | Two vaccine doses | One vaccine dose | Unvaccinated | Four vaccine doses | Three vaccine doses | Two vaccine doses | One vaccine dose | Unvaccinated |
| Deaths (n) |  |  | 1288 | 1172 | 10134 |  | 1 | 5459 | 1543 | 5623 |  | 92 | 8115 | 917 | 3864 | 1 | 4234 | 6015 | 672 | 3302 |
| Events per 100,000 person days |  |  | 34.5 | 27.02 | 29.41 |  |  | 26.37 | 22.15 | 41.71 |  | 34.12 | 27.26 | 48.65 | 46.20 |  | 29.16 | 37.12 | 48.82 | 53.58 |
| Crude HR (95%CI) |  |  | 1.18 (1.11 - 1.25) | 0.93 (0.88 - 0.99) | Ref |  |  | 0.62 (0.60 - 0.64) | 0.53 (0.50 - 0.56) | Ref |  | 0.74 (0.59 - 0.92) | 0.59 (0.57 - 0.61) | 1.06 (0.99 - 1.14) | Ref |  | 0.54 (0.51 - 0.57) | 0.74 (0.71 - 0.78) | 0.91 (0.84 - 0.99) | Ref |
| Age adjusted HR (95% CI) |  |  | 0.83 (0.78 - 0.88) | 0.72 (0.68 - 0.77) | Ref |  |  | 0.48 (0.46 - 0.50) | 0.55 (0.52 - 0.59) | Ref |  | 0.59 (0.47 - 0.73) | 0.54 (0.52 - 0.56) | 1.15 (1.07 - 1.24) | Ref |  | 0.44 (0.42 - 0.46) | 0.80 (0.76 - 0.83) | 0.95 (0.88 - 1.03) | Ref |
| Age and sex adjusted HR (95% CI) |  |  | 0.84 (0.79 - 0.89) | 0.72 (0.68 - 0.77) | Ref |  |  | 0.47 (0.46 - 0.49) | 0.54 (0.51 - 0.57) | Ref |  | 0.58 (0.46 - 0.72) | 0.53 (0.51 - 0.55) | 1.13 (1.05 - 1.21) | Ref |  | 0.43 (0.41 - 0.45) | 0.77 (0.74 - 0.81) | 0.93 (0.86 - 1.01) | Ref |
| Age, sex and education adjusted HR (95% CI) |  |  | 0.84 (0.79 - 0.89) | 0.72 (0.67 - 0.77) | Ref |  |  | 0.47 (0.45 - 0.49) | 0.54 (0.51 - 0.57) | Ref |  | 0.56 (0.45 - 0.70) | 0.52 (0.50 - 0.55) | 1.11 (1.03 - 1.19) | Ref |  | 0.43 (0.41 - 0.45) | 0.76 (0.73 - 0.80) | 0.92 (0.85 - 1.00) | Ref |
|  | 2022 | | | | | | | | | | | | | | | | | | | |
| Deaths (n) | 14 | 7820 | 2220 | 408 | 2039 | 156 | 6711 | 1343 | 163 | 1289 | 716 | 4038 | 679 | 76 | 859 |  |  |  |  | 0 |
| Events per 100,000 person days | 21.74 | 26.29 | 46.71 | 61.13 | 48.10 | 24.05 | 25.73 | 41.18 | 55.15 | 39.09 | 24.02 | 28.81 | 40.15 | 43.41 | 42.08 |  |  |  |  |  |
| Crude HR (95%CI) | 0.45 (0.27 - 0.76) | 0.55 (0.52 - 0.57) | 0.97 (0.91 - 1.03) | 1.28 (1.15 - 1.43) | Ref | 0.64 (0.54 - 0.76) | 0.66 (0.62 - 0.70) | 1.05 (0.97 - 1.13) | 1.41 (1.20 - 1.66) | Ref | 0.57 (0.52 - 0.63) | 0.69 (0.64 - 0.74) | 0.96 (0.87 - 1.06) | 1.00 (0.79 - 1.27) | Ref |  |  |  |  | Ref |
| Age adjusted HR (95% CI) | 0.47 (0.28 - 0.79) | 0.49 (0.47 - 0.52) | 1.00 (0.94 - 1.06) | 1.23 (1.11 - 1.37) | Ref | 0.50 (0.42 - 0.60) | 0.61 (0.57 - 0.64) | 1.05 (0.97 - 1.13) | 1.44 (1.22 - 1.69) | Ref | 0.45 (0.41 - 0.50) | 0.65 (0.60 - 0.70) | 0.96 (0.87 - 1.06) | 1.05 (0.83 - 1.33) | Ref |  |  |  |  | Ref |
| Age and sex adjusted HR (95% CI) | 0.43 (0.25 - 0.73) | 0.47 (0.45 - 0.50) | 0.98 (0.92 - 1.04) | 1.21 (1.09 - 1.34) | Ref | 0.47 (0.40 - 0.56) | 0.58 (0.55 - 0.62) | 1.02 (0.95 - 1.10) | 1.40 (1.19 - 1.65) | Ref | 0.43 (0.39 - 0.48) | 0.62 (0.58 - 0.67) | 0.94 (0.85 - 1.04) | 1.03 (0.81 - 1.30) | Ref |  |  |  |  | Ref |
| Age, sex and education adjusted HR (95% CI) | 0.42 (0.25 - 0.72) | 0.47 (0.45 - 0.50) | 0.97 (0.91 - 1.03) | 1.20 (1.08 - 1.34) | Ref | 0.48 (0.40 - 0.57) | 0.58 (0.54 - 0.61) | 1.02 (0.94 - 1.10) | 1.39 (1.18 - 1.64) | Ref | 0.43 (0.39 - 0.48) | 0.62 (0.57 - 0.67) | 0.94 (0.85 - 1.04) | 1.03 (0.81 - 1.30) | Ref |  |  |  |  | Ref |
| Estimates with less than 10 deaths in either the vaccinated or reference group are not shown | | | | | | | | | | | | | | | | | | | | |

| **Table S29:** Hazard ratios (HR) with 95% confidence intervals (95% CI) for all-cause mortality according to number of SARS-CoV-2 vaccine doses for the period from 2021 to 2022 split into 3 month intervals for non-ALTCA recipients only. | | | | | | | | | | | | | | | | | | | | |
| --- | --- | --- | --- | --- | --- | --- | --- | --- | --- | --- | --- | --- | --- | --- | --- | --- | --- | --- | --- | --- |
|  | 2021 | | | | | | | | | | | | | | | | | | | |
|  | All-cause mortality | | | | | | | | | | | | | | | | | | | |
|  | first quarter | | | | | second quarter | | | | | third quarter | | | | | fourth quarter | | | | |
|  | Four vaccine doses | Three vaccine doses | Two vaccine doses | One vaccine dose | Unvaccinated | Four vaccine doses | Three vaccine doses | Two vaccine doses | One vaccine dose | Unvaccinated | Four vaccine doses | Three vaccine doses | Two vaccine doses | One vaccine dose | Unvaccinated | Four vaccine doses | Three vaccine doses | Two vaccine doses | One vaccine dose | Unvaccinated |
| Deaths (n) |  |  | 86 | 199 | 5449 |  |  | 1451 | 1003 | 3200 |  | 8 | 3468 | 471 | 1978 |  | 1134 | 3153 | 302 | 1711 |
| Events per 100,000 person days |  |  | 0.85 | 1.08 | 0.92 |  |  | 1.48 | 0.74 | 0.85 |  |  | 0.97 | 0.85 | 1.00 |  | 1.03 | 1.03 | 0.94 | 1.21 |
| Crude HR (95%CI) |  |  | 0.94 (0.76 - 1.16) | 1.20 (1.03 - 1.38) | Ref |  |  | 1.77 (1.66 - 1.90) | 0.87 (0.81 - 0.94) | Ref |  |  | 0.96 (0.91 - 1.02) | 0.88 (0.79 - 0.97) | Ref |  | 0.87 (0.80 - 0.95) | 0.86 (0.81 - 0.91) | 0.77 (0.68 - 0.87) | Ref |
| Age adjusted HR (95% CI) |  |  | 0.44 (0.36 - 0.55) | 0.51 (0.44 - 0.59) | Ref |  |  | 0.38 (0.35 - 0.41) | 0.41 (0.38 - 0.44) | Ref |  |  | 0.43 (0.40 - 0.45) | 1.07 (0.97 - 1.19) | Ref |  | 0.26 (0.23 - 0.28) | 0.61 (0.57 - 0.64) | 0.96 (0.85 - 1.08) | Ref |
| Age and sex adjusted HR (95% CI) |  |  | 0.44 (0.35 - 0.55) | 0.51 (0.44 - 0.59) | Ref |  |  | 0.37 (0.34 - 0.39) | 0.40 (0.37 - 0.43) | Ref |  |  | 0.41 (0.39 - 0.44) | 1.04 (0.94 - 1.15) | Ref |  | 0.24 (0.22 - 0.27) | 0.59 (0.55 - 0.63) | 0.93 (0.82 - 1.05) | Ref |
| Age, sex and education adjusted HR (95% CI) |  |  | 0.45 (0.37 - 0.56) | 0.52 (0.45 - 0.60) | Ref |  |  | 0.37 (0.35 - 0.40) | 0.41 (0.38 - 0.44) | Ref |  |  | 0.42 (0.40 - 0.45) | 1.03 (0.93 - 1.14) | Ref |  | 0.24 (0.22 - 0.27) | 0.58 (0.54 - 0.62) | 0.88 (0.78 - 1.00) | Ref |
|  | 2022 | | | | | | | | | | | | | | | | | | | |
| Deaths (n) | 11 | 3241 | 979 | 159 | 1135 | 43 | 3213 | 610 | 75 | 825 | 215 | 2023 | 323 | 46 | 534 |  |  |  |  | 0 |
| Events per 100,000 person days | 4.65 | 1.05 | 1.1 | 2 | 1.32 | 2.27 | 1.2 | 1.21 | 1.63 | 1.26 | 1.73 | 1.32 | 1.14 | 1.58 | 1.28 |  |  |  |  |  |
| Crude HR (95%CI) | 3.44 (1.90 - 6.23) | 0.79 (0.74 - 0.84) | 0.84 (0.77 - 0.92) | 1.52 (1.29 - 1.79) | Ref | 1.80 (1.32 - 2.46) | 0.96 (0.89 - 1.03) | 0.96 (0.87 - 1.07) | 1.30 (1.02 - 1.64) | Ref | 1.31 (1.12 - 1.54) | 1.04 (0.94 - 1.14) | 0.90 (0.78 - 1.03) | 1.20 (0.89 - 1.62) | Ref |  |  |  |  | Ref |
| Age adjusted HR (95% CI) | 1.09 (0.60 - 1.97) | 0.40 (0.37 - 0.42) | 1.05 (0.96 - 1.14) | 1.60 (1.36 - 1.89) | Ref | 0.32 (0.23 - 0.44) | 0.52 (0.48 - 0.56) | 1.13 (1.02 - 1.26) | 1.52 (1.20 - 1.92) | Ref | 0.30 (0.25 - 0.35) | 0.61 (0.56 - 0.68) | 1.08 (0.94 - 1.24) | 1.34 (0.99 - 1.81) | Ref |  |  |  |  | Ref |
| Age and sex adjusted HR (95% CI) | 0.98 (0.54 - 1.77) | 0.38 (0.36 - 0.41) | 1.03 (0.94 - 1.12) | 1.58 (1.34 - 1.87) | Ref | 0.30 (0.22 - 0.41) | 0.51 (0.47 - 0.55) | 1.11 (1.00 - 1.24) | 1.50 (1.18 - 1.90) | Ref | 0.28 (0.24 - 0.33) | 0.60 (0.55 - 0.66) | 1.06 (0.92 - 1.22) | 1.36 (1.00 - 1.84) | Ref |  |  |  |  | Ref |
| Age, sex and education adjusted HR (95% CI) | 1.07 (0.59 - 1.95) | 0.38 (0.35 - 0.40) | 0.96 (0.88 - 1.05) | 1.49 (1.26 - 1.76) | Ref | 0.32 (0.23 - 0.44) | 0.50 (0.46 - 0.54) | 1.06 (0.96 - 1.18) | 1.41 (1.11 - 1.79) | Ref | 0.29 (0.24 - 0.35) | 0.59 (0.54 - 0.65) | 1.00 (0.87 - 1.15) | 1.25 (0.91 - 1.70) | Ref |  |  |  |  | Ref |
| Estimates with less than 10 deaths in either the vaccinated or reference group are not shown | | | | | | | | | | | | | | | | | | | | |

| **Table S30:** Hazard ratios (HR) with 95% confidence intervals (95% CI) for all-cause mortality according to number of SARS-CoV-2 vaccine doses for the period from 2021 to 2022 split into 3 month intervals for mRNA vaccines (last adiministered dose) only. | | | | | | | | | | | | | | | | | | | | |
| --- | --- | --- | --- | --- | --- | --- | --- | --- | --- | --- | --- | --- | --- | --- | --- | --- | --- | --- | --- | --- |
|  | 2021 | | | | | | | | | | | | | | | | | | | |
|  | All-cause mortality | | | | | | | | | | | | | | | | | | | |
|  | first quarter | | | | | second quarter | | | | | third quarter | | | | | fourth quarter | | | | |
|  | Four vaccine doses | Three vaccine doses | Two vaccine doses | One vaccine dose | Unvaccinated | Four vaccine doses | Three vaccine doses | Two vaccine doses | One vaccine dose | Unvaccinated | Four vaccine doses | Three vaccine doses | Two vaccine doses | One vaccine dose | Unvaccinated | Four vaccine doses | Three vaccine doses | Two vaccine doses | One vaccine dose | Unvaccinated |
| Deaths (n) |  |  | 1374 | 1343 | 15583 |  | 1 | 6834 | 1819 | 8823 |  | 98 | 10610 | 831 | 5842 | 1 | 5360 | 8095 | 526 | 5013 |
| Events per 100,000 person days |  |  | 9.89 | 9.2 | 2.50 |  |  | 6.45 | 1.99 | 2.26 |  | 12.81 | 3.27 | 2.43 | 2.84 |  | 4.3 | 2.88 | 3.43 | 3.38 |
| Crude HR (95%CI) |  |  | 4.32 (4.07 - 4.57) | 3.90 (3.69 - 4.13) | Ref |  |  | 2.97 (2.87 - 3.07) | 0.89 (0.84 - 0.94) | Ref |  | 4.38 (3.57 - 5.38) | 1.16 (1.12 - 1.19) | 0.91 (0.85 - 0.99) | Ref |  | 1.54 (1.47 - 1.61) | 0.85 (0.82 - 0.89) | 0.98 (0.90 - 1.07) | Ref |
| Age adjusted HR (95% CI) |  |  | 0.96 (0.90 - 1.02) | 0.78 (0.73 - 0.83) | Ref |  |  | 0.47 (0.45 - 0.48) | 0.51 (0.49 - 0.54) | Ref |  | 0.63 (0.51 - 0.77) | 0.49 (0.47 - 0.51) | 1.28 (1.19 - 1.38) | Ref |  | 0.37 (0.36 - 0.39) | 0.71 (0.69 - 0.74) | 0.99 (0.90 - 1.08) | Ref |
| Age and gender adjusted HR (95% CI) |  |  | 0.98 (0.92 - 1.04) | 0.78 (0.73 - 0.82) | Ref |  |  | 0.45 (0.44 - 0.47) | 0.50 (0.48 - 0.53) | Ref |  | 0.61 (0.50 - 0.75) | 0.48 (0.46 - 0.49) | 1.27 (1.18 - 1.37) | Ref |  | 0.36 (0.34 - 0.37) | 0.69 (0.67 - 0.72) | 0.98 (0.89 - 1.07) | Ref |
| Age, sex and ALTCA adjusted HR (95% CI) |  |  | 0.75 (0.71 - 0.80) | 0.68 (0.64 - 0.72) | Ref |  |  | 0.47 (0.45 - 0.48) | 0.60 (0.57 - 0.63) | Ref |  | 0.51 (0.41 - 0.63) | 0.52 (0.51 - 0.54) | 1.23 (1.14 - 1.32) | Ref |  | 0.38 (0.37 - 0.40) | 0.74 (0.71 - 0.76) | 0.90 (0.82 - 0.99) | Ref |
| Age, sex, ALTCA and education adjusted HR (95% CI) |  |  | 0.75 (0.71 - 0.80) | 0.68 (0.65 - 0.72) | Ref |  |  | 0.46 (0.45 - 0.48) | 0.59 (0.56 - 0.62) | Ref |  | 0.51 (0.42 - 0.63) | 0.52 (0.50 - 0.54) | 1.22 (1.13 - 1.31) | Ref |  | 0.38 (0.37 - 0.40) | 0.73 (0.70 - 0.75) | 0.88 (0.81 - 0.97) | Ref |
|  | 2022 | | | | | | | | | | | | | | | | | | | |
| Deaths (n) | 25 | 11053 | 3002 | 443 | 3174 | 199 | 9916 | 1890 | 193 | 2114 | 926 | 6054 | 971 | 101 | 1393 |  |  |  |  | 0 |
| Events per 100,000 person days | 8.35 | 3.28 | 3.3 | 6.55 | 3.52 | 7.87 | 3.39 | 3.61 | 5.02 | 3.07 | 6.02 | 3.61 | 3.31 | 4.14 | 3.18 |  |  |  |  |  |
| Crude HR (95%CI) | 2.33 (1.57 - 3.45) | 0.93 (0.89 - 0.96) | 0.95 (0.90 - 1.00) | 1.87 (1.69 - 2.07) | Ref | 2.64 (2.27 - 3.06) | 1.10 (1.05 - 1.16) | 1.17 (1.10 - 1.25) | 1.64 (1.41 - 1.90) | Ref | 1.87 (1.71 - 2.03) | 1.14 (1.07 - 1.21) | 1.05 (0.97 - 1.14) | 1.28 (1.04 - 1.56) | Ref |  |  |  |  |  |
| Age adjusted HR (95% CI) | 0.72 (0.48 - 1.06) | 0.45 (0.43 - 0.47) | 1.10 (1.05 - 1.16) | 1.38 (1.25 - 1.52) | Ref | 0.45 (0.39 - 0.52) | 0.57 (0.55 - 0.60) | 1.22 (1.14 - 1.30) | 1.60 (1.38 - 1.85) | Ref | 0.38 (0.35 - 0.41) | 0.64 (0.61 - 0.68) | 1.13 (1.04 - 1.23) | 1.29 (1.06 - 1.58) | Ref |  |  |  |  |  |
| Age and gender adjusted HR (95% CI) | 0.66 (0.44 - 0.98) | 0.44 (0.42 - 0.45) | 1.09 (1.03 - 1.14) | 1.37 (1.24 - 1.52) | Ref | 0.43 (0.37 - 0.50) | 0.55 (0.53 - 0.58) | 1.20 (1.13 - 1.28) | 1.58 (1.37 - 1.83) | Ref | 0.37 (0.33 - 0.40) | 0.63 (0.59 - 0.66) | 1.12 (1.03 - 1.21) | 1.29 (1.05 - 1.58) | Ref |  |  |  |  |  |
| Age, sex and ALTCA adjusted HR (95% CI) | 0.60 (0.41 - 0.89) | 0.45 (0.43 - 0.47) | 0.95 (0.91 - 1.00) | 1.17 (1.06 - 1.30) | Ref | 0.43 (0.37 - 0.50) | 0.57 (0.54 - 0.59) | 1.03 (0.97 - 1.10) | 1.40 (1.21 - 1.63) | Ref | 0.39 (0.36 - 0.42) | 0.63 (0.59 - 0.66) | 0.96 (0.88 - 1.04) | 1.17 (0.95 - 1.43) | Ref |  |  |  |  | Ref |
| Age, sex, ALTCA and education adjusted HR (95% CI) | 0.62 (0.42 - 0.93) | 0.45 (0.43 - 0.46) | 0.93 (0.89 - 0.98) | 1.16 (1.05 - 1.28) | Ref | 0.44 (0.38 - 0.52) | 0.56 (0.53 - 0.59) | 1.01 (0.95 - 1.08) | 1.36 (1.18 - 1.58) | Ref | 0.39 (0.36 - 0.43) | 0.61 (0.58 - 0.65) | 0.94 (0.86 - 1.02) | 1.14 (0.93 - 1.40) | Ref |  |  |  |  | Ref |
| Estimates with less than 10 deaths in either the vaccinated or reference group are not shown | | | | | | | | | | | | | | | | | | | | |

| **Table S31:** Hazard ratios (HR) with 95% confidence intervals (95% CI) for all-cause mortality according to number of SARS-CoV-2 vaccine doses for the period from 2021 to 2022 split into 3 month intervals for non-mRNA vaccines (last adiministered dose) only. | | | | | | | | | | | | | | | | | | | | |
| --- | --- | --- | --- | --- | --- | --- | --- | --- | --- | --- | --- | --- | --- | --- | --- | --- | --- | --- | --- | --- |
|  | 2021 | | | | | | | | | | | | | | | | | | | |
|  | All-cause mortality | | | | | | | | | | | | | | | | | | | |
|  | first quarter | | | | | second quarter | | | | | third quarter | | | | | fourth quarter | | | | |
|  | Four vaccine doses | Three vaccine doses | Two vaccine doses | One vaccine dose | Unvaccinated | Four vaccine doses | Three vaccine doses | Two vaccine doses | One vaccine dose | Unvaccinated | Four vaccine doses | Three vaccine doses | Two vaccine doses | One vaccine dose | Unvaccinated | Four vaccine doses | Three vaccine doses | Two vaccine doses | One vaccine dose | Unvaccinated |
| Deaths (n) |  |  |  | 28 | 15583 |  |  | 76 | 727 | 8823 |  | 2 | 973 | 557 | 5842 |  | 8 | 1073 | 448 | 5013 |
| Events per 100,000 person days |  |  |  | 0.35 | 2.50 |  |  | 0.58 | 1.43 | 2.26 |  |  | 1.51 | 2.44 | 2.84 |  |  | 2.57 | 2.47 | 3.38 |
| Crude HR (95%CI) |  |  |  | 0.15 (0.11 - 0.22) | Ref |  |  | 0.23 (0.18 - 0.28) | 0.63 (0.58 - 0.67) | Ref |  |  | 0.53 (0.50 - 0.57) | 0.86 (0.79 - 0.94) | Ref |  |  | 0.80 (0.75 - 0.86) | 0.74 (0.67 - 0.82) | Ref |
| Age adjusted HR (95% CI) |  |  |  | 0.26 (0.18 - 0.38) | Ref |  |  | 0.18 (0.14 - 0.22) | 0.32 (0.30 - 0.34) | Ref |  |  | 0.32 (0.30 - 0.34) | 1.04 (0.95 - 1.14) | Ref |  |  | 0.53 (0.49 - 0.57) | 1.11 (1.00 - 1.22) | Ref |
| Age and gender adjusted HR (95% CI) |  |  |  | 0.26 (0.18 - 0.38) | Ref |  |  | 0.17 (0.14 - 0.22) | 0.31 (0.29 - 0.33) | Ref |  |  | 0.31 (0.29 - 0.33) | 1.01 (0.92 - 1.10) | Ref |  |  | 0.51 (0.48 - 0.55) | 1.07 (0.97 - 1.18) | Ref |
| Age, sex and ALTCA adjusted HR (95% CI) |  |  |  | 0.27 (0.19 - 0.40) | Ref |  |  | 0.22 (0.17 - 0.28) | 0.39 (0.36 - 0.42) | Ref |  |  | 0.41 (0.38 - 0.44) | 0.95 (0.87 - 1.03) | Ref |  |  | 0.63 (0.59 - 0.67) | 0.98 (0.89 - 1.08) | Ref |
| Age, sex, ALTCA and education adjusted HR (95% CI) |  |  |  | 0.28 (0.19 - 0.40) | Ref |  |  | 0.22 (0.17 - 0.28) | 0.39 (0.36 - 0.42) | Ref |  |  | 0.41 (0.38 - 0.44) | 0.93 (0.86 - 1.02) | Ref |  |  | 0.62 (0.58 - 0.67) | 0.96 (0.87 - 1.06) | Ref |
|  | 2022 | | | | | | | | | | | | | | | | | | | |
| Deaths (n) |  | 8 | 197 | 124 | 3174 |  | 8 | 63 | 45 | 2114 | 5 | 7 | 31 | 21 | 1393 |  |  |  |  | 0 |
| Events per 100,000 person days |  |  | 8.22 | 6.68 | 3.52 |  |  | 4.88 | 4.28 | 3.07 |  |  | 4.12 | 3.24 | 3.18 |  |  |  |  |  |
| Crude HR (95%CI) |  |  | 2.36 (2.04 - 2.72) | 1.91 (1.60 - 2.29) | Ref |  |  | 1.59 (1.24 - 2.04) | 1.39 (1.04 - 1.87) | Ref |  |  | 1.32 (0.92 - 1.88) | 0.94 (0.61 - 1.45) | Ref |  |  |  |  |  |
| Age adjusted HR (95% CI) |  |  | 2.12 (1.83 - 2.45) | 2.37 (1.98 - 2.84) | Ref |  |  | 1.40 (1.09 - 1.79) | 1.82 (1.35 - 2.44) | Ref |  |  | 1.14 (0.80 - 1.62) | 1.26 (0.82 - 1.94) | Ref |  |  |  |  |  |
| Age and gender adjusted HR (95% CI) |  |  | 2.06 (1.78 - 2.38) | 2.33 (1.95 - 2.79) | Ref |  |  | 1.37 (1.07 - 1.76) | 1.81 (1.34 - 2.43) | Ref |  |  | 1.12 (0.78 - 1.60) | 1.23 (0.80 - 1.89) | Ref |  |  |  |  |  |
| Age, sex and ALTCA adjusted HR (95% CI) |  |  | 1.83 (1.58 - 2.11) | 1.95 (1.63 - 2.34) | Ref |  |  | 1.26 (0.98 - 1.62) | 1.55 (1.16 - 2.09) | Ref |  |  | 1.02 (0.72 - 1.46) | 1.02 (0.66 - 1.58) | Ref |  |  |  |  | Ref |
| Age, sex, ALTCA and education adjusted HR (95% CI) |  |  | 1.81 (1.56 - 2.09) | 1.93 (1.61 - 2.31) | Ref |  |  | 1.25 (0.97 - 1.61) | 1.54 (1.15 - 2.07) | Ref |  |  | 1.01 (0.71 - 1.44) | 1.01 (0.65 - 1.55) | Ref |  |  |  |  | Ref |
| Estimates with less than 10 deaths in either the vaccinated or reference group are not shown | | | | | | | | | | | | | | | | | | | | |

| **Table S32:** Hazard ratios (HR) with 95% confidence intervals (95% CI) for all-cause mortality according to number of SARS-CoV-2 vaccine doses during different time periods for males only. | | | | | | | | | | |
| --- | --- | --- | --- | --- | --- | --- | --- | --- | --- | --- |
|  | June and July 2021 (low COVID-19 disease burden) | | | | | October and November 2021 (high COVID-19 disease burden) | | | | |
|  | Four vaccine doses | Three vaccine doses | Two vaccine doses | One vaccine dose | Unvaccinated | Four vaccine doses | Three vaccine doses | Two vaccine doses | One vaccine dose | Unvaccinated |
|  | All-cause mortality | | | | | All-cause mortality | | | | |
| Deaths (n) |  |  | 3068 | 886 | 2290 |  | 786 | 4000 | 349 | 1631 |
| Events per 100.000 person days |  |  | 4.98 | 1.66 | 2.68 |  | 13.09 | 3.08 | 2.58 | 3.45 |
| Crude HR (95%CI) |  |  | 1.99 (1.88 - 2.10) | 0.62 (0.57 - 0.67) | Ref |  | 3.86 (3.52 - 4.24) | 0.89 (0.84 - 0.95) | 0.74 (0.66 - 0.83) | Ref |
| Age adjusted HR (95% CI) |  |  | 0.45 (0.43 - 0.48) | 0.50 (0.46 - 0.54) | Ref |  | 0.53 (0.49 - 0.59) | 0.50 (0.47 - 0.53) | 1.03 (0.92 - 1.16) | Ref |
| Age and ALTCA adjusted HR (95% CI) |  |  | 0.51 (0.48 - 0.54) | 0.59 (0.55 - 0.64) | Ref |  | 0.52 (0.47 - 0.57) | 0.58 (0.54 - 0.61) | 0.94 (0.84 - 1.06) | Ref |
| Age, ALTCA and education adjusted HR (95% CI) |  |  | 0.52 (0.49 - 0.55) | 0.60 (0.55 - 0.65) | Ref |  | 0.53 (0.48 - 0.58) | 0.58 (0.55 - 0.62) | 0.95 (0.84 - 1.06) | Ref |
|  | May and June 2022 (low COVID-19 disease burden) | | | | | February and March 2022 (high COVID-19 disease burden) | | | | |
|  | Four vaccine doses | Three vaccine doses | Two vaccine doses | One vaccine dose | Unvaccinated | Four vaccine doses | Three vaccine doses | Two vaccine doses | One vaccine dose | Unvaccinated |
|  | All-cause mortality | | | | | All-cause mortality | | | | |
| Deaths (n) | 75 | 3412 | 595 | 71 | 582 | 7 | 3717 | 982 | 145 | 848 |
| Events per 100.000 person days | 10.63 | 3.66 | 3.08 | 4.25 | 3.37 |  | 3.6 | 3.49 | 5.95 | 4.24 |
| Crude HR (95%CI) | 3.14 (2.46 - 4.01) | 1.09 (1.00 - 1.19) | 0.92 (0.82 - 1.03) | 1.26 (0.99 - 1.61) | Ref |  | 0.85 (0.79 - 0.91) | 0.82 (0.75 - 0.90) | 1.41 (1.18 - 1.68) | Ref |
| Age adjusted HR (95% CI) | 0.62 (0.48 - 0.80) | 0.64 (0.58 - 0.70) | 1.28 (1.15 - 1.44) | 1.74 (1.36 - 2.23) | Ref |  | 0.47 (0.44 - 0.51) | 1.21 (1.10 - 1.32) | 1.64 (1.37 - 1.95) | Ref |
| Age and ALTCA adjusted HR (95% CI) | 0.59 (0.46 - 0.75) | 0.64 (0.59 - 0.70) | 1.05 (0.94 - 1.18) | 1.45 (1.13 - 1.86) | Ref |  | 0.49 (0.45 - 0.53) | 1.01 (0.92 - 1.11) | 1.31 (1.09 - 1.56) | Ref |
| Age, ALTCA and education adjusted HR (95% CI) | 0.61 (0.48 - 0.79) | 0.64 (0.59 - 0.70) | 1.05 (0.94 - 1.18) | 1.41 (1.10 - 1.81) | Ref |  | 0.49 (0.46 - 0.53) | 1.01 (0.92 - 1.10) | 1.30 (1.09 - 1.55) | Ref |
| Estimates with less than 10 deaths in either the vaccinated or reference group are not shown | | | | | | | | | | |

| **Table S33:** Hazard ratios (HR) with 95% confidence intervals (95% CI) for all-cause mortality according to number of SARS-CoV-2 vaccine doses during different time periods for females only. | | | | | | | | | | |
| --- | --- | --- | --- | --- | --- | --- | --- | --- | --- | --- |
|  | June and July 2021 (low COVID-19 disease burden) | | | | | October and November 2021 (high COVID-19 disease burden) | | | | |
|  | Four vaccine doses | Three vaccine doses | Two vaccine doses | One vaccine dose | Unvaccinated | Four vaccine doses | Three vaccine doses | Two vaccine doses | One vaccine dose | Unvaccinated |
|  | All-cause mortality | | | | | All-cause mortality | | | | |
| Deaths (n) |  | 1 | 3099 | 711 | 2240 |  | 1002 | 3424 | 337 | 1878 |
| Events per 100.000 person days |  |  | 4.13 | 1.4 | 2.64 |  | 12.37 | 2.47 | 3.28 | 3.81 |
| Crude HR (95%CI) |  |  | 1.65 (1.56 - 1.74) | 0.54 (0.49 - 0.58) | Ref |  | 3.33 (3.06 - 3.62) | 0.65 (0.61 - 0.69) | 0.85 (0.76 - 0.96) | Ref |
| Age adjusted HR (95% CI) |  |  | 0.50 (0.47 - 0.52) | 0.61 (0.56 - 0.66) | Ref |  | 0.59 (0.54 - 0.64) | 0.51 (0.48 - 0.54) | 1.14 (1.02 - 1.28) | Ref |
| Age and ALTCA adjusted HR (95% CI) |  |  | 0.52 (0.50 - 0.55) | 0.71 (0.65 - 0.77) | Ref |  | 0.54 (0.50 - 0.59) | 0.56 (0.52 - 0.59) |  | Ref |
| Age, ALTCA and education adjusted HR (95% CI) |  |  | 0.52 (0.50 - 0.55) | 0.71 (0.65 - 0.77) | Ref |  | 0.55 (0.50 - 0.59) | 0.56 (0.53 - 0.59) |  | Ref |
|  | May and June 2022 (low COVID-19 disease burden) | | | | | February and March 2022 (high COVID-19 disease burden) | | | | |
|  | Four vaccine doses | Three vaccine doses | Two vaccine doses | One vaccine dose | Unvaccinated | Four vaccine doses | Three vaccine doses | Two vaccine doses | One vaccine dose | Unvaccinated |
|  | All-cause mortality | | | | | All-cause mortality | | | | |
| Deaths (n) | 68 | 3103 | 627 | 68 | 729 | 11 | 3434 | 888 | 158 | 1064 |
| Events per 100.000 person days | 8.97 | 3.13 | 3.96 | 4.92 | 4.05 | 12.35 | 3.07 | 3.57 | 7.11 | 5.16 |
| Crude HR (95%CI) | 2.18 (1.70 - 2.80) | 0.77 (0.71 - 0.84) | 0.98 (0.88 - 1.09) | 1.22 (0.95 - 1.56) | Ref | 2.36 (1.30 - 4.27) | 0.59 (0.55 - 0.64) | 0.69 (0.64 - 0.76) | 1.39 (1.17 - 1.64) | Ref |
| Age adjusted HR (95% CI) | 0.59 (0.46 - 0.76) | 0.59 (0.55 - 0.64) | 1.23 (1.10 - 1.37) | 1.44 (1.13 - 1.85) | Ref | 1.31 (0.72 - 2.37) | 0.47 (0.44 - 0.51) | 1.02 (0.94 - 1.12) | 1.37 (1.16 - 1.62) | Ref |
| Age and ALTCA adjusted HR (95% CI) | 0.57 (0.44 - 0.73) | 0.58 (0.53 - 0.63) | 1.07 (0.96 - 1.19) | 1.32 (1.03 - 1.69) | Ref | 1.14 (0.63 - 2.06) | 0.47 (0.43 - 0.50) | 0.90 (0.83 - 0.99) | 1.19 (1.01 - 1.41) | Ref |
| Age, ALTCA and education adjusted HR (95% CI) | 0.58 (0.45 - 0.75) | 0.58 (0.54 - 0.63) | 1.08 (0.97 - 1.20) | 1.33 (1.03 - 1.70) | Ref | 1.15 (0.63 - 2.08) | 0.47 (0.43 - 0.50) | 0.90 (0.83 - 0.99) | 1.19 (1.01 - 1.41) | Ref |
| Estimates with less than 10 deaths in either the vaccinated or reference group are not shown | | | | | | | | | | |

| **Table S34:** Hazard ratios (HR) with 95% confidence intervals (95% CI) for all-cause mortality according to number of SARS-CoV-2 vaccine doses during different time periods for 18-39 year olds only. | | | | | | | | | | |
| --- | --- | --- | --- | --- | --- | --- | --- | --- | --- | --- |
|  | June and July 2021 (low COVID-19 disease burden) | | | | | October and November 2021 (high COVID-19 disease burden) | | | | |
|  | Four vaccine doses | Three vaccine doses | Two vaccine doses | One vaccine dose | Unvaccinated | Four vaccine doses | Three vaccine doses | Two vaccine doses | One vaccine dose | Unvaccinated |
|  | All-cause mortality | | | | | All-cause mortality | | | | |
| Deaths (n) |  |  | 27 | 29 | 154 |  | 2 | 78 | 20 | 77 |
| Events per 100.000 person days |  |  | 0.12 | 0.09 | 0.19 |  |  | 0.1 | 0.16 | 0.18 |
| Crude HR (95%CI) |  |  | 0.65 (0.43 - 1.00) | 0.47 (0.31 - 0.70) | Ref |  |  | 0.57 (0.42 - 0.78) | 0.91 (0.56 - 1.49) | Ref |
| Age adjusted HR (95% CI) |  |  | 0.61 (0.40 - 0.94) | 0.45 (0.30 - 0.67) | Ref |  |  | 0.56 (0.41 - 0.77) | 0.97 (0.59 - 1.59) | Ref |
| Age and gender adjusted HR (95% CI) |  |  | 0.65 (0.43 - 0.99) | 0.44 (0.30 - 0.66) | Ref |  |  | 0.56 (0.41 - 0.77) | 0.92 (0.56 - 1.50) | Ref |
| Age, sex and ALTCA adjusted HR (95% CI) |  |  | 0.49 (0.32 - 0.76) | 0.46 (0.31 - 0.68) | Ref |  |  | 0.52 (0.38 - 0.71) | 0.96 (0.58 - 1.57) | Ref |
| Age, sex, ALTCA and education adjusted HR (95% CI) |  |  | 0.60 (0.39 - 0.94) | 0.56 (0.37 - 0.84) | Ref |  |  | 0.58 (0.42 - 0.80) | 0.98 (0.60 - 1.60) | Ref |
|  | May and June 2022 (low COVID-19 disease burden) | | | | | February and March 2022 (high COVID-19 disease burden) | | | | |
|  | Four vaccine doses | Three vaccine doses | Two vaccine doses | One vaccine dose | Unvaccinated | Four vaccine doses | Three vaccine doses | Two vaccine doses | One vaccine dose | Unvaccinated |
|  | All-cause mortality | | | | | All-cause mortality | | | | |
| Deaths (n) | 1 | 63 | 39 | 5 | 27 | 1 | 55 | 67 | 4 | 44 |
| Events per 100.000 person days |  | 0.15 | 0.24 |  | 0.21 |  | 0.11 | 0.26 |  | 0.30 |
| Crude HR (95%CI) |  | 0.71 (0.45 - 1.11) | 1.13 (0.69 - 1.84) |  | Ref |  | 0.37 (0.25 - 0.56) | 0.87 (0.60 - 1.28) |  | Ref |
| Age adjusted HR (95% CI) |  | 0.70 (0.45 - 1.10) | 1.19 (0.73 - 1.94) |  | Ref |  | 0.37 (0.25 - 0.56) | 0.88 (0.60 - 1.30) |  | Ref |
| Age and gender adjusted HR (95% CI) |  | 0.70 (0.45 - 1.10) | 1.13 (0.69 - 1.86) |  | Ref |  | 0.38 (0.25 - 0.56) | 0.87 (0.59 - 1.28) |  | Ref |
| Age, sex and ALTCA adjusted HR (95% CI) |  | 0.64 (0.41 - 1.01) | 1.18 (0.72 - 1.92) |  | Ref |  | 0.35 (0.24 - 0.52) | 0.91 (0.62 - 1.34) |  | Ref |
| Age, sex, ALTCA and education adjusted HR (95% CI) |  | 0.66 (0.42 - 1.05) | 1.10 (0.67 - 1.81) |  | Ref |  | 0.42 (0.28 - 0.64) | 0.91 (0.62 - 1.34) |  | Ref |
| Estimates with less than 10 deaths in either the vaccinated or reference group are not shown | | | | | | | | | | |

| **Table S35:** Hazard ratios (HR) with 95% confidence intervals (95% CI) for all-cause mortality according to number of SARS-CoV-2 vaccine doses during different time periods for 40-59 year olds only. | | | | | | | | | | | | | | | | | | | | |
| --- | --- | --- | --- | --- | --- | --- | --- | --- | --- | --- | --- | --- | --- | --- | --- | --- | --- | --- | --- | --- |
|  | June and July 2021 (low COVID-19 disease burden) | | | | | | | | | | October and November 2021 (high COVID-19 disease burden) | | | | | | | | | |
|  | Four vaccine doses | | Three vaccine doses | | Two vaccine doses | | One vaccine dose | | Unvaccinated | | Four vaccine doses | | Three vaccine doses | | Two vaccine doses | | One vaccine dose | | Unvaccinated | |
|  | All-cause mortality | | | | | | | | | | All-cause mortality | | | | | | | | | |
| Deaths (n) |  | |  | | 308 | | 194 | | 516 | |  | | 39 | | 562 | | 80 | | 325 | |
| Events per 100.000 person days |  | |  | | 0.75 | | 0.45 | | 0.88 | |  | | 1.47 | | 0.59 | | 1.01 | | 0.99 | |
| Crude HR (95%CI) |  | |  | | 0.87 (0.75 - 1.02) | | 0.51 (0.43 - 0.60) | | Ref | |  | | 1.44 (1.02 - 2.04) | | 0.60 (0.52 - 0.68) | | 1.01 (0.79 - 1.30) | | Ref | |
| Age adjusted HR (95% CI) |  | |  | | 0.74 (0.64 - 0.86) | | 0.46 (0.39 - 0.54) | | Ref | |  | | 1.23 (0.87 - 1.75) | | 0.54 (0.47 - 0.62) | | 1.04 (0.81 - 1.32) | | Ref | |
| Age and gender adjusted HR (95% CI) |  | |  | | 0.77 (0.66 - 0.89) | | 0.45 (0.38 - 0.54) | | Ref | |  | | 1.32 (0.93 - 1.88) | | 0.54 (0.47 - 0.62) | | 0.98 (0.76 - 1.25) | | Ref | |
| Age, sex and ALTCA adjusted HR (95% CI) |  | |  | | 0.64 (0.55 - 0.75) | | 0.52 (0.44 - 0.61) | | Ref | |  | | 0.81 (0.57 - 1.14) | | 0.56 (0.48 - 0.64) | | 0.99 (0.77 - 1.26) | | Ref | |
| Age, sex, ALTCA and education adjusted HR (95% CI) |  | |  | | 0.67 (0.57 - 0.78) | | 0.54 (0.45 - 0.63) | | Ref | |  | | 0.85 (0.60 - 1.21) | | 0.56 (0.49 - 0.65) | | 0.97 (0.76 - 1.24) | | Ref | |
|  | May and June 2022 (low COVID-19 disease burden) | | | | | | | | | | February and March 2022 (high COVID-19 disease burden) | | | | | | | | | |
|  | Four vaccine doses | | Three vaccine doses | | Two vaccine doses | | One vaccine dose | | Unvaccinated | | Four vaccine doses | | Three vaccine doses | | Two vaccine doses | | One vaccine dose | | Unvaccinated | |
|  | All-cause mortality | | | | | | | | | | All-cause mortality | | | | | | | | | |
| Deaths (n) | 6 | | 486 | | 122 | | 18 | | 116 | | 3 | | 398 | | 178 | | 26 | | 163 | |
| Events per 100.000 person days |  | | 0.77 | | 1.09 | | 2.14 | | 1.02 | |  | | 0.55 | | 1.04 | | 2.06 | | 1.19 | |
| Crude HR (95%CI) |  | | 0.75 (0.62 - 0.92) | | 1.07 (0.83 - 1.38) | | 2.10 (1.28 - 3.45) | | Ref | |  | | 0.46 (0.38 - 0.55) | | 0.87 (0.71 - 1.08) | | 1.73 (1.14 - 2.62) | | Ref | |
| Age adjusted HR (95% CI) |  | | 0.69 (0.56 - 0.85) | | 1.11 (0.86 - 1.43) | | 2.20 (1.34 - 3.62) | | Ref | |  | | 0.43 (0.36 - 0.51) | | 0.91 (0.73 - 1.12) | | 1.80 (1.19 - 2.73) | | Ref | |
| Age and gender adjusted HR (95% CI) |  | | 0.69 (0.57 - 0.85) | | 1.08 (0.83 - 1.39) | | 2.15 (1.31 - 3.53) | | Ref | |  | | 0.43 (0.36 - 0.52) | | 0.89 (0.72 - 1.10) | | 1.78 (1.18 - 2.70) | | Ref | |
| Age, sex and ALTCA adjusted HR (95% CI) |  | | 0.71 (0.58 - 0.87) | | 1.05 (0.81 - 1.35) | | 1.95 (1.18 - 3.20) | | Ref | |  | | 0.45 (0.37 - 0.54) | | 0.87 (0.71 - 1.08) | | 1.47 (0.97 - 2.24) | | Ref | |
| Age, sex, ALTCA and education adjusted HR (95% CI) |  | | 0.75 (0.61 - 0.93) | | 1.06 (0.82 - 1.37) | | 1.92 (1.16 - 3.16) | | Ref | |  | | 0.46 (0.38 - 0.55) | | 0.87 (0.70 - 1.07) | | 1.46 (0.96 - 2.21) | | Ref | |
| Estimates with less than 10 deaths in either the vaccinated or reference group are not shown | | | | | | | | | | | | | | | | | | | | |
| **Table S36:** Hazard ratios (HR) with 95% confidence intervals (95% CI) for all-cause mortality according to number of SARS-CoV-2 vaccine doses during different time periods for 60-74 year olds only. | | | | | | | | | | | | | | | | | | | | |
|  | | June and July 2021 (low COVID-19 disease burden) | | | | | | | | | | October and November 2021 (high COVID-19 disease burden) | | | | | | | | |
|  | | Four vaccine doses | | Three vaccine doses | | Two vaccine doses | | One vaccine dose | | Unvaccinated | | Four vaccine doses | | Three vaccine doses | | Two vaccine doses | | One vaccine dose | | Unvaccinated |
|  | | All-cause mortality | | | | | | | | | | All-cause mortality | | | | | | | | |
| Deaths (n) | |  | |  | | 1082 | | 535 | | 1173 | |  | | 175 | | 1756 | | 172 | | 801 |
| Events per 100.000 person days | |  | |  | | 2.72 | | 2.22 | | 5.62 | |  | | 6.04 | | 2.73 | | 6.82 | | 5.56 |
| Crude HR (95%CI) | |  | |  | | 0.51 (0.47 - 0.56) | | 0.40 (0.36 - 0.44) | | Ref | |  | | 1.11 (0.93 - 1.32) | | 0.49 (0.45 - 0.53) | | 1.23 (1.04 - 1.45) | | Ref |
| Age adjusted HR (95% CI) | |  | |  | | 0.45 (0.41 - 0.49) | | 0.39 (0.36 - 0.44) | | Ref | |  | | 0.92 (0.77 - 1.09) | | 0.46 (0.43 - 0.50) | | 1.24 (1.05 - 1.46) | | Ref |
| Age and gender adjusted HR (95% CI) | |  | |  | | 0.45 (0.41 - 0.49) | | 0.39 (0.35 - 0.43) | | Ref | |  | | 0.88 (0.74 - 1.05) | | 0.46 (0.42 - 0.50) | | 1.20 (1.01 - 1.41) | | Ref |
| Age, sex and ALTCA adjusted HR (95% CI) | |  | |  | | 0.49 (0.45 - 0.54) | | 0.47 (0.42 - 0.52) | | Ref | |  | | 0.64 (0.54 - 0.76) | | 0.52 (0.48 - 0.57) | | 1.03 (0.87 - 1.22) | | Ref |
| Age, sex, ALTCA and education adjusted HR (95% CI) | |  | |  | | 0.49 (0.45 - 0.54) | | 0.47 (0.42 - 0.52) | | Ref | |  | | 0.67 (0.56 - 0.80) | | 0.53 (0.49 - 0.58) | | 1.03 (0.88 - 1.22) | | Ref |
|  | | May and June 2022 (low COVID-19 disease burden) | | | | | | | | | | February and March 2022 (high COVID-19 disease burden) | | | | | | | | |
|  | | Four vaccine doses | | Three vaccine doses | | Two vaccine doses | | One vaccine dose | | Unvaccinated | | Four vaccine doses | | Three vaccine doses | | Two vaccine doses | | One vaccine dose | | Unvaccinated |
|  | | All-cause mortality | | | | | | | | | | All-cause mortality | | | | | | | | |
| Deaths (n) | | 29 | | 1480 | | 302 | | 36 | | 313 | | 4 | | 1488 | | 443 | | 83 | | 432 |
| Events per 100.000 person days | | 7.23 | | 2.77 | | 6.04 | | 7.58 | | 4.07 | |  | | 2.6 | | 6.52 | | 11.85 | | 5.14 |
| Crude HR (95%CI) | | 1.79 (1.22 - 2.63) | | 0.68 (0.60 - 0.77) | | 1.48 (1.27 - 1.74) | | 1.86 (1.32 - 2.63) | | Ref | |  | | 0.50 (0.45 - 0.56) | | 1.26 (1.11 - 1.44) | | 2.30 (1.82 - 2.91) | | Ref |
| Age adjusted HR (95% CI) | | 1.55 (1.05 - 2.27) | | 0.66 (0.59 - 0.75) | | 1.52 (1.30 - 1.78) | | 1.84 (1.30 - 2.60) | | Ref | |  | | 0.49 (0.44 - 0.54) | | 1.29 (1.13 - 1.48) | | 2.25 (1.78 - 2.85) | | Ref |
| Age and gender adjusted HR (95% CI) | | 1.48 (1.01 - 2.18) | | 0.65 (0.58 - 0.74) | | 1.50 (1.28 - 1.76) | | 1.80 (1.27 - 2.54) | | Ref | |  | | 0.48 (0.43 - 0.53) | | 1.27 (1.11 - 1.45) | | 2.22 (1.76 - 2.81) | | Ref |
| Age, sex and ALTCA adjusted HR (95% CI) | | 1.15 (0.78 - 1.70) | | 0.66 (0.59 - 0.75) | | 1.22 (1.04 - 1.43) | | 1.50 (1.06 - 2.12) | | Ref | |  | | 0.50 (0.45 - 0.56) | | 1.04 (0.91 - 1.19) | | 1.72 (1.35 - 2.18) | | Ref |
| Age, sex, ALTCA and education adjusted HR (95% CI) | | 1.21 (0.82 - 1.79) | | 0.67 (0.59 - 0.76) | | 1.22 (1.04 - 1.44) | | 1.50 (1.06 - 2.12) | | Ref | |  | | 0.50 (0.45 - 0.56) | | 1.03 (0.91 - 1.18) | | 1.70 (1.34 - 2.15) | | Ref |
| Estimates with less than 10 deaths in either the vaccinated or reference group are not shown | | | | | | | | | | | | | | | | | | | | |

| **Table S37:** Hazard ratios (HR) with 95% confidence intervals (95% CI) for all-cause mortality according to number of SARS-CoV-2 vaccine doses during different time periods for 75-84 year olds only. | | | | | | | | | | | | | | | | | | | | |
| --- | --- | --- | --- | --- | --- | --- | --- | --- | --- | --- | --- | --- | --- | --- | --- | --- | --- | --- | --- | --- |
|  | June and July 2021 (low COVID-19 disease burden) | | | | | | | | | | October and November 2021 (high COVID-19 disease burden) | | | | | | | | | |
|  | Four vaccine doses | | Three vaccine doses | | Two vaccine doses | | One vaccine dose | | Unvaccinated | | Four vaccine doses | | Three vaccine doses | | Two vaccine doses | | One vaccine dose | | Unvaccinated | |
|  | All-cause mortality | | | | | | | | | | All-cause mortality | | | | | | | | | |
| Deaths (n) |  | |  | | 1981 | | 463 | | 1075 | |  | | 533 | | 2451 | | 189 | | 903 | |
| Events per 100.000 person days |  | |  | | 7.88 | | 10.44 | | 18.64 | |  | | 11.92 | | 9.47 | | 23.92 | | 21.77 | |
| Crude HR (95%CI) |  | |  | | 0.42 (0.39 - 0.45) | | 0.60 (0.54 - 0.67) | | Ref | |  | | 0.58 (0.51 - 0.65) | | 0.44 (0.41 - 0.47) | | 1.10 (0.94 - 1.29) | | Ref | |
| Age adjusted HR (95% CI) |  | |  | | 0.41 (0.38 - 0.44) | | 0.68 (0.61 - 0.76) | | Ref | |  | | 0.53 (0.47 - 0.60) | | 0.45 (0.41 - 0.48) | | 1.11 (0.95 - 1.30) | | Ref | |
| Age and gender adjusted HR (95% CI) |  | |  | | 0.40 (0.37 - 0.43) | | 0.67 (0.59 - 0.75) | | Ref | |  | | 0.51 (0.46 - 0.58) | | 0.43 (0.40 - 0.47) | | 1.09 (0.93 - 1.28) | | Ref | |
| Age, sex and ALTCA adjusted HR (95% CI) |  | |  | | 0.46 (0.43 - 0.50) | | 0.71 (0.64 - 0.80) | | Ref | |  | | 0.52 (0.46 - 0.58) | | 0.50 (0.46 - 0.54) | | 0.96 (0.82 - 1.12) | | Ref | |
| Age, sex, ALTCA and education adjusted HR (95% CI) |  | |  | | 0.46 (0.43 - 0.49) | | 0.71 (0.63 - 0.79) | | Ref | |  | | 0.53 (0.47 - 0.59) | | 0.50 (0.46 - 0.54) | | 0.96 (0.82 - 1.12) | | Ref | |
|  | May and June 2022 (low COVID-19 disease burden) | | | | | | | | | | February and March 2022 (high COVID-19 disease burden) | | | | | | | | | |
|  | Four vaccine doses | | Three vaccine doses | | Two vaccine doses | | One vaccine dose | | Unvaccinated | | Four vaccine doses | | Three vaccine doses | | Two vaccine doses | | One vaccine dose | | Unvaccinated | |
|  | All-cause mortality | | | | | | | | | | All-cause mortality | | | | | | | | | |
| Deaths (n) | 50 | | 2148 | | 315 | | 28 | | 340 | | 7 | | 2300 | | 520 | | 84 | | 481 | |
| Events per 100.000 person days | 8.61 | | 8.47 | | 18.66 | | 18.39 | | 14.40 | |  | | 8.52 | | 23.68 | | 32.94 | | 19.15 | |
| Crude HR (95%CI) | 0.61 (0.45 - 0.83) | | 0.59 (0.53 - 0.66) | | 1.29 (1.11 - 1.51) | | 1.28 (0.87 - 1.88) | | Ref | |  | | 0.44 (0.40 - 0.49) | | 1.24 (1.09 - 1.40) | | 1.73 (1.37 - 2.18) | | Ref | |
| Age adjusted HR (95% CI) | 0.55 (0.40 - 0.74) | | 0.58 (0.51 - 0.65) | | 1.27 (1.09 - 1.49) | | 1.28 (0.87 - 1.88) | | Ref | |  | | 0.44 (0.39 - 0.48) | | 1.23 (1.08 - 1.39) | | 1.69 (1.34 - 2.14) | | Ref | |
| Age and gender adjusted HR (95% CI) | 0.52 (0.39 - 0.71) | | 0.56 (0.50 - 0.63) | | 1.27 (1.09 - 1.48) | | 1.27 (0.87 - 1.87) | | Ref | |  | | 0.42 (0.38 - 0.47) | | 1.22 (1.08 - 1.38) | | 1.69 (1.34 - 2.14) | | Ref | |
| Age, sex and ALTCA adjusted HR (95% CI) | 0.52 (0.39 - 0.71) | | 0.56 (0.50 - 0.63) | | 1.02 (0.88 - 1.19) | | 1.07 (0.73 - 1.57) | | Ref | |  | | 0.43 (0.39 - 0.48) | | 1.01 (0.89 - 1.15) | | 1.37 (1.08 - 1.73) | | Ref | |
| Age, sex, ALTCA and education adjusted HR (95% CI) | 0.55 (0.40 - 0.74) | | 0.56 (0.50 - 0.63) | | 1.03 (0.88 - 1.20) | | 1.07 (0.73 - 1.58) | | Ref | |  | | 0.44 (0.40 - 0.48) | | 1.01 (0.89 - 1.14) | | 1.36 (1.08 - 1.72) | | Ref | |
| Estimates with less than 10 deaths in either the vaccinated or reference group are not shown | | | | | | | | | | | | | | | | | | | | |
| **Table S38:** Hazard ratios (HR) with 95% confidence intervals (95% CI) for all-cause mortality according to number of SARS-CoV-2 vaccine doses during different time periods for 85+ year olds only. | | | | | | | | | | | | | | | | | | | | |
|  | | June and July 2021 (low COVID-19 disease burden) | | | | | | | | | | October and November 2021 (high COVID-19 disease burden) | | | | | | | | |
|  | | Four vaccine doses | | Three vaccine doses | | Two vaccine doses | | One vaccine dose | | Unvaccinated | | Four vaccine doses | | Three vaccine doses | | Two vaccine doses | | One vaccine dose | | Unvaccinated |
|  | | All-cause mortality | | | | | | | | | | All-cause mortality | | | | | | | | |
| Deaths (n) | |  | | 1 | | 2769 | | 376 | | 1612 | |  | | 1039 | | 2577 | | 225 | | 1403 |
| Events per 100.000 person days | |  | |  | | 30.69 | | 49.82 | | 60.42 | |  | | 35.01 | | 36.85 | | 68.65 | | 67.26 |
| Crude HR (95%CI) | |  | |  | | 0.51 (0.48 - 0.54) | | 0.83 (0.74 - 0.93) | | Ref | |  | | 0.51 (0.47 - 0.56) | | 0.56 (0.53 - 0.60) | | 1.02 (0.89 - 1.18) | | Ref |
| Age adjusted HR (95% CI) | |  | |  | | 0.57 (0.54 - 0.61) | | 0.91 (0.81 - 1.02) | | Ref | |  | | 0.56 (0.51 - 0.61) | | 0.65 (0.61 - 0.70) | | 1.07 (0.93 - 1.23) | | Ref |
| Age and gender adjusted HR (95% CI) | |  | |  | | 0.56 (0.53 - 0.60) | | 0.89 (0.79 - 1.00) | | Ref | |  | | 0.55 (0.50 - 0.60) | | 0.64 (0.59 - 0.68) | | 1.06 (0.92 - 1.22) | | Ref |
| Age, sex and ALTCA adjusted HR (95% CI) | |  | |  | | 0.56 (0.53 - 0.60) | | 0.88 (0.78 - 0.98) | | Ref | |  | | 0.54 (0.50 - 0.59) | | 0.64 (0.60 - 0.68) | | 1.01 (0.88 - 1.16) | | Ref |
| Age, sex, ALTCA and education adjusted HR (95% CI) | |  | |  | | 0.56 (0.53 - 0.60) | | 0.88 (0.78 - 0.98) | | Ref | |  | | 0.54 (0.50 - 0.59) | | 0.64 (0.60 - 0.68) | | 1.00 (0.87 - 1.16) | | Ref |
|  | | May and June 2022 (low COVID-19 disease burden) | | | | | | | | | | February and March 2022 (high COVID-19 disease burden) | | | | | | | | |
|  | | Four vaccine doses | | Three vaccine doses | | Two vaccine doses | | One vaccine dose | | Unvaccinated | | Four vaccine doses | | Three vaccine doses | | Two vaccine doses | | One vaccine dose | | Unvaccinated |
|  | | All-cause mortality | | | | | | | | | | All-cause mortality | | | | | | | | |
| Deaths (n) | | 57 | | 2338 | | 444 | | 52 | | 515 | | 3 | | 2910 | | 662 | | 106 | | 792 |
| Events per 100.000 person days | | 21.82 | | 28.52 | | 55.22 | | 72.2 | | 49.50 | |  | | 33.25 | | 66.22 | | 71.68 | | 67.93 |
| Crude HR (95%CI) | | 0.42 (0.32 - 0.56) | | 0.58 (0.52 - 0.63) | | 1.12 (0.98 - 1.27) | | 1.46 (1.10 - 1.94) | | Ref | |  | | 0.49 (0.45 - 0.53) | | 0.98 (0.88 - 1.08) | | 1.05 (0.86 - 1.29) | | Ref |
| Age adjusted HR (95% CI) | | 0.47 (0.36 - 0.62) | | 0.67 (0.61 - 0.74) | | 1.15 (1.02 - 1.31) | | 1.49 (1.12 - 1.98) | | Ref | |  | | 0.58 (0.53 - 0.63) | | 1.02 (0.92 - 1.13) | | 1.06 (0.87 - 1.30) | | Ref |
| Age and gender adjusted HR (95% CI) | | 0.46 (0.34 - 0.60) | | 0.65 (0.59 - 0.72) | | 1.15 (1.01 - 1.30) | | 1.49 (1.12 - 1.98) | | Ref | |  | | 0.56 (0.52 - 0.61) | | 1.01 (0.91 - 1.12) | | 1.06 (0.87 - 1.30) | | Ref |
| Age, sex and ALTCA adjusted HR (95% CI) | | 0.45 (0.34 - 0.60) | | 0.61 (0.56 - 0.67) | | 1.05 (0.93 - 1.20) | | 1.39 (1.04 - 1.85) | | Ref | |  | | 0.54 (0.50 - 0.58) | | 0.93 (0.84 - 1.03) | | 0.98 (0.80 - 1.20) | | Ref |
| Age, sex, ALTCA and education adjusted HR (95% CI) | | 0.46 (0.34 - 0.61) | | 0.61 (0.56 - 0.67) | | 1.05 (0.92 - 1.19) | | 1.36 (1.02 - 1.81) | | Ref | |  | | 0.54 (0.50 - 0.58) | | 0.93 (0.84 - 1.03) | | 0.97 (0.80 - 1.19) | | Ref |
| Estimates with less than 10 deaths in either the vaccinated or reference group are not shown | | | | | | | | | | | | | | | | | | | | |

| **Table S39:** Hazard ratios (HR) with 95% confidence intervals (95% CI) for all-cause mortality according to number of SARS-CoV-2 vaccine doses during different time periods for individuals with compulsory schooling (9 years) or lower only. | | | | | | | | | | | | | | | | | | | | |
| --- | --- | --- | --- | --- | --- | --- | --- | --- | --- | --- | --- | --- | --- | --- | --- | --- | --- | --- | --- | --- |
|  | | June and July 2021 (low COVID-19 disease burden) | | | | | | | | | | October and November 2021 (high COVID-19 disease burden) | | | | | | | | |
|  | | Four vaccine doses | | Three vaccine doses | | Two vaccine doses | | One vaccine dose | | Unvaccinated | | Four vaccine doses | Three vaccine doses | | Two vaccine doses | | One vaccine dose | | Unvaccinated | |
|  | | All-cause mortality | | | | | | | | | | All-cause mortality | | | | | | | | |
| Deaths (n) | |  | | 1 | | 2420 | | 533 | | 1886 | |  | 835 | | 2639 | | 282 | | 1660 | |
| Events per 100.000 person days | |  | | 195.31 | | 8.9 | | 3.24 | | 4.47 | |  | 23.04 | | 5.39 | | 4.43 | | 6.57 | |
| Crude HR (95%CI) | |  | | 40.14 (5.64 - 285.45) | | 2.05 (1.93 - 2.18) | | 0.73 (0.66 - 0.80) | | Ref | |  | 3.53 (3.23 - 3.86) | | 0.82 (0.77 - 0.87) | | 0.67 (0.59 - 0.76) | | Ref | |
| Age adjusted HR (95% CI) | |  | |  | | 0.53 (0.50 - 0.57) | | 0.65 (0.59 - 0.71) | | Ref | |  | 0.66 (0.60 - 0.72) | | 0.51 (0.48 - 0.54) | | 1.06 (0.94 - 1.21) | | Ref | |
| Age and sex adjusted HR (95% CI) | |  | |  | | 0.53 (0.50 - 0.57) | | 0.65 (0.59 - 0.72) | | Ref | |  | 0.65 (0.60 - 0.72) | | 0.51 (0.48 - 0.54) | | 1.05 (0.93 - 1.19) | | Ref | |
| Age, sex and ALTCA adjusted HR (95% CI) | |  | |  | | 0.54 (0.51 - 0.58) | | 0.73 (0.66 - 0.80) | | Ref | |  | 0.59 (0.54 - 0.65) | | 0.54 (0.51 - 0.57) | | 1.00 (0.88 - 1.13) | | Ref | |
|  | | May and June 2022 (low COVID-19 disease burden) | | | | | | | | | | February and March 2022 (high COVID-19 disease burden) | | | | | | | | |
|  | | Four vaccine doses | | Three vaccine doses | | Two vaccine doses | | One vaccine dose | | Unvaccinated | | Four vaccine doses | Three vaccine doses | | Two vaccine doses | | One vaccine dose | | Unvaccinated | |
|  | | All-cause mortality | | | | | | | | | | All-cause mortality | | | | | | | | |
| Deaths (n) | | 48 | | 2342 | | 515 | | 67 | | 604 | | 5 | 2653 | | 780 | | 145 | | 877 | |
| Events per 100.000 person days | | 17.56 | | 6.03 | | 4.47 | | 5.78 | | 6.09 | |  | 6.62 | | 5.03 | | 8.49 | | 7.98 | |
| Crude HR (95%CI) | | 2.83 (2.11 - 3.81) | | 0.99 (0.91 - 1.08) | | 0.73 (0.65 - 0.83) | | 0.95 (0.74 - 1.22) | | Ref | |  | 0.83 (0.77 - 0.90) | | 0.63 (0.57 - 0.69) | | 1.07 (0.90 - 1.28) | | Ref | |
| Age adjusted HR (95% CI) | | 0.77 (0.57 - 1.04) | | 0.59 (0.54 - 0.65) | | 1.09 (0.97 - 1.22) | | 1.48 (1.15 - 1.90) | | Ref | |  | 0.49 (0.45 - 0.53) | | 0.99 (0.90 - 1.10) | | 1.37 (1.15 - 1.64) | | Ref | |
| Age and sex adjusted HR (95% CI) | | 0.76 (0.57 - 1.03) | | 0.59 (0.54 - 0.65) | | 1.08 (0.96 - 1.22) | | 1.47 (1.14 - 1.89) | | Ref | |  | 0.49 (0.45 - 0.53) | | 0.99 (0.90 - 1.09) | | 1.36 (1.14 - 1.62) | | Ref | |
| Age, sex and ALTCA adjusted HR (95% CI) | | 0.68 (0.50 - 0.92) | | 0.57 (0.52 - 0.62) | | 0.95 (0.85 - 1.07) | | 1.35 (1.04 - 1.73) | | Ref | |  | 0.47 (0.44 - 0.51) | | 0.89 (0.80 - 0.98) | | 1.21 (1.01 - 1.44) | | Ref | |
| Estimates with less than 10 deaths in either the vaccinated or reference group are not shown | | | | | | | | | | | | | | | | | | | | |
| **Table S40:** Hazard ratios (HR) with 95% confidence intervals (95% CI) for all-cause mortality according to number of SARS-CoV-2 vaccine doses during different time periods for individuals with non-secondary education only. | | | | | | | | | | | | | | | | | | | | |
|  | June and July 2021 (low COVID-19 disease burden) | | | | | | | | | | October and November 2021 (high COVID-19 disease burden) | | | | | | | | | |
|  | Four vaccine doses | | Three vaccine doses | | Two vaccine doses | | One vaccine dose | | Unvaccinated | | Four vaccine doses | | | Three vaccine doses | | Two vaccine doses | | One vaccine dose | | Unvaccinated |
|  | All-cause mortality | | | | | | | | | | All-cause mortality | | | | | | | | | |
| Deaths (n) |  | |  | | 2870 | | 826 | | 2065 | |  | | | 739 | | 3764 | | 321 | | 1464 |
| Events per 100.000 person days |  | |  | | 4.5 | | 1.68 | | 2.66 | |  | | | 11.94 | | 3.05 | | 3 | | 3.19 |
| Crude HR (95%CI) |  | |  | | 1.80 (1.70 - 1.91) | | 0.64 (0.59 - 0.69) | | Ref | |  | | | 3.73 (3.39 - 4.11) | | 0.96 (0.90 - 1.02) | | 0.93 (0.82 - 1.05) | | Ref |
| Age adjusted HR (95% CI) |  | |  | | 0.45 (0.42 - 0.47) | | 0.51 (0.47 - 0.55) | | Ref | |  | | | 0.54 (0.49 - 0.59) | | 0.52 (0.49 - 0.55) | | 1.09 (0.96 - 1.23) | | Ref |
| Age and sex adjusted HR (95% CI) |  | |  | | 0.44 (0.41 - 0.46) | | 0.50 (0.46 - 0.54) | | Ref | |  | | | 0.52 (0.47 - 0.57) | | 0.50 (0.47 - 0.53) | | 1.06 (0.94 - 1.19) | | Ref |
| Age, sex and ALTCA adjusted HR (95% CI) |  | |  | | 0.50 (0.47 - 0.53) | | 0.61 (0.56 - 0.66) | | Ref | |  | | | 0.49 (0.45 - 0.54) | | 0.60 (0.56 - 0.63) | | 0.96 (0.85 - 1.09) | | Ref |
|  | May and June 2022 (low COVID-19 disease burden) | | | | | | | | | | February and March 2022 (high COVID-19 disease burden) | | | | | | | | | |
|  | Four vaccine doses | | Three vaccine doses | | Two vaccine doses | | One vaccine dose | | Unvaccinated | | Four vaccine doses | | | Three vaccine doses | | Two vaccine doses | | One vaccine dose | | Unvaccinated |
|  | All-cause mortality | | | | | | | | | | All-cause mortality | | | | | | | | | |
| Deaths (n) | 64 | | 3212 | | 560 | | 56 | | 535 | | 11 | | | 3522 | | 891 | | 125 | | 803 |
| Events per 100.000 person days | 10.48 | | 3.51 | | 3.62 | | 5.49 | | 3.65 | | 14.45 | | | 3.5 | | 3.78 | | 7.28 | | 4.46 |
| Crude HR (95%CI) | 2.80 (2.16 - 3.64) | | 0.96 (0.88 - 1.05) | | 0.99 (0.88 - 1.12) | | 1.50 (1.14 - 1.98) | | Ref | | 3.18 (1.75 - 5.76) | | | 0.78 (0.72 - 0.84) | | 0.85 (0.77 - 0.93) | | 1.64 (1.36 - 1.99) | | Ref |
| Age adjusted HR (95% CI) | 0.62 (0.47 - 0.81) | | 0.62 (0.57 - 0.68) | | 1.33 (1.18 - 1.50) | | 1.71 (1.30 - 2.25) | | Ref | | 1.21 (0.67 - 2.20) | | | 0.47 (0.43 - 0.51) | | 1.18 (1.07 - 1.29) | | 1.57 (1.30 - 1.90) | | Ref |
| Age and sex adjusted HR (95% CI) | 0.59 (0.45 - 0.77) | | 0.60 (0.55 - 0.66) | | 1.30 (1.15 - 1.46) | | 1.68 (1.27 - 2.21) | | Ref | | 1.14 (0.63 - 2.08) | | | 0.45 (0.42 - 0.49) | | 1.14 (1.04 - 1.26) | | 1.55 (1.28 - 1.88) | | Ref |
| Age, sex and ALTCA adjusted HR (95% CI) | 0.56 (0.43 - 0.73) | | 0.63 (0.57 - 0.69) | | 1.12 (0.99 - 1.26) | | 1.44 (1.09 - 1.89) | | Ref | | 0.92 (0.51 - 1.67) | | | 0.48 (0.45 - 0.52) | | 1.00 (0.91 - 1.10) | | 1.27 (1.05 - 1.53) | | Ref |

| **Table S41:** Hazard ratios (HR) with 95% confidence intervals (95% CI) for all-cause mortality according to number of SARS-CoV-2 vaccine doses during different time periods for individuals with secondary education only. | | | | | | | | | | |
| --- | --- | --- | --- | --- | --- | --- | --- | --- | --- | --- |
|  | June and July 2021 (low COVID-19 disease burden) | | | | | October and November 2021 (high COVID-19 disease burden) | | | | |
|  | Four vaccine doses | Three vaccine doses | Two vaccine doses | One vaccine dose | Unvaccinated | Four vaccine doses | Three vaccine doses | Two vaccine doses | One vaccine dose | Unvaccinated |
|  | All-cause mortality | | | | | All-cause mortality | | | | |
| Deaths (n) |  |  | 464 | 121 | 335 |  | 114 | 514 | 44 | 208 |
| Events per 100.000 person days |  |  | 2.57 | 0.69 | 1.24 |  | 7.76 | 1.2 | 1.14 | 1.54 |
| Crude HR (95%CI) |  |  | 2.30 (1.98 - 2.67) | 0.56 (0.46 - 0.70) | Ref |  | 5.71 (4.44 - 7.33) | 0.78 (0.67 - 0.92) | 0.74 (0.54 - 1.03) | Ref |
| Age adjusted HR (95% CI) |  |  | 0.45 (0.39 - 0.52) | 0.46 (0.37 - 0.57) | Ref |  | 0.55 (0.43 - 0.71) | 0.51 (0.44 - 0.60) | 1.24 (0.89 - 1.71) | Ref |
| Age and sex adjusted HR (95% CI) |  |  | 0.43 (0.37 - 0.50) | 0.44 (0.36 - 0.55) | Ref |  | 0.53 (0.41 - 0.68) | 0.49 (0.42 - 0.58) | 1.21 (0.88 - 1.68) | Ref |
| Age, sex and ALTCA adjusted HR (95% CI) |  |  | 0.49 (0.42 - 0.57) | 0.55 (0.45 - 0.68) | Ref |  | 0.50 (0.39 - 0.65) | 0.55 (0.46 - 0.64) | 1.03 (0.74 - 1.42) | Ref |
|  | May and June 2022 (low COVID-19 disease burden) | | | | | February and March 2022 (high COVID-19 disease burden) | | | | |
|  | Four vaccine doses | Three vaccine doses | Two vaccine doses | One vaccine dose | Unvaccinated | Four vaccine doses | Three vaccine doses | Two vaccine doses | One vaccine dose | Unvaccinated |
|  | All-cause mortality | | | | | All-cause mortality | | | | |
| Deaths (n) | 14 | 537 | 80 | 8 | 72 |  | 517 | 102 | 20 | 114 |
| Events per 100.000 person days | 6.98 | 1.93 | 1.72 |  | 1.50 |  | 1.59 | 1.3 | 3.26 | 2.02 |
| Crude HR (95%CI) | 4.75 (2.66 - 8.49) | 1.28 (1.00 - 1.64) | 1.15 (0.83 - 1.58) |  | Ref |  | 0.79 (0.64 - 0.97) | 0.64 (0.49 - 0.83) | 1.61 (1.00 - 2.59) | Ref |
| Age adjusted HR (95% CI) | 0.64 (0.36 - 1.17) | 0.92 (0.72 - 1.18) | 1.92 (1.40 - 2.64) |  | Ref |  | 0.54 (0.44 - 0.66) | 1.23 (0.94 - 1.60) | 1.99 (1.23 - 3.20) | Ref |
| Age and sex adjusted HR (95% CI) | 0.56 (0.31 - 1.03) | 0.88 (0.68 - 1.12) | 1.90 (1.38 - 2.61) |  | Ref |  | 0.52 (0.42 - 0.64) | 1.21 (0.93 - 1.59) | 1.98 (1.23 - 3.19) | Ref |
| Age, sex and ALTCA adjusted HR (95% CI) | 0.51 (0.28 - 0.94) | 0.84 (0.66 - 1.08) | 1.52 (1.10 - 2.10) |  | Ref |  | 0.51 (0.42 - 0.63) | 0.95 (0.72 - 1.24) | 1.52 (0.94 - 2.46) | Ref |
| Estimates with less than 10 deaths in either the vaccinated or reference group are not shown | | | | | | | | | | |

| **Table S42:** Hazard ratios (HR) with 95% confidence intervals (95% CI) for all-cause mortality according to number of SARS-CoV-2 vaccine doses during different time periods for individuals withnon-tertiary education only. | | | | | | | | | | |
| --- | --- | --- | --- | --- | --- | --- | --- | --- | --- | --- |
|  | June and July 2021 (low COVID-19 disease burden) | | | | | October and November 2021 (high COVID-19 disease burden) | | | | |
|  | Four vaccine doses | Three vaccine doses | Two vaccine doses | One vaccine dose | Unvaccinated | Four vaccine doses | Three vaccine doses | Two vaccine doses | One vaccine dose | Unvaccinated |
|  | All-cause mortality | | | | | All-cause mortality | | | | |
| Deaths (n) |  |  | 81 | 31 | 42 |  | 20 | 114 | 3 | 29 |
| Events per 100.000 person days |  |  | 1.67 | 1.13 | 1.44 |  | 4.58 | 1.48 |  | 1.59 |
| Crude HR (95%CI) |  |  | 1.38 (0.94 - 2.03) | 0.80 (0.50 - 1.29) | Ref |  | 4.06 (2.10 - 7.86) | 0.93 (0.62 - 1.40) |  | Ref |
| Age adjusted HR (95% CI) |  |  | 0.51 (0.35 - 0.74) | 0.65 (0.40 - 1.06) | Ref |  | 0.87 (0.45 - 1.71) | 0.66 (0.44 - 0.99) |  | Ref |
| Age and sex adjusted HR (95% CI) |  |  | 0.50 (0.34 - 0.73) | 0.65 (0.40 - 1.06) | Ref |  | 0.87 (0.44 - 1.69) | 0.65 (0.43 - 0.97) |  | Ref |
| Age, sex and ALTCA adjusted HR (95% CI) |  |  | 0.59 (0.40 - 0.86) | 0.87 (0.54 - 1.42) | Ref |  | 0.78 (0.40 - 1.50) | 0.75 (0.50 - 1.13) |  | Ref |
|  | May and June 2022 (low COVID-19 disease burden) | | | | | February and March 2022 (high COVID-19 disease burden) | | | | |
|  | Four vaccine doses | Three vaccine doses | Two vaccine doses | One vaccine dose | Unvaccinated | Four vaccine doses | Three vaccine doses | Two vaccine doses | One vaccine dose | Unvaccinated |
|  | All-cause mortality | | | | | All-cause mortality | | | | |
| Deaths (n) | 2 | 75 | 11 |  | 18 |  | 88 | 29 | 3 | 29 |
| Events per 100.000 person days |  | 1.44 | 2.51 |  | 2.22 |  | 1.4 | 3.95 |  | 3.42 |
| Crude HR (95%CI) |  | 0.65 (0.39 - 1.09) | 1.12 (0.53 - 2.37) |  | Ref |  | 0.41 (0.27 - 0.62) | 1.15 (0.69 - 1.92) |  | Ref |
| Age adjusted HR (95% CI) |  | 0.46 (0.28 - 0.78) | 1.26 (0.59 - 2.66) |  | Ref |  | 0.30 (0.20 - 0.46) | 1.47 (0.88 - 2.45) |  | Ref |
| Age and sex adjusted HR (95% CI) |  | 0.46 (0.28 - 0.77) | 1.26 (0.60 - 2.67) |  | Ref |  | 0.30 (0.20 - 0.45) | 1.47 (0.88 - 2.47) |  | Ref |
| Age, sex and ALTCA adjusted HR (95% CI) |  | 0.45 (0.27 - 0.75) | 0.89 (0.42 - 1.90) |  | Ref |  | 0.31 (0.20 - 0.47) | 1.12 (0.67 - 1.87) |  | Ref |
| Estimates with less than 10 deaths in either the vaccinated or reference group are not shown | | | | | | | | | | |

| **Table S43:** Hazard ratios (HR) with 95% confidence intervals (95% CI) for all-cause mortality according to number of SARS-CoV-2 vaccine doses during different time periods for individuals with tertiary education only. | | | | | | | | | | |
| --- | --- | --- | --- | --- | --- | --- | --- | --- | --- | --- |
|  | June and July 2021 (low COVID-19 disease burden) | | | | | October and November 2021 (high COVID-19 disease burden) | | | | |
|  | Four vaccine doses | Three vaccine doses | Two vaccine doses | One vaccine dose | Unvaccinated | Four vaccine doses | Three vaccine doses | Two vaccine doses | One vaccine dose | Unvaccinated |
|  | All-cause mortality | | | | | All-cause mortality | | | | |
| Deaths (n) |  |  | 331 | 85 | 181 |  | 80 | 388 | 33 | 135 |
| Events per 100.000 person days |  |  | 1.45 | 0.47 | 0.91 |  | 3.35 | 0.85 | 1.29 | 1.41 |
| Crude HR (95%CI) |  |  | 1.73 (1.43 - 2.09) | 0.51 (0.40 - 0.66) | Ref |  | 2.38 (1.76 - 3.21) | 0.60 (0.50 - 0.73) | 0.91 (0.62 - 1.33) | Ref |
| Age adjusted HR (95% CI) |  |  | 0.48 (0.40 - 0.58) | 0.59 (0.45 - 0.78) | Ref |  | 0.47 (0.35 - 0.63) | 0.58 (0.48 - 0.71) | 1.59 (1.08 - 2.32) | Ref |
| Age and sex adjusted HR (95% CI) |  |  | 0.46 (0.38 - 0.55) | 0.58 (0.44 - 0.76) | Ref |  | 0.44 (0.32 - 0.59) | 0.56 (0.46 - 0.68) | 1.57 (1.07 - 2.29) | Ref |
| Age, sex and ALTCA adjusted HR (95% CI) |  |  | 0.51 (0.42 - 0.62) | 0.64 (0.49 - 0.84) | Ref |  | 0.42 (0.31 - 0.57) | 0.59 (0.49 - 0.72) | 1.16 (0.79 - 1.71) | Ref |
|  | May and June 2022 (low COVID-19 disease burden) | | | | | February and March 2022 (high COVID-19 disease burden) | | | | |
|  | Four vaccine doses | Three vaccine doses | Two vaccine doses | One vaccine dose | Unvaccinated | Four vaccine doses | Three vaccine doses | Two vaccine doses | One vaccine dose | Unvaccinated |
|  | All-cause mortality | | | | | All-cause mortality | | | | |
| Deaths (n) | 15 | 348 | 55 | 6 | 67 | 2 | 371 | 68 | 10 | 85 |
| Events per 100.000 person days | 4.53 | 1.21 | 1.82 |  | 1.61 |  | 1.04 | 1.28 | 1.89 | 1.83 |
| Crude HR (95%CI) | 2.96 (1.68 - 5.20) | 0.75 (0.58 - 0.98) | 1.13 (0.79 - 1.62) |  | Ref |  | 0.57 (0.45 - 0.72) | 0.70 (0.51 - 0.97) | 1.04 (0.54 - 2.00) | Ref |
| Age adjusted HR (95% CI) | 0.59 (0.33 - 1.05) | 0.66 (0.51 - 0.86) | 1.67 (1.17 - 2.38) |  | Ref |  | 0.52 (0.41 - 0.66) | 1.33 (0.96 - 1.82) | 1.31 (0.68 - 2.52) | Ref |
| Age and sex adjusted HR (95% CI) | 0.56 (0.32 - 1.01) | 0.64 (0.49 - 0.83) | 1.65 (1.16 - 2.36) |  | Ref |  | 0.50 (0.39 - 0.63) | 1.32 (0.96 - 1.82) | 1.31 (0.68 - 2.52) | Ref |
| Age, sex and ALTCA adjusted HR (95% CI) | 0.51 (0.28 - 0.92) | 0.57 (0.44 - 0.74) | 1.16 (0.81 - 1.67) |  | Ref |  | 0.46 (0.36 - 0.59) | 0.93 (0.67 - 1.28) | 0.93 (0.48 - 1.81) | Ref |
| Estimates with less than 10 deaths in either the vaccinated or reference group are not shown | | | | | | | | | | |

| **Table S44:** Hazard ratios (HR) with 95% confidence intervals (95% CI) for all-cause mortality according to number of SARS-CoV-2 vaccine doses during different time periods for ALTCA recipients only. | | | | | | | | | | |
| --- | --- | --- | --- | --- | --- | --- | --- | --- | --- | --- |
|  | June and July 2021 (low COVID-19 disease burden) | | | | | October and November 2021 (high COVID-19 disease burden) | | | | |
|  | Four vaccine doses | Three vaccine doses | Two vaccine doses | One vaccine dose | Unvaccinated | Four vaccine doses | Three vaccine doses | Two vaccine doses | One vaccine dose | Unvaccinated |
|  | All-cause mortality | | | | | All-cause mortality | | | | |
| Deaths (n) |  | 1 | 4591 | 956 | 2935 |  | 1533 | 4914 | 472 | 2312 |
| Events per 100.000 person days |  |  | 27.6 | 28.79 | 43.59 |  | 39.32 | 30.87 | 49.11 | 51.18 |
| Crude HR (95%CI) |  |  | 0.63 (0.61 - 0.66) | 0.67 (0.62 - 0.72) | Ref |  | 0.76 (0.71 - 0.81) | 0.61 (0.58 - 0.64) | 0.96 (0.87 - 1.06) | Ref |
| Age adjusted HR (95% CI) |  |  | 0.54 (0.52 - 0.57) | 0.76 (0.70 - 0.82) | Ref |  | 0.59 (0.55 - 0.64) | 0.62 (0.59 - 0.65) | 1.04 (0.94 - 1.15) | Ref |
| Age and sex adjusted HR (95% CI) |  |  | 0.53 (0.51 - 0.56) | 0.74 (0.69 - 0.80) | Ref |  | 0.58 (0.54 - 0.62) | 0.60 (0.57 - 0.63) | 1.01 (0.92 - 1.12) | Ref |
| Age, sex and education adjusted HR (95% CI) |  |  | 0.53 (0.51 - 0.56) | 0.74 (0.69 - 0.80) | Ref |  | 0.58 (0.54 - 0.62) | 0.60 (0.57 - 0.63) | 1.01 (0.91 - 1.12) | Ref |
|  | May and June 2022 (low COVID-19 disease burden) | | | | | February and March 2022 (high COVID-19 disease burden) | | | | |
|  | Four vaccine doses | Three vaccine doses | Two vaccine doses | One vaccine dose | Unvaccinated | Four vaccine doses | Three vaccine doses | Two vaccine doses | One vaccine dose | Unvaccinated |
|  | All-cause mortality | | | | | All-cause mortality | | | | |
| Deaths (n) | 109 | 4341 | 841 | 93 | 799 | 12 | 4978 | 1332 | 221 | 1233 |
| Events per 100.000 person days | 28.58 | 25.45 | 39.65 | 48.86 | 37.51 | 28.52 | 26.41 | 46.12 | 57.78 | 48.30 |
| Crude HR (95%CI) | 0.76 (0.62 - 0.93) | 0.68 (0.63 - 0.73) | 1.06 (0.96 - 1.17) | 1.30 (1.05 - 1.61) | Ref | 0.59 (0.33 - 1.04) | 0.55 (0.51 - 0.58) | 0.95 (0.88 - 1.03) | 1.20 (1.04 - 1.38) | Ref |
| Age adjusted HR (95% CI) | 0.64 (0.52 - 0.79) | 0.65 (0.60 - 0.70) | 1.10 (1.00 - 1.21) | 1.38 (1.12 - 1.72) | Ref | 0.65 (0.37 - 1.14) | 0.52 (0.49 - 0.55) | 1.02 (0.94 - 1.10) | 1.21 (1.05 - 1.39) | Ref |
| Age and sex adjusted HR (95% CI) | 0.61 (0.49 - 0.74) | 0.62 (0.58 - 0.67) | 1.07 (0.97 - 1.18) | 1.35 (1.09 - 1.68) | Ref | 0.60 (0.34 - 1.05) | 0.50 (0.47 - 0.53) | 0.99 (0.92 - 1.07) | 1.19 (1.03 - 1.37) | Ref |
| Age, sex and education adjusted HR (95% CI) | 0.61 (0.49 - 0.75) | 0.62 (0.58 - 0.67) | 1.07 (0.97 - 1.18) | 1.35 (1.09 - 1.67) | Ref | 0.59 (0.33 - 1.04) | 0.50 (0.47 - 0.53) | 0.99 (0.92 - 1.07) | 1.19 (1.03 - 1.37) | Ref |
| Estimates with less than 10 deaths in either the vaccinated or reference group are not shown | | | | | | | | | | |

| **Table S45:** Hazard ratios (HR) with 95% confidence intervals (95% CI) for all-cause mortality according to number of SARS-CoV-2 vaccine doses during different time periods for non-ALTCA recipients only. | | | | | | | | | | |
| --- | --- | --- | --- | --- | --- | --- | --- | --- | --- | --- |
|  | June and July 2021 (low COVID-19 disease burden) | | | | | October and November 2021 (high COVID-19 disease burden) | | | | |
|  | Four vaccine doses | Three vaccine doses | Two vaccine doses | One vaccine dose | Unvaccinated | Four vaccine doses | Three vaccine doses | Two vaccine doses | One vaccine dose | Unvaccinated |
|  | All-cause mortality | | | | | All-cause mortality | | | | |
| Deaths (n) |  |  | 1576 | 641 | 1595 |  | 255 | 2510 | 214 | 1197 |
| Events per 100.000 person days |  |  | 1.31 | 0.64 | 0.97 |  | 2.5 | 0.99 | 0.94 | 1.30 |
| Crude HR (95%CI) |  |  | 1.39 (1.30 - 1.50) | 0.65 (0.60 - 0.72) | Ref |  | 1.86 (1.61 - 2.15) | 0.76 (0.71 - 0.82) | 0.72 (0.62 - 0.83) | Ref |
| Age adjusted HR (95% CI) |  |  | 0.41 (0.38 - 0.45) | 0.47 (0.43 - 0.52) | Ref |  | 0.35 (0.30 - 0.41) | 0.45 (0.42 - 0.48) | 0.99 (0.85 - 1.14) | Ref |
| Age and sex adjusted HR (95% CI) |  |  | 0.40 (0.37 - 0.44) | 0.46 (0.42 - 0.51) | Ref |  | 0.33 (0.28 - 0.38) | 0.44 (0.41 - 0.47) | 0.95 (0.82 - 1.10) | Ref |
| Age, sex and education adjusted HR (95% CI) |  |  | 0.41 (0.38 - 0.45) | 0.48 (0.43 - 0.52) | Ref |  | 0.34 (0.30 - 0.40) | 0.45 (0.42 - 0.48) | 0.93 (0.81 - 1.08) | Ref |
|  | May and June 2022 (low COVID-19 disease burden) | | | | | February and March 2022 (high COVID-19 disease burden) | | | | |
|  | Four vaccine doses | Three vaccine doses | Two vaccine doses | One vaccine dose | Unvaccinated | Four vaccine doses | Three vaccine doses | Two vaccine doses | One vaccine dose | Unvaccinated |
|  | All-cause mortality | | | | | All-cause mortality | | | | |
| Deaths (n) | 34 | 2174 | 381 | 46 | 512 | 6 | 2173 | 538 | 82 | 679 |
| Events per 100.000 person days | 3.14 | 1.24 | 1.15 | 1.61 | 1.54 |  | 1.11 | 1.07 | 1.92 | 1.78 |
| Crude HR (95%CI) | 1.99 (1.40 - 2.82) | 0.80 (0.73 - 0.88) | 0.75 (0.66 - 0.85) | 1.04 (0.77 - 1.41) | Ref |  | 0.62 (0.57 - 0.67) | 0.60 (0.54 - 0.68) | 1.08 (0.86 - 1.36) | Ref |
| Age adjusted HR (95% CI) | 0.47 (0.33 - 0.67) | 0.55 (0.50 - 0.61) | 1.11 (0.97 - 1.27) | 1.48 (1.09 - 2.00) | Ref |  | 0.41 (0.38 - 0.45) | 0.93 (0.83 - 1.04) | 1.45 (1.15 - 1.82) | Ref |
| Age and gender adjusted HR (95% CI) | 0.44 (0.31 - 0.63) | 0.54 (0.49 - 0.59) | 1.09 (0.95 - 1.24) | 1.46 (1.08 - 1.98) | Ref |  | 0.40 (0.36 - 0.43) | 0.91 (0.81 - 1.02) | 1.44 (1.14 - 1.81) | Ref |
| Age, gender and education adjusted HR (95% CI) | 0.48 (0.34 - 0.69) | 0.55 (0.50 - 0.61) | 1.09 (0.95 - 1.24) | 1.40 (1.03 - 1.91) | Ref |  | 0.41 (0.37 - 0.44) | 0.89 (0.79 - 1.00) | 1.41 (1.12 - 1.77) | Ref |
| Estimates with less than 10 deaths in either the vaccinated or reference group are not shown | | | | | | | | | | |

| **Table S46:** Hazard ratios (HR) with 95% confidence intervals (95% CI) for all-cause mortality according to number of SARS-CoV-2 vaccine doses during different time periods for mRNA vaccines (last adiministered dose) only. | | | | | | | | | | |
| --- | --- | --- | --- | --- | --- | --- | --- | --- | --- | --- |
|  | June and July 2021 (low COVID-19 disease burden) | | | | | October and November 2021 (high COVID-19 disease burden) | | | | |
|  | Four vaccine doses | Three vaccine doses | Two vaccine doses | One vaccine dose | Unvaccinated | Four vaccine doses | Three vaccine doses | Two vaccine doses | One vaccine dose | Unvaccinated |
|  | All-cause mortality | | | | | All-cause mortality | | | | |
| Deaths (n) |  | 1 | 5938 | 1045 | 4530 |  | 1782 | 6575 | 337 | 3509 |
| Events per 100.000 person days |  |  | 5.18 | 1.37 | 2.66 |  | 12.67 | 2.91 | 4.22 | 3.64 |
| Crude HR (95%CI) |  |  | 2.05 (1.97 - 2.13) | 0.52 (0.48 - 0.55) | Ref |  | 3.56 (3.35 - 3.79) | 0.80 (0.77 - 0.83) | 1.14 (1.02 - 1.27) | Ref |
| Age adjusted HR (95% CI) |  |  | 0.49 (0.48 - 0.51) | 0.62 (0.58 - 0.67) | Ref |  | 0.58 (0.54 - 0.61) | 0.53 (0.51 - 0.55) | 1.30 (1.16 - 1.45) | Ref |
| Age and gender adjusted HR (95% CI) |  |  | 0.48 (0.46 - 0.50) | 0.61 (0.57 - 0.65) | Ref |  | 0.56 (0.53 - 0.60) | 0.51 (0.49 - 0.53) | 1.29 (1.15 - 1.44) | Ref |
| Age, sex and ALTCA adjusted HR (95% CI) |  |  | 0.53 (0.51 - 0.55) | 0.70 (0.66 - 0.75) | Ref |  | 0.53 (0.50 - 0.56) | 0.57 (0.55 - 0.60) | 1.17 (1.05 - 1.31) | Ref |
| Age, sex, ALTCA and education adjusted HR (95% CI) |  |  | 0.53 (0.51 - 0.55) | 0.71 (0.66 - 0.76) | Ref |  | 0.54 (0.50 - 0.57) | 0.58 (0.55 - 0.60) | 1.17 (1.04 - 1.31) | Ref |
|  | May and June 2022 (low COVID-19 disease burden) | | | | | February and March 2022 (high COVID-19 disease burden) | | | | |
|  | Four vaccine doses | Three vaccine doses | Two vaccine doses | One vaccine dose | Unvaccinated | Four vaccine doses | Three vaccine doses | Two vaccine doses | One vaccine dose | Unvaccinated |
|  | All-cause mortality | | | | | All-cause mortality | | | | |
| Deaths (n) | 143 | 6509 | 1190 | 111 | 1311 | 18 | 7146 | 1776 | 244 | 1912 |
| Events per 100.000 person days | 9.84 | 3.39 | 3.47 | 4.56 | 3.72 | 9.38 | 3.32 | 3.42 | 6.45 | 4.71 |
| Crude HR (95%CI) | 2.62 (2.20 - 3.12) | 0.91 (0.86 - 0.97) | 0.93 (0.86 - 1.01) | 1.23 (1.01 - 1.49) | Ref | 1.97 (1.24 - 3.13) | 0.71 (0.67 - 0.74) | 0.73 (0.68 - 0.78) | 1.38 (1.21 - 1.57) | Ref |
| Age adjusted HR (95% CI) | 0.62 (0.52 - 0.74) | 0.62 (0.58 - 0.66) | 1.27 (1.18 - 1.38) | 1.54 (1.27 - 1.87) | Ref | 0.87 (0.55 - 1.39) | 0.48 (0.46 - 0.51) | 1.10 (1.03 - 1.17) | 1.39 (1.22 - 1.59) | Ref |
| Age and gender adjusted HR (95% CI) | 0.59 (0.49 - 0.71) | 0.60 (0.57 - 0.64) | 1.26 (1.17 - 1.36) | 1.53 (1.26 - 1.86) | Ref | 0.81 (0.51 - 1.28) | 0.47 (0.44 - 0.49) | 1.09 (1.02 - 1.16) | 1.39 (1.22 - 1.59) | Ref |
| Age, sex and ALTCA adjusted HR (95% CI) | 0.57 (0.48 - 0.69) | 0.60 (0.57 - 0.64) | 1.06 (0.98 - 1.15) | 1.34 (1.11 - 1.63) | Ref | 0.71 (0.45 - 1.13) | 0.47 (0.45 - 0.50) | 0.93 (0.88 - 1.00) | 1.16 (1.02 - 1.33) | Ref |
| Age, sex, ALTCA and education adjusted HR (95% CI) | 0.60 (0.50 - 0.71) | 0.61 (0.57 - 0.64) | 1.07 (0.98 - 1.15) | 1.32 (1.09 - 1.61) | Ref | 0.74 (0.46 - 1.18) | 0.48 (0.45 - 0.50) | 0.93 (0.87 - 0.99) | 1.16 (1.02 - 1.33) | Ref |
| Estimates with less than 10 deaths in either the vaccinated or reference group are not shown | | | | | | | | | | |

| **Table S47:** Hazard ratios (HR) with 95% confidence intervals (95% CI) for all-cause mortality according to number of SARS-CoV-2 vaccine doses during different time periods for non-mRNA vaccines (last adiministered dose) only. | | | | | | | | | | |
| --- | --- | --- | --- | --- | --- | --- | --- | --- | --- | --- |
|  | June and July 2021 (low COVID-19 disease burden) | | | | | October and November 2021 (high COVID-19 disease burden) | | | | |
|  | Four vaccine doses | Three vaccine doses | Two vaccine doses | One vaccine dose | Unvaccinated | Four vaccine doses | Three vaccine doses | Two vaccine doses | One vaccine dose | Unvaccinated |
|  | All-cause mortality | | | | | All-cause mortality | | | | |
| Deaths (n) |  |  | 229 | 552 | 4530 |  | 6 | 849 | 349 | 3509 |
| Events per 100.000 person days |  |  | 1.04 | 1.98 | 2.66 |  |  | 2.02 | 2.21 | 3.64 |
| Crude HR (95%CI) |  |  | 0.37 (0.32 - 0.42) | 0.75 (0.69 - 0.82) | Ref |  |  | 0.55 (0.51 - 0.60) | 0.60 (0.54 - 0.67) | Ref |
| Age adjusted HR (95% CI) |  |  | 0.27 (0.24 - 0.31) | 0.44 (0.40 - 0.48) | Ref |  |  | 0.41 (0.38 - 0.44) | 0.98 (0.87 - 1.09) | Ref |
| Age and gender adjusted HR (95% CI) |  |  | 0.26 (0.23 - 0.30) | 0.43 (0.39 - 0.47) | Ref |  |  | 0.40 (0.37 - 0.43) | 0.94 (0.84 - 1.05) | Ref |
| Age, sex and ALTCA adjusted HR (95% CI) |  |  | 0.34 (0.30 - 0.39) | 0.53 (0.48 - 0.58) | Ref |  |  | 0.50 (0.46 - 0.54) | 0.87 (0.78 - 0.97) | Ref |
| Age, sex, ALTCA and education adjusted HR (95% CI) |  |  | 0.34 (0.30 - 0.39) | 0.53 (0.49 - 0.58) | Ref |  |  | 0.51 (0.47 - 0.55) | 0.87 (0.78 - 0.97) | Ref |
|  | May and June 2022 (low COVID-19 disease burden) | | | | | February and March 2022 (high COVID-19 disease burden) | | | | |
|  | Four vaccine doses | Three vaccine doses | Two vaccine doses | One vaccine dose | Unvaccinated | Four vaccine doses | Three vaccine doses | Two vaccine doses | One vaccine dose | Unvaccinated |
|  | All-cause mortality | | | | | All-cause mortality | | | | |
| Deaths (n) |  | 6 | 32 | 28 | 1311 |  | 5 | 94 | 59 | 1912 |
| Events per 100.000 person days |  |  | 3.89 | 4.53 | 3.72 |  |  | 7.91 | 6.74 | 4.71 |
| Crude HR (95%CI) |  |  | 1.05 (0.74 - 1.49) | 1.22 (0.84 - 1.77) | Ref |  |  | 1.69 (1.37 - 2.08) | 1.44 (1.11 - 1.86) | Ref |
| Age adjusted HR (95% CI) |  |  | 1.15 (0.81 - 1.64) | 1.88 (1.29 - 2.73) | Ref |  |  | 1.96 (1.59 - 2.41) | 2.13 (1.65 - 2.76) | Ref |
| Age and gender adjusted HR (95% CI) |  |  | 1.13 (0.80 - 1.61) | 1.86 (1.28 - 2.71) | Ref |  |  | 1.91 (1.56 - 2.35) | 2.10 (1.62 - 2.72) | Ref |
| Age, sex and ALTCA adjusted HR (95% CI) |  |  | 1.03 (0.73 - 1.47) | 1.59 (1.09 - 2.31) | Ref |  |  | 1.68 (1.36 - 2.06) | 1.74 (1.35 - 2.26) | Ref |
| Age, sex, ALTCA and education adjusted HR (95% CI) |  |  | 1.04 (0.74 - 1.48) | 1.59 (1.09 - 2.32) | Ref |  |  | 1.68 (1.37 - 2.07) | 1.75 (1.35 - 2.27) | Ref |
| Estimates with less than 10 deaths in either the vaccinated or reference group are not shown | | | | | | | | | | |

### **Matched Analysis Table (S48)**

| **Table S48:** Hazard ratios (HR) with 95% confidence intervals (95% CI) for all-cause mortality, cancer deaths and deaths from external causes according to number of SARS-CoV-2 vaccine doses in the two weeks after vaccination with controls matched based on age group, sex, education and ALTCA status, including age adjusted values. | | | | |
| --- | --- | --- | --- | --- |
|  | All-cause mortality | | | |
|  | Four vaccine doses | Unvaccinated | Three vaccine doses | Unvaccinated |
| Deaths (n) | 211 | 788 | 1185 | 4780 |
| Events per 100,000 person days | 3.71 | 13.87 | 1.91 | 7.78 |
| Crude HR (95%CI) | 0.27 (0.23 - 0.31) | Reference | 0.24 (0.23 - 0.26) | Reference |
| Age adjusted HR (95% CI) | 0.27 (0.23 - 0.31) | Reference | 0.25 (0.23 - 0.26) | Reference |
|  | Two vaccine doses | Unvaccinated | One vaccine dose | Unvaccinated |
| Deaths (n) | 1251 | 4319 | 1299 | 4049 |
| Events per 100,000 person days | 1.61 | 5.58 | 1.64 | 5.11 |
| Crude HR (95%CI) | 0.29 (0.27 - 0.31) | Reference | 0.32 (0.30 - 0.34) | Reference |
| Age adjusted HR (95% CI) | 0.29 (0.27 - 0.31) | Reference | 0.32 (0.30 - 0.34) | Reference |
|  | Cancer | | | |
|  | Four vaccine doses | Unvaccinated | Three vaccine doses | Unvaccinated |
| Deaths (n) | 25 | 126 | 167 | 893 |
| Events per 100,000 person days | 0.44 | 2.22 | 0.27 | 1.45 |
| Crude HR (95%CI) | 0.20 (0.13 - 0.30) | Reference | 0.18 (0.16 - 0.22) | Reference |
| Age adjusted HR (95% CI) | 0.20 (0.13 - 0.30) | Reference | 0.19 (0.16 - 0.22) | Reference |
|  | Two vaccine doses | Unvaccinated | One vaccine dose | Unvaccinated |
| Deaths (n) | 222 | 1100 | 182 | 1033 |
| Events per 100,000 person days | 0.29 | 1.42 | 0.23 | 1.3 |
| Crude HR (95%CI) | 0.20 (0.17 - 0.23) | Reference | 0.18 (0.15 - 0.21) | Reference |
| Age adjusted HR (95% CI) | 0.20 (0.17 - 0.23) | Reference | 0.18 (0.15 - 0.21) | Reference |
|  | External Causes | | | |
|  | Four vaccine doses | Unvaccinated | Three vaccine doses | Unvaccinated |
| Deaths (n) | 13 | 37 | 76 | 199 |
| Events per 100,000 person days | 0.23 | 0.65 | 0.12 | 0.32 |
| Crude HR (95%CI) | 0.35 (0.19 - 0.66) | Reference | 0.38 (0.29 - 0.49) | Reference |
| Age adjusted HR (95% CI) | 0.35 (0.19 - 0.66) | Reference | 0.38 (0.29 - 0.49) | Reference |
|  | Two vaccine doses | Unvaccinated | One vaccine dose | Unvaccinated |
| Deaths (n) | 95 | 185 | 107 | 203 |
| Events per 100,000 person days | 0.12 | 0.24 | 0.13 | 0.26 |
| Crude HR (95%CI) | 0.51 (0.40 - 0.66) | Reference | 0.53 (0.42 - 0.67) | Reference |
| Age adjusted HR (95% CI) | 0.51 (0.40 - 0.65) | Reference | 0.52 (0.42 - 0.66) | Reference |
| While matched based on age groups, age adjustment addresses the possibility of heterogenous age distributions within age groups | | | | |

### **Supplementary Discussion Table (S49)**

| **Table S49:** Selected findings of previous studies with comparable approaches assessing HVE | | | | | | | | |
| --- | --- | --- | --- | --- | --- | --- | --- | --- |
| **Study** | **population** | **analysis** | **covariates** | **n deaths (vaccinted/control)** | **time-window/ follow-up** | **dose vaccinated** | **dose control** | **rates** |
| Riedmann et al. 2025^1^ | Nationwide analysis of individuals with a previous documented infection | all-cause mortality: quarter-yearly outcomes | adjusted for: age, sex and nursing home residency | 283/423 | quarter 4 2021 | 1 | unvaccinated | HR: 0.66 (0.57 - 0.77) |
|  |  |  |  | 594/423 | quarter 4 2021 | 2 | unvaccinated | HR: 0.63 (0.55 - 0.71) |
|  |  |  |  | 415/423 | quarter 4 2021 | 3 | unvaccinated | HR: 0.53 (0.45 - 0.62) |
|  |  | all-cause mortality: matched 14-day post vaccination outcomes | matched for: age-group (5 years), sex and nursing home residency | 150/456 | 2021 - 2023, first 14 days after day of vaccination | 1 | unvaccinated | HR: 0.33 (0.27 - 0.40) |
|  |  |  |  | 136/568 | 2021 - 2023, first 14 days after day of vaccination | 2 | unvaccinated | HR: 0.24 (0.20 - 0.29) |
|  |  |  |  | 126/419 | 2021 - 2023, first 14 days after day of vaccination | 3 | unvaccinated | HR: 0.30 (0.25 - 0.37) |
|  |  |  |  | 223/505 | 2021 - 2023, first 14 days after day of vaccination | 4 or more | unvaccinated | HR: 0.44 (0.38 - 0.52) |
| Lyth et al. 2026^2^ | Total population aged ≥65 years, Southeast Sweden | all-cause mortality, October 1, 2024 to March 31, 2025. | adjusted for: age group, sex, immunosuppression status, Charlson Comorbidity Index and country of birth | 2607/2215 | Oct 1 2024 - Mar 31 2025 | 2024–2025 booster | no booster | HR: 0.43 (0.40 - 0.45) |
| Chemaitelly et al. 2025^3^ | National matched cohorts, Qatar (two-dose vs unvaccinated: n=812,583 each; three-dose vs two-dose: n=330,568 each) | non-COVID-19 mortality: two-dose (primary series) vs unvaccinated (matched cohort) | matched/adjusted for: sex, 10-year age group, nationality, coexisting conditions, prior infection status, calendar week of vaccination | 237/306 | Jan 5 2021 - Apr 9 2024 (overall) | 2 | unvaccinated | HR: 0.76 (0.64 - 0.90) |
|  |  |  |  | not reported | first 6 months of follow-up | 2 | unvaccinated | HR: 0.35 (0.27 - 0.46) |
|  |  | non-COVID-19 mortality: three-dose (booster) vs two-dose (matched cohort) | matched/adjusted for: sex, 10-year age group, nationality, coexisting conditions, prior infection status, calendar week of vaccination | 132/147 | Jan 5 2021 - Apr 9 2024 (overall) | 3 | 2 | HR: 0.85 (0.67 - 1.07) |
|  |  |  |  | not reported | first 6 months of follow-up | 3 | 2 | HR: 0.31 (0.20 - 0.50) |
| Note that approaches and time windows differ starkly which may effect the specific estimates hazard ratios. Riedmann et al. 2025 most closley followes the here presented approach. | | | | | | | | |

### **Supplementary Figures**

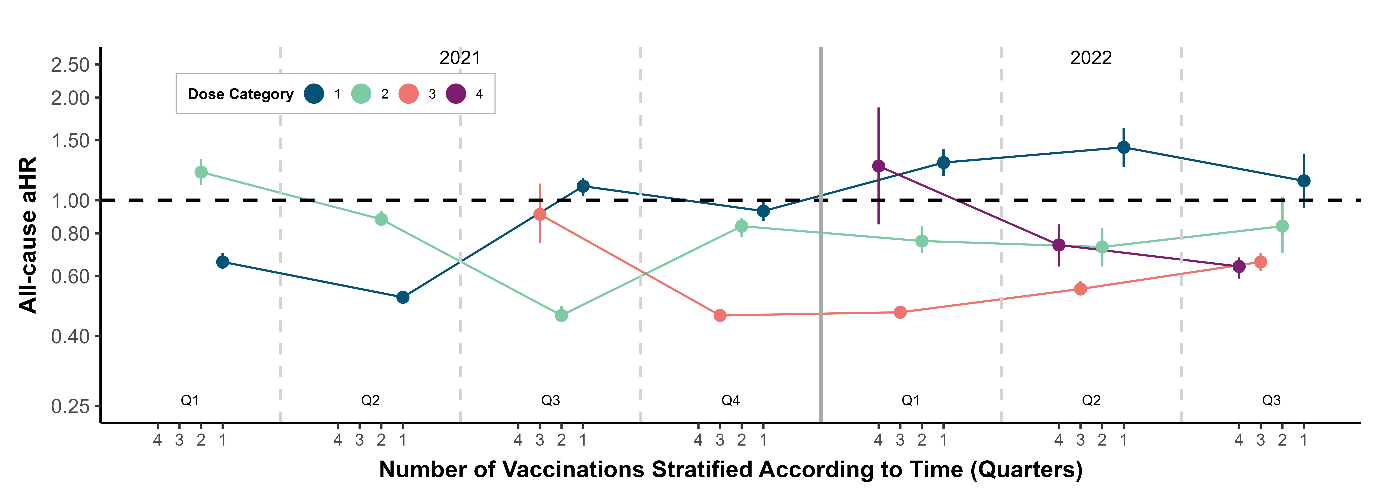

**Figure S1:** Age, sex and ALTCA recipiency adjusted all-cause-mortality HRs and their CIs for 1, 2, 3 and 4 vaccinations over the whole observation period split into three-month periods. The reference group for every dose is the preceding dose. See Table S13 for details and crude estimates.
